# Calibration and processing sensitivity of time-embedded three-dimensional electrocardiographic descriptors: a two-dataset benchmark

**DOI:** 10.64898/2026.09.14.26363023

**Authors:** Alejandro Jesús Bermejo Valdés

**Affiliations:** Emergency Medical Service 061, Riojan Health Service, Spain

**Keywords:** electrocardiography, torsion, curvature, calibration, numerical differentiation, reproducibility

## Abstract

Time-embedded electrocardiographic descriptors combine high-order derivatives in nonlinear ratios, making their numerical calibration essential. We analyzed 200 LUDB participants (1,748 QRS windows) and a fixed external sample of 1,000 PTB-XL participants (9,735 windows). Four prespecified derivative operators were evaluated at three nominal Butterworth cutoffs; a fifth, regularized operator was added post hoc. Extensions added transfer functions, matched response crossings and a Gaussian transient with exact derivatives. The expanded calibration comprised 1,005 mathematical signal/noise realizations. At 150 Hz, spectral differentiation (SPEC) increased external participant-level mean absolute torsion by a median 78.97% relative to successive finite differences (FD; 95% confidence interval 77.39–80.53). Reducing the cutoff to 40 Hz yielded weak rank preservation: Spearman correlations were 0.045 for SPEC and 0.099 for FD. Matching each derivative’s first normalized −3-dB crossing at 20 Hz reduced the external SPEC–FD median difference to 0.016%, but introduced 4.26–4.70% noiseless error on the Gaussian transient. Agreement was therefore not accuracy against the known target. For the Gaussian transient with 20-microvolt noise, relative root mean squared errors at 150 Hz were 719.52% for SPEC and 99.96% for the regularizer; the latter error remained comparable to the target magnitude, and regularization also introduced noiseless bias. Almost-curvature approximated a normalized voltage slope under the specified coordinates and did not converge to classical curvature under reparametrization. Explicit transfer functions, absolute descriptor levels and analytic targets explain important processing effects while separating numerical agreement from clinical validity. No universally optimal estimator or diagnostic benefit is established.

## 1. Introduction

Recent three-dimensional ECG representations combine standard precordial voltages with a temporal coordinate, allowing geometric descriptors and projections to be examined without additional electrodes. Published applications include a first geometric framework, R-wave progression and rotation, and the appearance of terminal QRS deflections [1–3]. A related preprint also investigated torsion and ischemia classification [4]. These reports motivate scrutiny of how the resulting descriptors are defined and calculated before their clinical meaning is inferred.

Geometric ECG analysis is not new. Curvature, planarity and related quantities have been studied in vectorcardiography [5, 6], and the dependence of ECG measurements on acquisition and processing is a longstanding standardization concern [7]. A trajectory containing two voltages and time, however, is not a directly measured anatomical cardiac vector. Its numerical geometry depends on the relative scales assigned to voltage and time. A rotation of the displayed curve can reveal a feature of the representation without independently identifying its physiological origin.

Numerical differentiation introduces a separate problem. Higher derivatives emphasize high-frequency components, including noise. Regularization and the balance between fidelity and smoothness have established mathematical treatments [8, 9]. Savitzky-Golay operators also have order-dependent frequency responses that matter when several derivatives are combined in a nonlinear ratio [10]. Less variation across filters therefore cannot be equated with smaller error against a known curve.

We constructed a reproducible measurement benchmark that separates coordinate conventions, numerical calibration, and processing sensitivity in real ECGs. A common trigonometric interpolant was used to obtain coherent first, second and third derivatives, following established spectral principles [11]. The benchmark combines an audit of coordinate dependence, derivative calibration against analytic trajectories, and a fixed external assessment of processing sensitivity. These components distinguish errors against known mathematical targets from disagreements among measurements on real signals. The aim was to assess the spectral implementation against conventional operators and replicate processing effects in an external cohort. The study did not develop a diagnostic classifier or assume that spectral differentiation would be uniformly preferable.

## 2. Methods

### 2.1. Study design and datasets

This computational study followed exploratory development in LUDB and an external assessment in PTB-XL. The final phase protocol and external selection were fixed before its descriptor calculations. An annotation-reader amendment, documented before external calculations, specified ECGDeli event labels after structural inspection of a non-test record. The protocol was locally time-stamped and hashed, not publicly preregistered. Prior LUDB exploration informed the question, so development findings remain exploratory.

LUDB version 1.0.1 supplied 200 ten-second, 12-lead recordings sampled at 500 Hz, with waveform annotations [12, 13]. Exactly the 1,748 QRS windows selected during the internal exploratory phase of this study were retained; these are not windows reproduced from reference 1. External waveforms came from PTB-XL version 1.0.3 [14, 15], with automatic ECGDeli annotations distributed in PTB-XL+ version 1.0.1 [16–18]. Both resources were accessed through PhysioNet [19]. The external metadata test fold contained 2,198 records from 1,904 people. We retained the lowest ECG identifier per person, ranked patients by a fixed SHA-256 rule, and selected the first 1,000. This fixed computational sample size was not derived from a clinical power calculation. The selection rule prohibited replacement; in the resulting sample, every selected participant retained eligible windows.

Record identifiers, source versions and downloaded files were archived. External files were checked against published SHA-256 manifests. Waveforms were read in physical millivolts, with checks for 500 Hz sampling, 5,000 samples, 12 channels and finite values. V1 and V6 were fixed as right and left precordial examples before external descriptor calculations; this two-voltage embedding was not selected by optimizing external results and does not reproduce every earlier polar 3D representation. Whole-record median centering removed constant offsets. All four descriptors depend only on derivatives, so centering does not affect their mathematical values and is reported for completeness. No participant-specific amplitude normalization, which changes the geometry, was applied.

### 2.2. QRS windows and coordinate conventions

For LUDB, complete QRS onset, peak and offset annotations in V1 and V6 were mutually matched by the nearest annotated peak within 100 ms. The window spanned the earlier onset and later offset. Windows outside 40-300 ms or within one second of a recording edge were excluded. For ECGDeli, explicit QRS onset, R peak and QRS offset labels were parsed at their declared 500 Hz sampling frequency. Each peak required a unique flanking onset and offset within neighboring-peak intervals; incomplete or ambiguous triples were counted. The same cross-lead matching, duration and edge rules were then applied. A separate computational reconstruction from both annotation sources reproduced every retained window: all starts were at least sample 500 and all ends at most sample 4499. LUDB lost 28 windows at the initial edge and 35 at the final edge; PTB-XL lost 1,321 and 1,132. LUDB annotations were already concentrated centrally: median first complete QRS onset was 1.330 s in both leads and median last offset was 8.635 s in V1 and 8.632 s in V6. These are observed annotation landmarks, not a known continuous coverage interval. Table S12 supplies their distributions and per-record audit. These external boundaries were algorithmic, not manually adjudicated. The first five hash-ranked records were visually inspected for waveform alignment without manual correction or outcome-based exclusion.

The numerical coordinates were voltage divided by 1 mV on both lead axes and time divided by 1 ms. These normalized coordinates are dimensionless. Classical curvature and torsion are expressed in inverse normalized-coordinate units, hence are numerically dimensionless; their parameter units cancel. Almost-curvature is a numerical index tied additionally to the specified differentiation parameter. None is reported in anatomical inverse millimeters or interchangeable with vectorcardiographic units. QRS boundaries were fixed across processing conditions. Classical curvature and signed torsion were calculated from the first three derivatives of the curve:

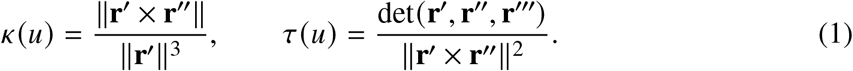

The primary descriptor was the temporal mean of absolute torsion within each QRS. Secondary descriptors were mean classical curvature, mean equation-6 almost-curvature from the original report [1], and denominator-weighted mean absolute torsion. The last quantity divides the integral of the absolute determinant by the integral of the squared cross-product norm and is a different estimand from the temporal mean. Per-window values were integrated by the trapezoidal rule and summarized by their median within each participant. Formulas and degeneracy handling are detailed in the supplementary methods.

### 2.3. Derivative operators and mathematical calibration

Fourth-order Butterworth low-pass filters were applied in both directions at nominal cutoffs of 40, 100 and 150 Hz, using the same implementation for every operator. The nominal frequency denotes the single-pass design; applying the filter twice gives approximately −6 dB there. No additional high-pass or notch filter was introduced. These manipulations compare processing specifications and do not restore frequencies absent from the acquired files.

The four operators were successive finite differences with a 2-ms step (FD); direct Savitzky-Golay derivatives of orders 1-3, degree 3 and an 11-sample window (SG); Fourier differentiation of a reflected common trigonometric interpolant (SPEC); and order-zero SG smoothing followed by SPEC (SG0SPEC). For SPEC, even reflection yielded 2*N* − 2 samples, Fourier coefficients were multiplied by the corresponding powers of imaginary angular frequency in radians/ms, and derivatives were cropped to the original record. The Nyquist coefficient was removed consistently for all derivative orders. No settings were optimized against external measurements. SG and SG0SPEC include additional smoothing; equal nominal Butterworth cutoffs do not imply identical effective bandwidths. Even reflection need not have continuous first derivatives at its endpoints. A one-second guard reduces exposure to edges but does not guarantee uniform spectral convergence or eliminate global boundary effects.

Calibration used three unit-radius helices at 10, 30 and 60 Hz and a phase-modulated circular trajectory with analytically available derivatives. Eight-second signals were sampled at 500 Hz and evaluated over the central 2-6-second interval. Independent Gaussian noise with standard deviations of 0, 5 or 20 microvolts was added to each voltage coordinate. There was one deterministic realization without noise and 100 shared realizations per nonzero noise level, producing 804 signal/noise realizations. These were mathematical controls, not synthetic patients, and their noise levels were not estimated from the clinical recordings.

Every operator/filter combination was compared with each descriptor’s analytic value using signed relative bias and relative root mean squared error (RMSE). For the helices, the exact post-filter target was additionally calculated from the known Butterworth amplitude response. This separates changing the curve by filtering from derivative estimation error. A periodic-input spectral calculation and explicit Fourier summation provided independent numerical checks of the implementation. The term independent refers to a separate computational formulation, not to an independent research team.

### 2.4. Transfer functions and post hoc extensions

Revision analyses were specified in a separate, locally hashed plan after the original results were available; they are post hoc, not part of the original external confirmation. For interior successive central differences, the order-*k* derivative response relative to the ideal derivative is *s*^*k*^, with *s* = sin(*ωh*)/(*ωh*), *h* = 2 ms. For a filtered helix of radius *R*, this gives

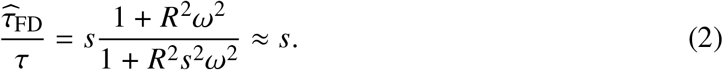

The approximation requires a small slope correction. A multicomponent ECG has no single harmonic frequency; substituting the nominal filter cutoff is a heuristic, not an exact correction. We computed the normalized response of each derivative order, including the double-pass Butterworth filter, and its first −3-dB crossing.

A fifth operator (TIK) minimized squared reconstruction error plus a squared third-derivative penalty on the reflected periodic interpolant. Its common smoothing response was fixed as

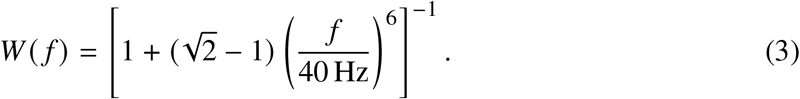

All three derivatives were computed from this same regularized curve. This transparent quadratic baseline evaluates regularization directly; it is not an implementation of total-variation regularization and was not tuned to minimize external differences.

In a separate comparison, each of the five operators after nominal 150-Hz filtering received an additional order-specific Gaussian taper so that every normalized derivative response first crossed 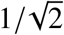 at 20 Hz. This value lies below all native crossings. Matching one crossing controls a bandwidth criterion, not the entire response shape or noise gain; order-specific tapers also need not be derivatives of one common curve. The 40-Hz regularizer and 20-Hz matching parameters were fixed before these added calculations, without an optimization search.

We added a QRS-like transient formed by sums of three Gaussians per voltage coordinate, with exact derivatives and a fixed central 160-ms evaluation window. The same 0-, 5- and 20-microvolt noise design added 201 realizations, for 1,005 in total. The expanded benchmark contains 20 processing configurations: 15 native operator/cutoff combinations and five additional matched configurations. All 1,200 model/noise/configuration/descriptor summary rows were retained. The supplementary methods specify the Gaussian coefficients and regularization.

### 2.5. Coordinate audits and sensitivity analyses

At 150 Hz with FD, we tested a proper 90-degree rotation in the voltage plane and a linear reparametrization from milliseconds to seconds while retaining exactly the same coordinates. Classical curvature and torsion should be preserved under these operations. Separately, changing the time coordinate from *t* /(1 ms) to *t*/(100 ms) deliberately produces a different geometry; it was not treated as a failed invariance test. The equation-6 index was evaluated under the same conditions, without assuming it had the invariances of classical curvature.

QRS boundary sensitivity was assessed by shifting the onset and offset by minus or plus 10 ms in all four combinations, for FD and SPEC at 150 Hz. The maximum absolute percentage change among the four participant summaries was reported descriptively. For SPEC, reflected and periodic full-record extensions were also compared using identical QRS windows. Neither analysis estimates the unknown true biological curve. Post hoc audits decomposed almost-curvature into its slope and second-derivative terms. A common-duration sensitivity analysis retained windows from 80 to 160 ms, recomputed participant medians and omitted people without retained windows; the range was fixed before this calculation.

### 2.6. Statistical analysis and verification

The primary external contrast was SPEC versus FD at 150 Hz for the participant summary of absolute torsion. Secondary comparisons were SG and SG0SPEC versus FD at 150 Hz, and 40 versus 150 and 100 versus 150 Hz within each operator, for all four descriptors. All 88 planned cohort/descriptor/comparison combinations were retained. Relative change was 100 times candidate minus reference, divided by reference. Median absolute relative change and Spearman correlation were reported alongside signed changes; correlation was not interpreted as agreement.

Paired percentile bootstrap confidence intervals for median percentage changes used 5,000 participant resamples with a fixed random seed. Participants, not beats, were the sampling units. Intervals were descriptive and not adjusted for the secondary comparisons; no significance threshold selected the preferred method. Undefined values were explicitly counted and paired summaries used complete participants. Checks covered official file hashes, selection reproducibility, units, algebraic identities, direct Fourier sums, absence of duplicate measurement units, condition counts, participant aggregation and reproduction of FD results from this study’s internal LUDB exploratory phase. Computation used Python 3.12.14, NumPy 2.3.5, SciPy 1.18.1, pandas 2.2.3 and WFDB 4.3.1.

Post hoc comparisons used the same participant-level procedure with a separate fixed seed. Absolute levels are participant medians with interquartile ranges. After undefined matched measurements were counted, an additional comparison restricted every matched operator to the same valid windows. Gaussian derivatives were checked through Hermite polynomials and TIK through a separate dense normal-equation solution. Figures were plotted from saved result tables with Matplotlib 3.11.2; their source data and plotting code are supplied.

## 3. Results

### 3.1. Included recordings and processing comparisons

All 200 LUDB participants contributed 1,748 QRS windows. Of 1,000 selected external participants, 1,000 contributed 9,735 eligible windows and none were excluded from participant-level analysis. Table 1 reports the acquisition and window counts. Every eligible window was evaluated under the same original 12 operator/filter combinations, with no undefined temporal absolute-torsion measurements. Added native TIK measurements were also all defined. Matching produced 17 undefined measurements across six PTB-XL windows (FD 5, SG 6, SPEC 5, SG0SPEC 1, TIK 0), with none in LUDB. Every participant retained valid windows. Inclusion details and verification outputs accompany the supplementary data archive. Figure 4 and Supplementary Table S8 report inclusion and the complete duration distributions. The external 12,188 matched candidate windows lost 2,453 at the edges and none to the duration rule. In LUDB, 1,811 matched candidates lost 63 at the edges. Complete original and added results are identified in Supplementary Tables S1–S13.

**Table 1.** Cohort and measurement characteristics.

| Characteristic | LUDB | PTB-XL |
| --- | --- | --- |
| Participants selected | 200 | 1,000 |
| Participants included | 200 | 1,000 |
| Participants excluded | 0 | 0 |
| Eligible QRS windows | 1,748 | 9,735 |
| Median QRS per participant | 8 | 9 |
| Median window duration (ms) | 102 | 150 |
| Sampling frequency | 500 Hz | 500 Hz |
| Recording duration | 10 s | 10 s |
| QRS annotation source | LUDB manual annotations | Published ECGDeli annotations |

One recording was used per participant. The external sample was selected before descriptor analysis. Window duration is summarized across eligible windows, whereas the principal descriptor statistics use participant-level summaries.

The primary external SPEC-versus-FD contrast at 150 Hz was 78.97% (95% CI 77.39 to 80.53), with Spearman 0.365. The corresponding LUDB change was 92.27% (95% CI 87.18 to 96.30), with Spearman 0.573. These are differences between computations on the same recordings, with a calculable transfer-function contribution. They do not measure error against clinical ground truth. Absolute levels for all five native operators and three cutoffs are reported in Table 3; all four descriptors, including matched conditions, appear in Supplementary Table S6. Table 2 also reports SG and SG0SPEC contrasts, including changes in participant ordering.

**Table 2.** Absolute torsion differences between operators at 150 Hz.

| Cohort | Operator | $n$ | Median change (95% CI), % | Median absolute change, % | Spearman $\rho$ vs FD, 150 Hz |
| --- | --- | --- | --- | --- | --- |
| LUDB | SG | 200 | -34.93 (-37.53 to -33.38) | 34.93 | 0.553 |
| LUDB | SPEC | 200 | 92.27 (87.18 to 96.30) | 92.27 | 0.573 |
| LUDB | SG0SPEC | 200 | -7.56 (-9.20 to -5.37) | 11.10 | 0.697 |
| PTB-XL | SG | 1,000 | -37.76 (-38.54 to -36.78) | 37.76 | 0.301 |
| PTB-XL | SPEC | 1,000 | 78.97 (77.39 to 80.53) | 78.97 | 0.365 |
| PTB-XL | SG0SPEC | 1,000 | -4.69 (-5.56 to -3.89) | 9.20 | 0.345 |

FD is the reference. Positive signed change indicates a higher candidate value. For each participant, the outcome is the median across their QRS windows of the temporal mean absolute torsion; comparisons are paired within participant. These differences include operator-response effects and are not clinical measurement errors.

**Table 3.** Absolute levels of temporal mean absolute torsion.

| Operator | Cutoff (Hz) | LUDB ($n = 200$) | PTB-XL ($n = 1,000$) |
| --- | --- | --- | --- |
| FD | 40 | 0.1367 [0.1248–0.1517] | 0.1516 [0.1408–0.1627] |
| FD | 100 | 0.2353 [0.2124–0.2583] | 0.2732 [0.2548–0.2926] |
| FD | 150 | 0.2779 [0.2569–0.3096] | 0.3214 [0.2992–0.3472] |
| SG | 40 | 0.1335 [0.1215–0.1470] | 0.1447 [0.1330–0.1568] |
| SG | 100 | 0.1742 [0.1537–0.2022] | 0.1915 [0.1720–0.2118] |
| SG | 150 | 0.1764 [0.1561–0.2051] | 0.2019 [0.1801–0.2245] |
| SG0SPEC | 40 | 0.1388 [0.1286–0.1527] | 0.1541 [0.1428–0.1660] |
| SG0SPEC | 100 | 0.1977 [0.1791–0.2207] | 0.2348 [0.2149–0.2527] |
| SG0SPEC | 150 | 0.2574 [0.2230–0.2952] | 0.3065 [0.2804–0.3326] |
| SPEC | 40 | 0.1432 [0.1300–0.1582] | 0.1579 [0.1466–0.1683] |
| SPEC | 100 | 0.3170 [0.2859–0.3517] | 0.3616 [0.3391–0.3863] |
| SPEC | 150 | 0.5346 [0.4770–0.5953] | 0.5764 [0.5352–0.6180] |
| TIK | 40 | 0.1370 [0.1257–0.1461] | 0.1518 [0.1407–0.1625] |
| TIK | 100 | 0.1543 [0.1402–0.1702] | 0.1675 [0.1563–0.1790] |
| TIK | 150 | 0.1543 [0.1405–0.1704] | 0.1681 [0.1563–0.1798] |

With FD, reducing the nominal cutoff from 150 to 40 Hz changed the participant summary of absolute torsion by −50.86% (95% CI −53.53 to −49.75) in LUDB and −53.24% (95% CI −53.78 to −52.76) in PTB-XL. For SPEC the corresponding changes were −73.28% (95% CI −74.29 to −72.34) and −72.84% (95% CI −73.13 to −72.51). Figure 1 presents all four original operators. In PTB-XL, rank correlations between 150 and 40 Hz were only 0.099 for FD and 0.045 for SPEC, indicating weak preservation of participant ordering. These correlations do not establish random ordering or diagnostic failure. The corresponding LUDB correlations were 0.242 and 0.168.

**Figure 1.**
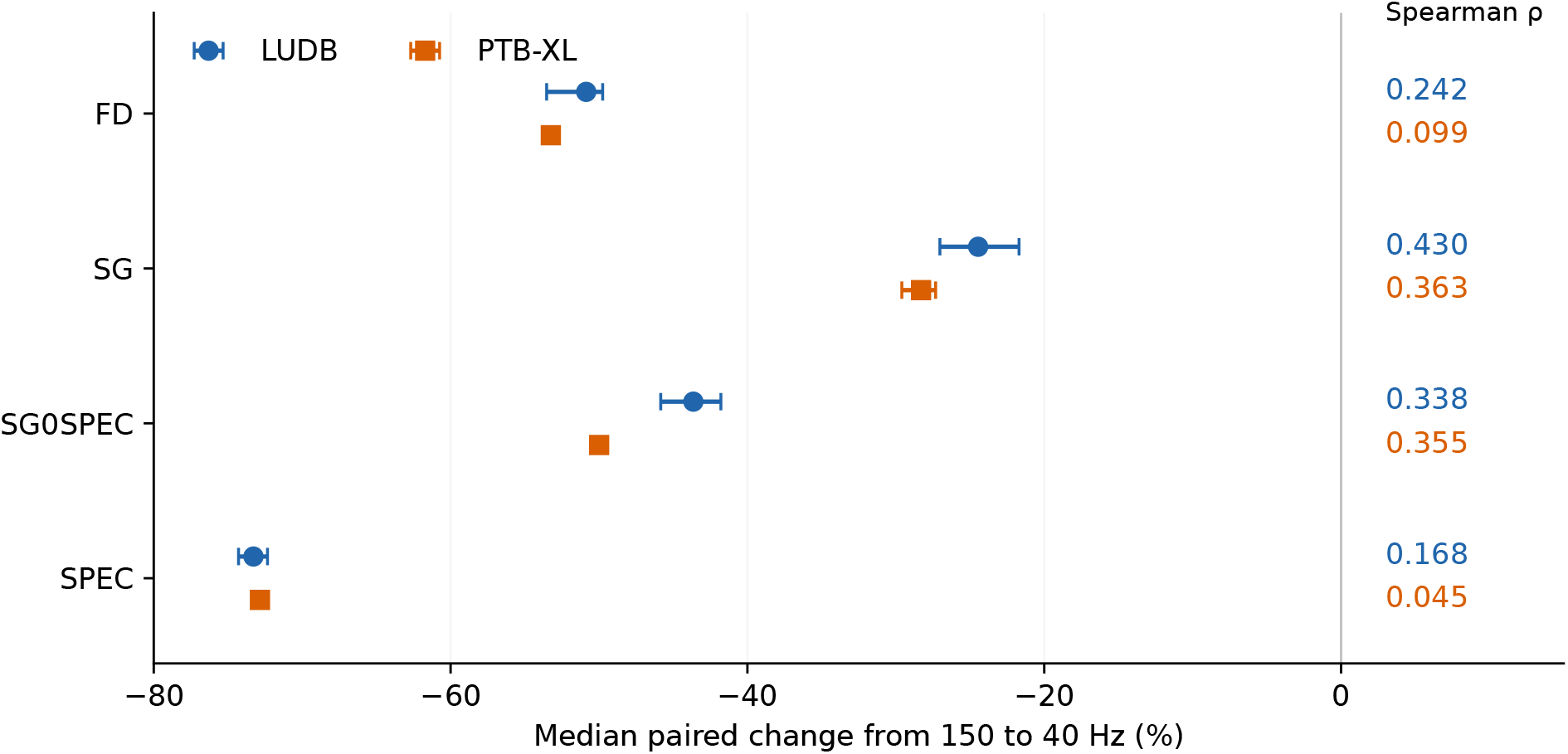
Low-pass filtering and participant rank preservation. Median paired percentage changes in temporal mean absolute torsion from a nominal cutoff of 150 to 40 Hz, with 95% participant-bootstrap intervals. Each value is the median of window summaries within a participant. The horizontal range includes every estimate and interval. Spearman correlations at right quantify rank preservation, not agreement or clinical discrimination. LUDB: 200 participants; PTB-XL: 1,000.

### 3.2. Transfer-function explanation and regularization

Evaluating *s* at the nominal cutoff frequency—a single-harmonic substitution, heuristic for a multicomponent transient rather than a property of the filter—gives 0.9584, 0.7568 and 0.5046 at 40, 100 and 150 Hz. The resulting approximate harmonic SPEC/FD increases are 4.34%, 32.13% and 98.20%. Observed median paired differences at 40, 100 and 150 Hz were 4.16%, 34.44% and 92.27% in LUDB, and 3.66%, 32.60% and 78.97% in PTB-XL. At 150 Hz, the external observation was 19.23 percentage points below the harmonic approximation. These are medians of within-participant percentage changes, not percentage changes calculated from cohort medians: at 40 Hz the latter were 4.80% and 4.15% using unrounded values. Both aggregation routes are supplied in Table S11. A cutoff-edge heuristic combining SPEC’s within-person cutoff ratio with the FD harmonic factors predicted FD changes from 150 to 40 Hz of −49.25% and −48.41%, compared with observed −50.86% and −53.24%. However, the median individual absolute prediction errors were 5.02 and 5.75 percentage points; at 100 Hz they were 10.56 and 12.12 points. Agreement between cohort medians is therefore insufficient evidence for an individual correction.

Figure 3 and Supplementary Table S5 expose the different native responses. At nominal 150 Hz, normalized third-derivative crossings were 65.48 Hz for FD, 36.61 Hz for SG, 49.32 Hz for SG0SPEC, 141.53 Hz for SPEC and 40.00 Hz for TIK. After matching the 20-Hz crossings, median SPEC–FD differences were 0.011% in LUDB and 0.016% in PTB-XL, with correlations above 0.9999. Median absolute differences were 0.017% and 0.033%. The external matched TIK–FD median difference was 0.943%, with a median absolute difference of 5.43% and correlation 0.721. Thus, agreement improved greatly for SPEC and FD but remained dependent on response shape and operator. The plotted responses are consistent with this pattern: FD approaches unity at low frequencies, so its matching taper closely approximates the SPEC chain, whereas TIK retains its additional sixth-power roll-off around 40 Hz. Matching one crossing does not remove that shape difference; it is not a unique causal decomposition of the residual ECG discrepancy. Restricting all five matched operators to 9,729 common valid external windows preserved the SPEC–FD median difference of 0.016% (Supplementary Table S11).

**Figure 2.**
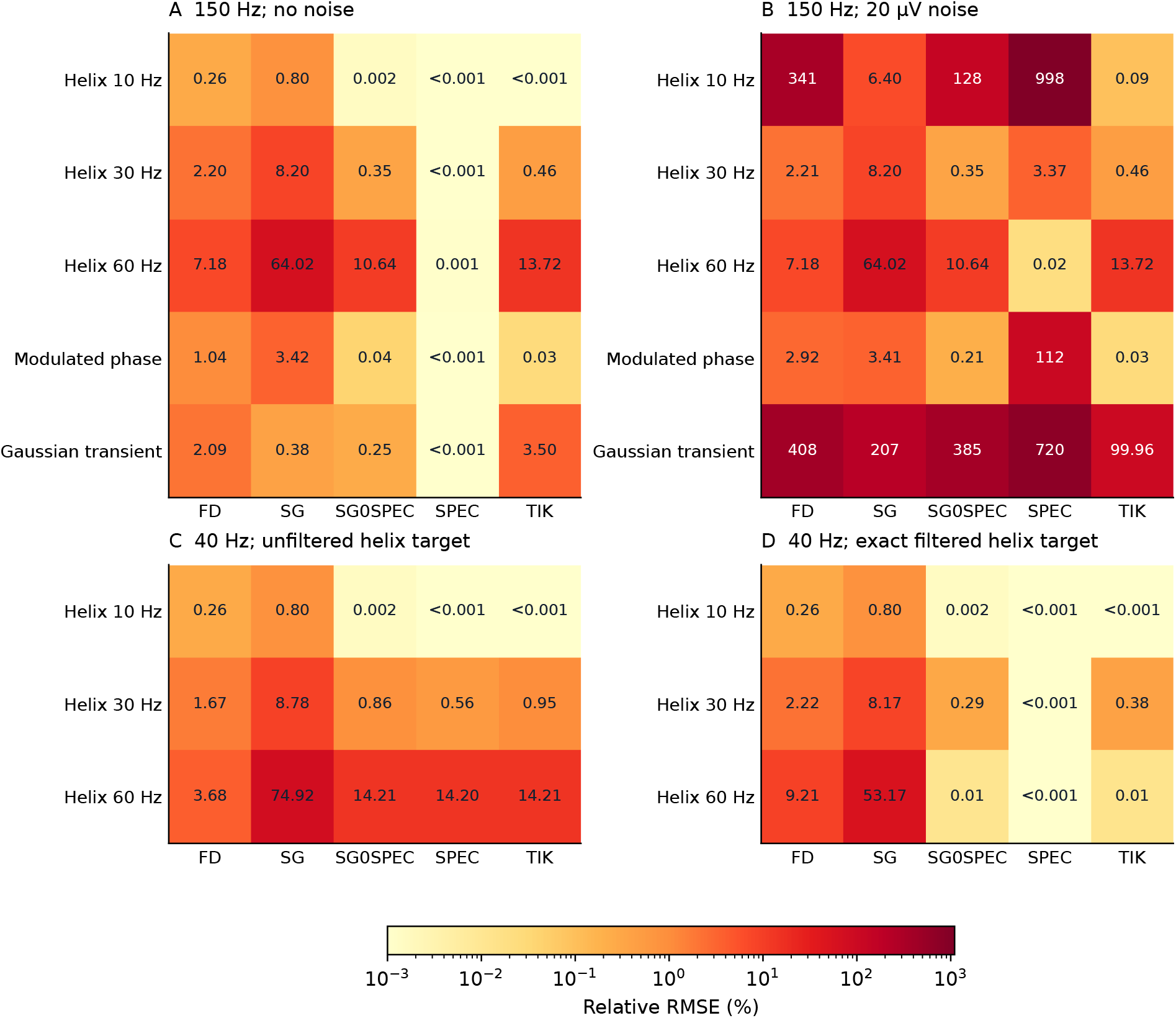
Analytic calibration with distinct targets. Relative RMSE (%) of temporal mean absolute torsion. Panels A and B use the unfiltered analytic targets at a 150-Hz nominal cutoff, without noise and with 20-*μ*V independent Gaussian noise per voltage coordinate. Panels C and D show the same noiseless 40-Hz helix estimates against, respectively, the unfiltered and exact steady-state Butterworth-filtered trajectories. These targets exclude any additional operator smoothing. One realization is used without noise and 100 per noisy condition. Labels below 0.001% are censored for display only; shading has a common logarithmic scale. TIK is the fixed third-derivative Tikhonov regularizer. Complete bias and RMSE, including 5-*μ*V noise, are supplied in Supplementary Table S7.

**Figure 3.**
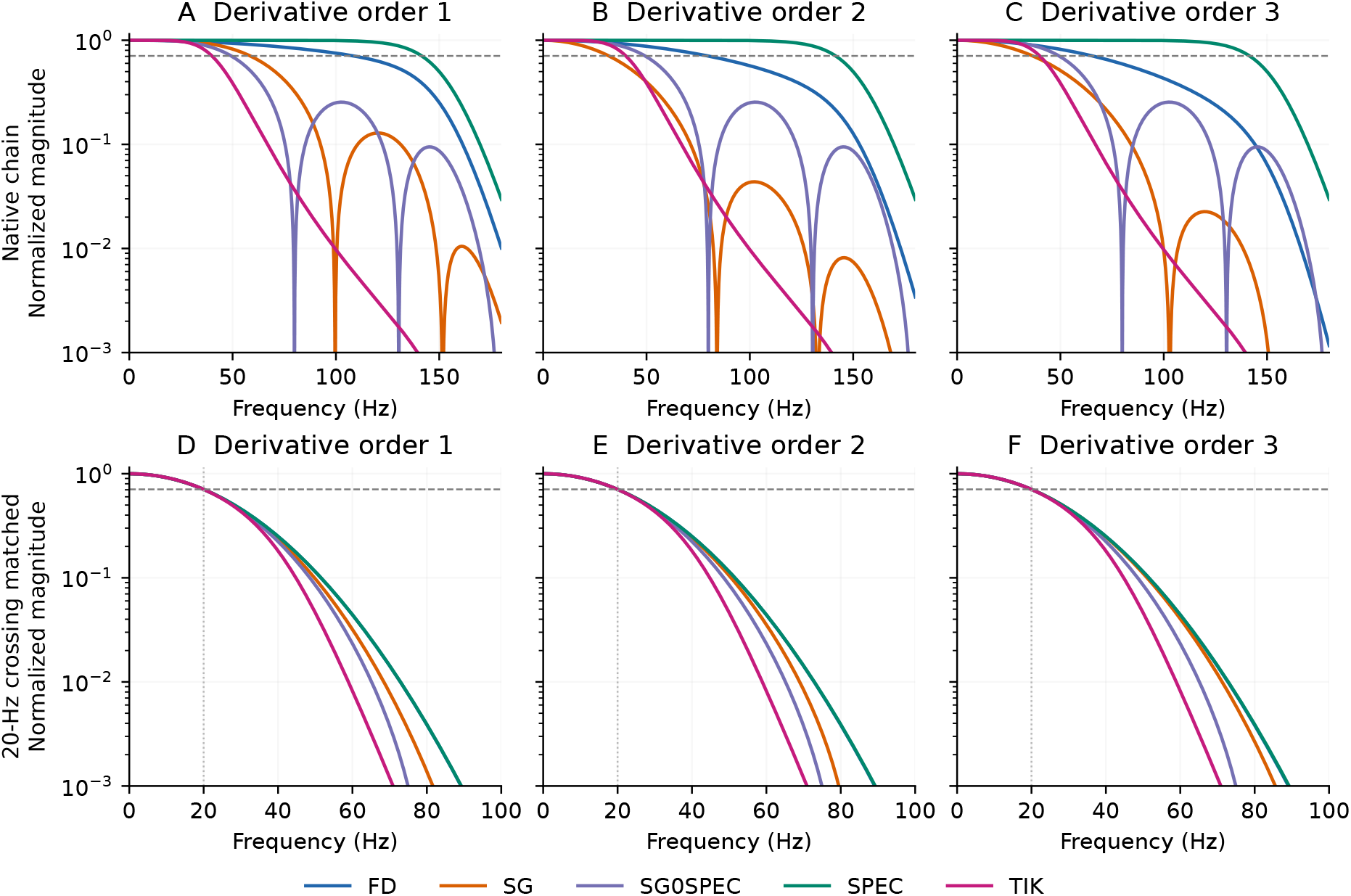
Effective derivative-chain responses. Magnitudes divided by the ideal derivative magnitude (2*π f*/1000)^*k*^, for orders *k* = 1, 2, 3. Panels A–C include the nominal 150-Hz double-pass Butterworth filter and each native operator. Panels D–F add orderspecific Gaussian tapers to place the first normalized −3-dB crossing at 20 Hz. Horizontal dashed lines mark 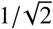; vertical dotted lines mark 20 Hz. Matching this crossing does not match the full response. Curves describe the interior, shift-invariant response; finite-record boundary behavior is assessed separately.

**Figure 4.**
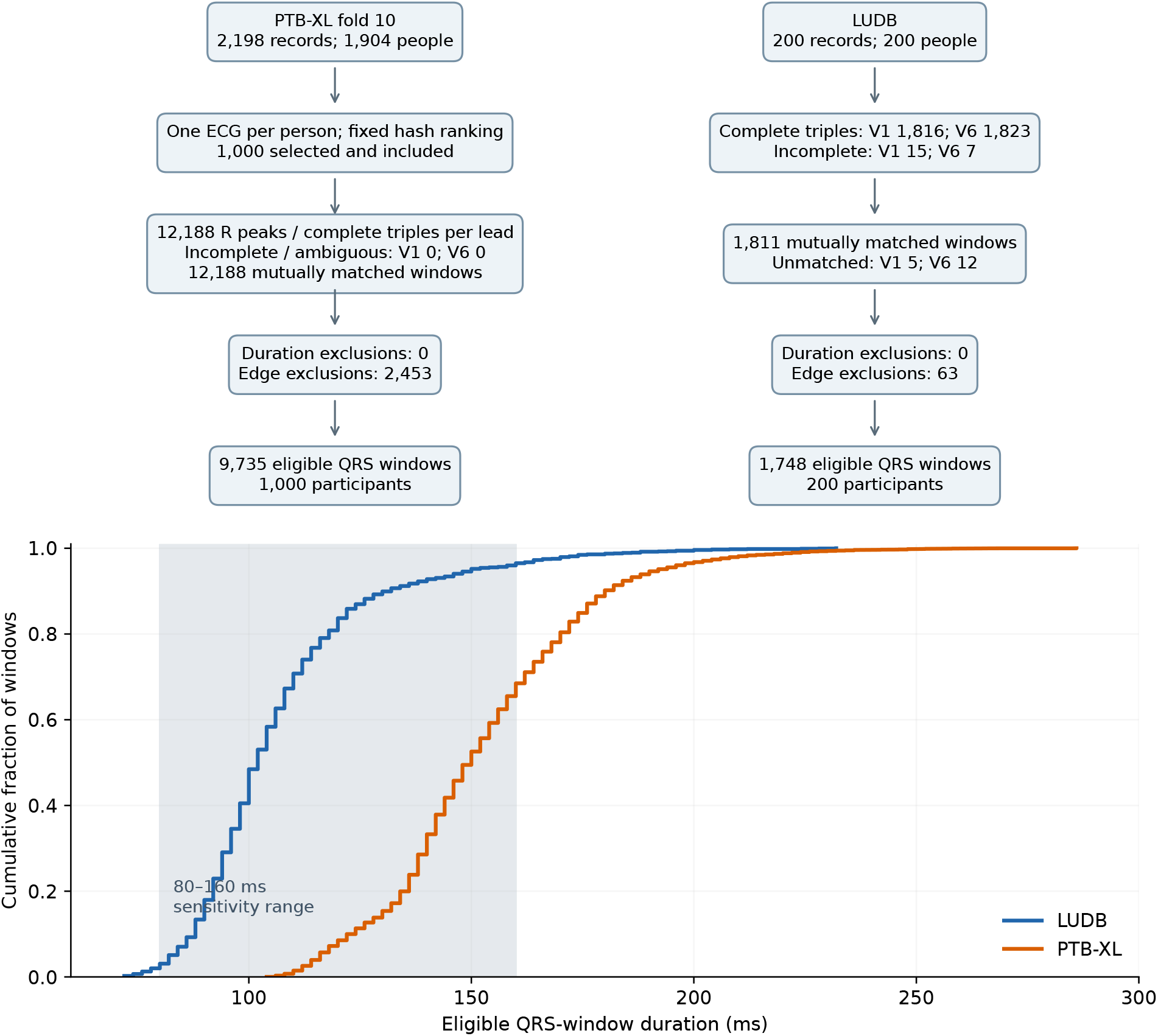
Inclusion flow and QRS-window durations. Complete triples comprise onset, peak and offset in one lead; mutual peak matching produces one cross-lead window. The same one-second guard was independently verified in both cohorts: LUDB exclusions were 28 initial and 35 final; PTB-XL exclusions were 1,321 initial and 1,132 final. LUDB annotated candidates were already concentrated centrally (Table S12). PTB-XL had zero incomplete or ambiguous triples in each lead. All 12,188 external pairs passed mutual-nearest matching within 100 ms; their absolute peak separation was a median 6 ms and a maximum 98 ms, not identically zero. No participant was excluded after window selection. The lower panel shows the empirical cumulative distribution across eligible windows, with the post hoc 80–160-ms sensitivity range shaded. It retains 1,652 windows in 196 LUDB participants and 6,668 windows in 930 PTB-XL participants. The restriction does not equate the duration distributions or annotation provenance.

### 3.3. Calibration against known trajectories

SPEC relative RMSE was below 0.002% for the four original noiseless controls at 150 Hz. FD errors in the 30- and 60-Hz helices were 2.20% and 7.18%; SG errors were 8.20% and 64.02%. The large noiseless SG error is consistent with its restricted derivative response, not noise suppression alone. At 20-microvolt noise, the 10-Hz-helix errors were 997.82% for SPEC, 340.77% for FD, 6.40% for SG, 128.18% for SG0SPEC and 0.09% for TIK. Regularization nevertheless imposed bias: TIK’s noiseless 60-Hz-helix error was 13.72%. Complete original calibration results appear in Table S4. SPEC’s noisy helix errors fell from 997.82% at 10 Hz to 3.37% at 30 Hz and 0.02% at 60 Hz. For a radius-*R* noiseless helix with *z* = *t*, the torsion denominator is *R*^2^*ω*^4^ (1 + *R*^2^*ω*^2^); therefore, fixed coordinate-noise amplitude does not imply equal perturbation relative to the derivative signal across frequencies. These controls do not establish a general monotonic noise-response law for multicomponent ECGs.

The target distinction is quantitative (Figure 2). For the noiseless 60-Hz helix after 40-Hz filtering, SPEC RMSE was 14.20% against the unfiltered curve and 0.000282% against the exact filtered curve. FD errors were 3.68% and 9.21%, respectively. Its smaller error against the unfiltered target reflects cancellation between filtering and differentiation effects, rather than a more accurate derivative of the filtered curve. Supplementary Table S7 reports both targets at all three cutoffs.

For the Gaussian transient at 150 Hz, noiseless errors were below 0.001% for SPEC, 2.09% for FD and 3.50% for TIK. SG improved from a 64.02% error on the 60-Hz helix to 0.38% on the transient. This small summary error concealed local deviations: integrated positive and negative errors in absolute torsion contributed +16.29% and −16.67% of the target integral, leaving a net −0.38%; the integrated absolute error was 32.96% (Table S13). A single-frequency derivative-ratio heuristic cannot predict this cancellation or independently validate the helix formula on a transient. At 20-microvolt noise, these increased to 719.52%, 408.29% and 99.96%; SG and SG0SPEC errors were 207.39% and 384.67%. At 5 microvolts, SPEC and TIK errors were 510.46% and 47.91%. The complete results retain this intermediate noise level. After 20-Hz crossing matching, the transient’s noiseless errors ranged from 4.26% to 4.70%, while errors at 20 microvolts ranged from 28.72% to 40.57% (Supplementary Figure S2). Agreement under smoothing therefore coexisted with distortion of the known target.

### 3.4. Coordinate dependence and boundary sensitivity

Classical curvature and torsion were numerically preserved by the proper rotation and same-curve reparametrization. Almost-curvature changed by −85.00% (95% CI −85.51 to −84.56) in LUDB and −84.09% (95% CI −84.44 to −83.56) externally under reparametrization. External Spearman correlation for this index was 0.302. Reference 1 explicitly describes the derivative-order reduction used to define equation 6; the source does not support interpreting it as a typographical error. Let *C* = *x*^′^*y*^′′^ − *y*^′^*x*^′′^ and *G* = (1 + *x*^′2^ + *y*^′2^)^3/2^. With *z* = *t*, the printed index and its same-curve reparametrization *t* = *bu* are

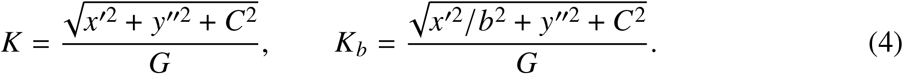

Consequently, *K*_*b*_ tends to 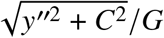, not classical curvature, whose numerator also contains *x*^′′2^. The slope proxy |*x*^′^|/*G* differed from *K* by a median −3.17% in LUDB and −3.78% in PTB-XL, with correlations 0.997 and 0.994. The limiting expression differed from classical curvature by −33.23% and −30.70%. These directly computed quantities replace an inference about term dominance from an aggregate percentage change. The pointwise term fraction, ratio of integrated descriptors and ratio of participant medians use different weighting and nonlinear aggregation, so they need not imply the same effective fraction. The limiting-index/classical-index comparison above is distinct from the limiting-index/original-index ratio: the latter is 0.149961 in LUDB and 0.159050 in PTB-XL, directly explaining approximately −85.00% and −84.09% reparametrization changes (Table S10b). The supplementary derivation includes a straight-line rotation control and a counterexample to the proposed classical-curvature limit.

Changing the time-axis scale to *t*/(100 ms), which changes the geometry, yielded absolute-torsion rank correlations of 0.638 in LUDB and 0.715 in PTB-XL. External median maximum absolute changes after the four QRS-boundary perturbations were 8.14% for FD and 7.18% for SPEC. Changing SPEC extension from reflection to periodicity gave an external median absolute change of 4.21%. Full coordinate-audit results are in Table S2; boundary and extension results are in Table S3.

### 3.5. Window-duration sensitivity

Eligible-window durations were 102 [94–114] ms in LUDB and 150 [138–166] ms in PTB-XL (median [interquartile range]); the 5th–95th percentiles were 82–150 and 116–192 ms. The 80–160-ms restriction retained 196 LUDB participants with 1,652 windows and 930 PTB-XL participants with 6,668 windows. External FD and SPEC changes from 150 to 40 Hz remained −52.53% and −72.50%, with correlations 0.103 and 0.084. The external SPEC–FD difference at 150 Hz was 79.84% (95% CI 77.28–81.38). Complete restricted comparisons are supplied in Table S9. Persistence in this restricted sample reduces concern that the original contrast was solely due to outlying durations; it does not isolate duration as a cause or equate delineation methods.

Although eligible PTB-XL windows had smaller median V1–V6 peak separation than LUDB (6 versus 24 ms), their annotated single-lead durations were already longer: medians were 140 versus 98 ms in V1 and 114 versus 90 ms in V6. The median amount added by taking the union beyond the longer single-lead interval was 6 versus 0 ms. Thus, peak separation alone does not explain the 150-versus-102-ms union-duration difference. These observations support sensitivity to annotation provenance but do not isolate the effect of ECGDeli from cohort differences (Table S12c). In a post hoc count sensitivity check, lowering the fixed matching tolerance from 100 to 80 ms retained 9,724 of 9,735 external windows and 1,729 of 1,748 LUDB windows, with one participant losing all windows in each cohort. This check concerns selection counts; descriptor contrasts were not recomputed at 80 ms (Table S12b).

## 4. Discussion

This benchmark identifies three different requirements for interpretable time-embedded 3D ECG measurements. The curve must be specified through its coordinates and scales; the numerical derivatives must be calibrated against an appropriate target; and robustness must be assessed under plausible processing and boundary changes. External replication shows that the observed operator and filter dependence is not confined to the small development dataset. It does not turn any operator into a reference measurement of the underlying cardiac electrical process. The transfer-function analysis identifies a concrete explanation for a large part of the native SPEC–FD difference.

The harmonic formula is exact under its stated assumptions, and the 20-Hz comparison empirically tests the contribution of bandwidth. It does not authorize division of every ECG descriptor by the sinc factor at its nominal cutoff. Nonlinear ratios, transient content, cancellations and noise prevent that inference.

The spectral implementation addresses a precise numerical issue: all derivative orders are taken from one explicitly defined interpolant. Its excellent noiseless calibration in these controls is consistent with spectral approximation principles [11]. The noisy controls demonstrate why that property is insufficient. Exact differentiation of a noisy reconstruction can accurately reproduce the noise and consequently yield an inaccurate descriptor of the intended noiseless curve. The tested Tikhonov baseline makes the benefit and cost of regularization explicit: noise reduction in these controls accompanied by nonzero noiseless bias. This is consistent with the established need for regularization [8, 9], while the SG findings illustrate why different derivative orders cannot be treated as a single common smoothing response [10]. These are adaptations and tests of existing numerical principles, not a claim that Fourier differentiation or the bias-noise tradeoff is new.

A practical improvement is therefore to accompany a 3D ECG descriptor with a reproducible measurement specification and a calibration profile. At minimum, that specification should state lead order, physical units, voltage/time scales, bandwidth, derivative operator and settings, QRS boundaries, aggregation and invalid-value handling. An apparent benefit from a different smoother should be assessed for both noise suppression and loss of relevant signal content. Measurements from incompatible specifications should not be pooled merely because they share a descriptor name. The present results do not justify a single clinical reliability threshold or a universally recommended cutoff.

The almost-curvature audit clarifies the behavior of an intentionally modified empirical index. Under the fixed millivolt/millisecond convention, it closely approximates a normalized V1 slope, while its parameter dependence is algebraically distinct from classical curvature. This does not disprove its predictive usefulness, but any such usefulness must be tested against the simpler slope proxy and classical curvature with identical signals, windows and validation. We evaluated equation 6 as printed; other variants and all original software implementations were not audited. This study cannot establish or retract the clinical claims in the earlier publications.

For a curve with coordinates (*x*(*t*), *y*(*t*), *t*), classical torsion can be expressed as the angular velocity of the two-dimensional second-derivative vector multiplied by a scale-dependent correction. This identity helps explain why high-order signal structure matters, without establishing an anatomical twisting mechanism. A deterministic transformation of the same voltages may help feature extraction or learning with finite samples, but does not add independent physiological observations. Future diagnostic studies should compare models receiving the same underlying leads and windows and should not infer clinical sensitivity from percentage changes in a descriptor [1–4].

### 4.1. Limitations

The development dataset had been explored before the final protocol. External selection was fixed in advance of descriptor calculations, but represented a computational sample from one PTB-XL fold rather than an unselected clinical population. Automatic external delineation and manually annotated development windows differ in provenance. Acquisition transfer functions and true noise levels were not estimated; post-acquisition filtering cannot recover removed information. Between-cohort differences cannot be attributed to physiology alone. Analyses were not stratified by recorded sex; generalizability across sex strata remains unassessed.

The analytic controls provide exact values but cover only four smooth trajectories, one Gaussian transient and additive Gaussian noise at two nonzero levels. They do not reproduce the full range of electrode artifact, baseline wander, muscle activity, clipping, pacing or complex QRS morphology. Only V1/V6 QRS geometry was studied, so the results are not a replication of all polar representations, J-wave analyses, signed-torsion summaries or ischemia-classification windows in earlier work. The added regularizer, bandwidth criterion, transient and duration restriction were post hoc. Matching a single response crossing does not equalize full bandwidth, signal fidelity or curve coherence, and 20 Hz is not a recommended clinical cutoff. TIK was not optimized, and no total-variation, data-adaptive parameter selection or exhaustive method comparison was performed. No clinical outcomes, prognostic endpoints, diagnostic thresholds or external truth for ECG torsion were evaluated. Computational cross-checks reduce implementation risk but do not substitute for independent scientific review.

## 5. Conclusions

Transfer functions explain an important, testable component of processing sensitivity in time-embedded 3D ECG descriptors. Matching a defined derivative-response crossing greatly reduced SPEC–FD differences but produced 4.26–4.70% noiseless error on the Gaussian transient; numerical agreement did not establish accuracy. Analytic controls exposed filtering bias, cancellation within an aggregated descriptor, and the benefits and costs of regularization. Almost-curvature behaved as a parameter-dependent index close to a normalized voltage slope under the specified convention. Absolute levels, inclusion counts and complete calibration outputs provide reproducible measurement specifications. These results establish neither a biological ground truth nor diagnostic superiority.

## Supporting information

Research materials and reproducible analysis code

## Data Availability

The source ECG datasets and annotations are openly available from PhysioNet: LUDB version 1.0.1 (https://physionet.org/content/ludb/1.0.1/), PTB-XL version 1.0.3 (https://physionet.org/content/ptb-xl/1.0.3/), and PTB-XL+ version 1.0.1 (https://physionet.org/content/ptb-xl-plus/1.0.1/). The accompanying Research_materials.zip contains analysis code, selection manifests, source checksums, derived per-window and participant-level results, and calibration outputs. Supplementary_tables_and_figure_data.zip contains supplementary tables and numerical source data for the figures. Original ECG recordings are not redistributed in these archives; the included download scripts retrieve them from the cited repositories. No separate repository DOI has yet been assigned to the derived research materials.

## 6. Data availability and research governance

Source ECGs and annotations are publicly available in LUDB 1.0.1, PTB-XL 1.0.3 and PTBXL+ 1.0.1 [13, 15, 17]. The accompanying ancillary archive (Research_materials.zip) contains the selection manifest, source hashes, protocol, annotation amendment, analysis code, per-window results and participant summaries. The original public ECG recordings and annotations are retrieved from their cited repositories by the supplied download scripts. Source data for all figures and Supplementary Tables S1–S13 are supplied as separate files. The added analyses, locally hashed revision plan and independent computational checks are included in the same archive. A permanent repository identifier for the derived research materials has not yet been issued. No new recruitment, intervention, contact or reidentification was performed. Source-dataset governance is described in the original publications; this manuscript does not assert a new institutional ethics approval or exemption.

## 9. CRediT authorship contribution statement

Alejandro Jesús Bermejo Valdés: Conceptualization, Methodology, Formal analysis, Validation, Investigation, Software, Data curation, Visualization, Writing – original draft, Writing – review and editing.

