## Supplementary material for "Calibration and processing sensitivity of time-embedded three-dimensional electrocardiographic descriptors: a two-dataset benchmark": Research materials and reproducible analysis code: integrated_analysis.py

```
"""Exploratory extension. Real LUDB records and analytic simulations kept separate."""
from pathlib import Path
from datetime import datetime,timezone
import sys,json,hashlib
import numpy as np
import pandas as pd
from scipy.signal import savgol_filter
from scipy.stats import spearmanr

ROOT=Path(__file__).resolve().parent
PHASE1=ROOT.parent/'estudio_3decg_reproducibilidad'
sys.path.insert(0,str(PHASE1))
import analyze as old
OUT=ROOT/'results';OUT.mkdir(exist_ok=True)
MEASURES=['curvature_mean','abs_torsion_mean','almost_curvature_mean','weighted_abs_torsion']
METHODS=['fd','sg','sg0fd']
CUTOFFS=[20,30,40,50,100,150]

def sha(p):return hashlib.sha256(p.read_bytes()).hexdigest()
def lock():
    f=ROOT/'plan_lock.json'
    record=dict(sha256=sha(ROOT/'PLAN_FASE2.md'),utc=datetime.now(timezone.utc).isoformat())
    if not f.exists():f.write_text(json.dumps(record,indent=2))
    assert json.loads(f.read_text())['sha256']==record['sha256']

def derive(xy,method):
    if method=='sg0fd':
        xy=savgol_filter(xy,11,3,deriv=0,axis=0,mode='interp')
        return old.derivatives(xy,2.,'fd')
    return old.derivatives(xy,2.,method)

def geom(d,temporal_scale=1.,rotate=False,reparam=1.):
    d1,d2,d3=[a.copy() for a in d]
    if rotate:
        d1,d2,d3=[np.column_stack((-a[:,1],a[:,0])) for a in (d1,d2,d3)]
    n=len(d1)
    p1=np.column_stack((d1,np.full(n,temporal_scale)))*reparam
    p2=np.column_stack((d2,np.zeros(n)))*reparam**2
    p3=np.column_stack((d3,np.zeros(n)))*reparam**3
    cross=np.cross(p1,p2); D=np.sum(cross**2,axis=1)
    N=np.sum(cross*p3,axis=1)
    norm3=np.sum(p1**2,axis=1)**1.5
    valid=D>np.finfo(float).eps*np.max(D)
    tau=np.divide(N,D,out=np.full(n,np.nan),where=valid)
    x,y,z=p1.T; xx,yy,zz=p2.T
    almost=np.sqrt((yy*z-y*zz)**2+x*x*(zz-z)**2+(xx*y-x*yy)**2)/norm3
    # Equivalent explicit expression, independently checked.
    A=np.sum(d2*d2,axis=1)
    C=d1[:,0]*d2[:,1]-d1[:,1]*d2[:,0]
    B=d2[:,0]*d3[:,1]-d2[:,1]*d3[:,0]
    den2=temporal_scale**2*A+C*C
    tau2=np.divide(temporal_scale*B,den2,out=np.full(n,np.nan),where=valid)
    np.testing.assert_allclose(tau[valid],tau2[valid],rtol=1e-10,atol=1e-11)
    acc_rotation=np.divide(B,A,out=np.full(n,np.nan),where=A>0)
    q=np.divide(C*C,A,out=np.zeros(n),where=A>0)
    assert np.all(q<=np.sum(d1*d1,axis=1)+1e-12)
    if temporal_scale==1:
        np.testing.assert_allclose(tau[valid],(acc_rotation/(1+q))[valid],rtol=1e-10,atol=1e-11)
    return dict(curvature=np.sqrt(D)/norm3,torsion=tau,almost=almost,
                determinant=N,denominator=D,acc_rotation=acc_rotation,q=q)

def measure(g,lo,hi,dt=2.):
    # Inclusive integer sample bounds; same pre-existing QRS window in every arm.
    idx=slice(lo,hi+1)
    T=(hi-lo)*dt
    assert hi>lo
    arrays={k:v[idx] for k,v in g.items()}
    for k,v in arrays.items():
        assert np.isfinite(v).all(),f'Undefined samples in {k}: {lo}:{hi}'
    avg=lambda v:float(np.trapezoid(v,dx=dt)/T)
    out=dict(curvature_mean=avg(arrays['curvature']),
             abs_torsion_mean=avg(abs(arrays['torsion'])),
             almost_curvature_mean=avg(arrays['almost']),
             weighted_abs_torsion=float(np.trapezoid(abs(arrays['determinant']),dx=dt)/
                                        np.trapezoid(arrays['denominator'],dx=dt)),
             abs_acc_rotation_mean=avg(abs(arrays['acc_rotation'])),
             max_q=float(arrays['q'].max()))
    return out

def analytic_checks():
    t=np.arange(2000)*2.
    omega=2*np.pi*30/1000
    phase=omega*t
    d1=np.column_stack((-omega*np.sin(phase),omega*np.cos(phase)))
    d2=np.column_stack((-omega**2*np.cos(phase),-omega**2*np.sin(phase)))
    d3=np.column_stack((omega**3*np.sin(phase),-omega**3*np.cos(phase)))
    d=(d1,d2,d3)
    g=geom(d);rot=geom(d,rotate=True);rep=geom(d,reparam=1000)
    np.testing.assert_allclose(g['torsion'],omega/(1+omega**2),atol=1e-14)
    for key in ['curvature','torsion']:
        np.testing.assert_allclose(g[key],rot[key],rtol=1e-12,atol=1e-12)
        np.testing.assert_allclose(g[key],rep[key],rtol=1e-12,atol=1e-12)
    # Proper rotation of a straight line: almost-curvature changes, curvature does not.
    p1=np.tile([1.,0.],(20,1));p2=np.zeros_like(p1)
    straight=geom((p1,p2,p2));straight_rot=geom((p1,p2,p2),rotate=True)
    assert np.all(straight['curvature']==0) and np.isnan(straight['torsion']).all()
    np.testing.assert_allclose(straight['almost'],1/(2*np.sqrt(2)))
    assert np.all(straight_rot['almost']==0)
    return dict(analytic_helix='pass',rotation_classical_geometry='pass',
                reparametrization_classical_geometry='pass',
                straight_line_almost=float(straight['almost'][0]),
                rotated_straight_line_almost=float(straight_rot['almost'][0]),
                angular_acceleration_identity='asserted on all arrays',
                cauchy_schwarz_bound='asserted on all arrays')

def ecg():
    windows=pd.read_csv(PHASE1/'results/qrs_windows.csv')
    beatrows=[];auditrows=[]
    for i,(record,ws) in enumerate(windows.groupby('record'),1):
        _,xy=old.read(record)
        for cutoff in CUTOFFS:
            signal=old.lowpass(xy,cutoff)
            for method in METHODS:
                d=derive(signal,method);g=geom(d)
                for _,w in ws.iterrows():
                    row=dict(record=record,beat=int(w.beat),cutoff=cutoff,method=method)
                    row.update(measure(g,int(w.start_sample),int(w.end_sample)))
                    beatrows.append(row)
                if cutoff==150 and method=='fd':
                    variants={'baseline':g,'rotation90':geom(d,rotate=True),
                              'parameter_seconds':geom(d,reparam=1000),
                              'time_scale_10ms':geom(d,temporal_scale=.1),
                              'time_scale_100ms':geom(d,temporal_scale=.01)}
                    for name,variant in variants.items():
                        if name in ('rotation90','parameter_seconds'):
                            for key in ('curvature','torsion'):
                                np.testing.assert_allclose(g[key],variant[key],rtol=1e-9,atol=1e-11,equal_nan=True)
                        for _,w in ws.iterrows():
                            row=dict(record=record,beat=int(w.beat),variant=name)
                            row.update(measure(variant,int(w.start_sample),int(w.end_sample)))
                            if name=='baseline':
                                relative=row['abs_acc_rotation_mean']/row['abs_torsion_mean']-1
                                assert -1e-12<=relative<=row['max_q']+1e-10
                                row['approximation_relative_error_percent']=100*relative
                            auditrows.append(row)
        if i%50==0:print(f'Real records processed: {i}',flush=True)
    beats=pd.DataFrame(beatrows);audit=pd.DataFrame(auditrows)
    beats.to_csv(OUT/'real_beat_metrics.csv',index=False)
    subjects=beats.groupby(['record','cutoff','method'])[MEASURES+['abs_acc_rotation_mean','max_q']].median().reset_index()
    subjects.to_csv(OUT/'real_subject_metrics.csv',index=False)
    audit.to_csv(OUT/'geometric_audit_beats.csv',index=False)
    a=audit.groupby(['record','variant'])[MEASURES+['abs_acc_rotation_mean','max_q','approximation_relative_error_percent']].median().reset_index()
    a.to_csv(OUT/'geometric_audit_subjects.csv',index=False)
    # Replicate phase 1 measurements before drawing any extension conclusions.
    previous=pd.read_csv(PHASE1/'results/subject_metrics.csv')
    for cutoff in [40,100,150]:
        left=subjects[(subjects.cutoff==cutoff)&(subjects.method=='fd')].set_index('record')[MEASURES[:3]].sort_index()
        right=previous[previous.condition==f'fd{cutoff}'].set_index('record')[MEASURES[:3]].sort_index()
        np.testing.assert_allclose(left,right,rtol=1e-10,atol=1e-12)
    return subjects,a

def ci(values,rng):
    x=np.asarray(values);assert np.isfinite(x).all()
    v=np.median(x[rng.integers(0,len(x),size=(5000,len(x)))],axis=1)
    return np.quantile(v,[.025,.975])

def compare(a,b,rng):
    assert len(a)==len(b)==200
    pct=100*(a-b)/b
    sym=200*abs(a-b)/(abs(a)+abs(b))
    lo,hi=ci(pct,rng)
    return dict(n=len(a),change_median=float(np.median(pct)),
                q25=float(np.quantile(pct,.25)),q75=float(np.quantile(pct,.75)),
                ci025=float(lo),ci975=float(hi),
                absolute_change_median=float(np.median(abs(pct))),
                symmetric_difference_median=float(np.median(sym)),
                spearman=float(spearmanr(a,b).statistic))

def summarize(subjects,audit):
    rng=np.random.default_rng(20260915);out=[]
    for method in METHODS:
        for metric in MEASURES:
            matrix=subjects[subjects.method==method].pivot(index='record',columns='cutoff',values=metric)
            for cutoff in CUTOFFS[:-1]:
                row=dict(method=method,metric=metric,cutoff=cutoff,reference=150)
                row.update(compare(matrix[cutoff].to_numpy(),matrix[150].to_numpy(),rng));out.append(row)
    pd.DataFrame(out).to_csv(OUT/'real_comparisons.csv',index=False)
    out=[]
    for metric in MEASURES:
        matrix=audit.pivot(index='record',columns='variant',values=metric)
        for variant in ['rotation90','parameter_seconds','time_scale_10ms','time_scale_100ms']:
            row=dict(metric=metric,variant=variant)
            row.update(compare(matrix[variant].to_numpy(),matrix.baseline.to_numpy(),rng));out.append(row)
    pd.DataFrame(out).to_csv(OUT/'geometry_comparisons.csv',index=False)

def truth_signal(kind,t):
    if kind.startswith('helix_'):
        freq=float(kind.split('_')[1]);phase=2*np.pi*freq*t/1000
        f1=np.full(len(t),2*np.pi*freq/1000);f2=np.zeros(len(t));f3=f2.copy()
    else:
        omega=2*np.pi*20/1000;nu=2*np.pi*3/1000
        phase=omega*t+.8*np.sin(nu*t)
        f1=omega+.8*nu*np.cos(nu*t)
        f2=-.8*nu**2*np.sin(nu*t);f3=-.8*nu**3*np.cos(nu*t)
    sine,cosine=np.sin(phase),np.cos(phase)
    xy=np.column_stack((cosine,sine))
    d1=np.column_stack((-sine*f1,cosine*f1))
    d2=np.column_stack((-cosine*f1**2-sine*f2,-sine*f1**2+cosine*f2))
    d3=np.column_stack((sine*f1**3-3*cosine*f1*f2-sine*f3,
                       -cosine*f1**3-3*sine*f1*f2+cosine*f3))
    return xy,(d1,d2,d3)

def simulations():
    rng=np.random.default_rng(20260916);t=np.arange(2000)*2.
    rows=[];truthrows=[]
    for kind in ['helix_10','helix_30','helix_60','modulated_phase']:
        xy,derivs=truth_signal(kind,t)
        true=measure(geom(derivs),500,1500)
        truthrows.append(dict(model=kind,**true))
        for noise in [0.,.005,.02]:
            count=1 if noise==0 else 32
            for realization in range(count):
                noisy=xy+rng.normal(0,noise,xy.shape)
                for cutoff in [40,150]:
                    filtered=old.lowpass(noisy,cutoff)
                    for method in METHODS:
                        estimates=measure(geom(derive(filtered,method)),500,1500)
                        for metric in ['abs_torsion_mean','weighted_abs_torsion']:
                            rows.append(dict(model=kind,noise_sd_mV=noise,realization=realization,
                               cutoff=cutoff,method=method,metric=metric,truth=true[metric],
                               estimate=estimates[metric],relative_error_percent=100*(estimates[metric]/true[metric]-1)))
    frame=pd.DataFrame(rows)
    frame.to_csv(OUT/'simulation_estimates.csv',index=False)
    pd.DataFrame(truthrows).to_csv(OUT/'simulation_truth.csv',index=False)
    summary=[]
    for key,part in frame.groupby(['model','noise_sd_mV','cutoff','method','metric']):
        err=part.relative_error_percent.to_numpy()
        row=dict(zip(['model','noise_sd_mV','cutoff','method','metric'],key))
        row.update(n=len(part),relative_bias_percent=float(np.mean(err)),
                   relative_rmse_percent=float(np.sqrt(np.mean(err**2))))
        summary.append(row)
    pd.DataFrame(summary).to_csv(OUT/'simulation_summary.csv',index=False)
    print('Separate analytic controls processed: 4 models, 260 signal/noise realizations.',flush=True)

def main():
    lock();checks=analytic_checks();s,a=ecg();summarize(s,a);simulations()
    approx=a[a.variant=='baseline'].approximation_relative_error_percent
    summary=dict(real_subjects=200,real_QRS_windows=1748,factorial_conditions=18,
                 comparisons=60,geometric_variants=5,
                 analytic_signal_noise_realizations=260,simulated_patients=0,
                 approximation_error_percent_median=float(approx.median()),
                 approximation_error_percent_q25=float(approx.quantile(.25)),
                 approximation_error_percent_q75=float(approx.quantile(.75)),
                 checks=checks,phase1_reproduction='passed for all 3 shared filters and 3 metrics',
                 phase1_window_sha256=sha(PHASE1/'results/qrs_windows.csv'),
                 script_sha256=sha(Path(__file__)),plan_sha256=sha(ROOT/'PLAN_FASE2.md'),
                 utc=datetime.now(timezone.utc).isoformat())
    (OUT/'summary.json').write_text(json.dumps(summary,indent=2))
    print(json.dumps(summary,indent=2))

if __name__=='__main__':main()
```
