## Supplementary material for "Calibration and processing sensitivity of time-embedded three-dimensional electrocardiographic descriptors: a two-dataset benchmark": Research materials and reproducible analysis code: PLAN_FASE2.md

# Plan de integración y evaluación de soluciones

Fijado antes de calcular los resultados de esta fase. Es una ampliación exploratoria motivada por los resultados previos de LUDB, no un estudio independiente ni un prerregistro público. Se mantienen los 200 registros y las mismas 1.748 ventanas QRS para aislar los factores de procesamiento.

## Preguntas

1. ¿Cuánto depende almost-curvature de rotar los ejes o de cambiar el parámetro de derivación, conservando la misma curva geométrica?
2. ¿Qué relación exacta tiene la torsión de (V1,V6,tiempo) con la velocidad angular del vector de segundas derivadas?
3. ¿Una ponderación que evita dividir por denominadores locales pequeños reduce la dependencia del procesamiento? Es un descriptor distinto y debe evaluarse con su propio valor de referencia.
4. ¿La menor dependencia entre filtros predice menor error cuando la geometría verdadera sí es conocida?

## Análisis en ECG reales

- Misma decodificación y mismas ventanas que en fase 1; sin nuevas exclusiones por diagnóstico, edad o resultado.
- Diseño cruzado: seis filtros nominales (20, 30, 40, 50, 100, 150 Hz) y tres métodos de derivación: diferencias finitas; Savitzky-Golay directo, grado 3 y 11 muestras; Savitzky-Golay de orden de derivación cero seguido de diferencias finitas. La última condición ayuda a examinar el efecto del suavizado previo, pero no garantiza igualdad exacta de las respuestas de los operadores.
- Medidas: curvatura clásica media, torsión absoluta media, almost-curvature media y torsión absoluta ponderada por D=||r'×r''||². Esta última es integral(|det(r',r'',r''')|)/integral(D), no una estimación equivalente a la media temporal de |torsión| cuando la torsión varía.
- Comparaciones de todos los filtros frente a 150 Hz dentro de cada estimador. Agregar primero por la mediana de latidos de cada persona. Reportar diferencias porcentuales, diferencia relativa simétrica y Spearman; intervalos bootstrap descriptivos por persona, 5.000 réplicas, semilla 20260915. No escoger un ganador por valor p.
- Auditoría a 150 Hz y diferencias finitas: rotación propia de 90 grados en el plano de voltajes; reparametrización t_ms=1000*u sin cambiar coordenadas; escalas geométricas temporales t/1 ms, t/10 ms y t/100 ms. Las dos primeras son pruebas de invariancia sobre la misma curva. La última cambia deliberadamente la geometría y no es una conversión equivalente de unidades.
- Comparar la torsión con la velocidad angular del vector (x'',y'') y cuantificar la corrección geométrica exacta. Con A=x''²+y''², C=x'y''-y'x'', N=x''y'''-y''x''', se tiene τ=(N/A)/(1+C²/A), si A>0 y z'=1. Además, C²/A <= x'²+y'².

## Controles matemáticos separados de los pacientes

- Hélices analíticas con x=cos(φ), y=sin(φ), z=t_ms y φ=2π*f*t_ms/1000, para f=10,30,60 Hz.
- Una hélice de fase modulada: φ=2π*20*t_ms/1000 + 0,8*sin(2π*3*t_ms/1000). Su torsión varía; cada descriptor tiene su propio valor analítico de referencia.
- Señales de 4 s a 500 Hz; evaluar entre 1 y 3 s para limitar bordes. Añadir ruido gaussiano independiente a los dos voltajes con desviación 0, 0,005 o 0,020 mV. Son niveles de prueba, no una estimación del ruido de LUDB ni simulaciones fisiológicas de pacientes.
- Una realización sin ruido y 32 realizaciones por nivel con ruido; semilla 20260916. La misma realización se procesa con los filtros de 40 y 150 Hz y los tres estimadores.
- Comparar la media temporal de torsión absoluta y su versión ponderada contra sus respectivos valores verdaderos: sesgo relativo y raíz del error cuadrático medio relativo. Conservar todos los escenarios. Estos controles no acreditan fidelidad clínica de un método.

## Rigor e interpretación

- No llamar «solución» a un método solo por cambiar menos entre filtros.
- Separar: corrección matemática; estabilidad computacional; preservación de información; utilidad diagnóstica. Cada nivel necesita evidencia distinta.
- La rotación de ejes no representa una nueva derivación medida ni una recolocación real de electrodos.
- Ninguna cifra de las simulaciones se sumará a los 200 sujetos reales.
- Conservar resultados completos, fórmulas, identidades verificadas, versiones y huellas de los archivos. Mantener intacta la fase 1.
