## Supplementary material for "Calibration and processing sensitivity of time-embedded three-dimensional electrocardiographic descriptors: a two-dataset benchmark": Research materials and reproducible analysis code: analysis.py

```
"""Locked multicohort measurement benchmark; no diagnostic modeling."""
from pathlib import Path
from datetime import datetime,timezone
from functools import lru_cache
import sys,json,hashlib,argparse
import numpy as np
import pandas as pd
import wfdb
from scipy.signal import butter,sosfiltfilt,savgol_filter,sosfreqz
from scipy.stats import spearmanr

ROOT=Path(__file__).resolve().parent;OUT=ROOT/'results';OUT.mkdir(exist_ok=True)
P1=ROOT.parent/'estudio_3decg_reproducibilidad';P2=ROOT.parent/'estudio_3decg_integrado_fase2'
sys.path.insert(0,str(P1));import analyze as prior
sys.path.insert(0,str(P2));from integrated_analysis import truth_signal
METHODS=['FD','SG','SPEC','SG0SPEC'];CUTS=[40,100,150]
METRICS=['abs_torsion','curvature','almost_curvature','weighted_abs_torsion']

def sha(p):return hashlib.sha256(p.read_bytes()).hexdigest()
def lock(task):
    assert sha(ROOT/'PROTOCOL.md')==json.loads((ROOT/'protocol_lock.json').read_text())['sha256']
    stamp={'script_sha256':sha(Path(__file__)),'protocol_sha256':sha(ROOT/'PROTOCOL.md'),
           'selection_sha256':sha(ROOT/'data/external_selection.csv'),'utc':datetime.now(timezone.utc).isoformat()}
    stamp['annotation_amendment_sha256']=sha(ROOT/'ANNOTATION_AMENDMENT.md')
    (OUT/f'{task}_provenance.json').write_text(json.dumps(stamp,indent=2))

@lru_cache(None)
def angular_frequencies(n):return 2*np.pi*np.fft.rfftfreq(n,d=2.)

def spectral(xy,periodic=False):
    ext=xy if periodic else np.concatenate((xy,xy[-2:0:-1]),axis=0)
    n=len(ext);w=angular_frequencies(n);ft=np.fft.rfft(ext,axis=0)
    if n%2==0:ft[-1]=0 # one common interpolant, including the second derivative
    return tuple(np.fft.irfft(ft*(1j*w[:,None])**k,n=n,axis=0)[:len(xy)] for k in [1,2,3])

def derive(xy,method,periodic=False):
    if method=='FD':return prior.derivatives(xy,2.,'fd')
    if method=='SG':return prior.derivatives(xy,2.,'sg')
    if method=='SG0SPEC':xy=savgol_filter(xy,11,3,axis=0,mode='interp')
    return spectral(xy,periodic=periodic)

def geometry(d,rotation=False,reparam=1.,scale=1.):
    p,q,s=[x.copy() for x in d]
    if rotation:p,q,s=[np.column_stack((-x[:,1],x[:,0])) for x in (p,q,s)]
    p=np.column_stack((p,np.full(len(p),scale)))*reparam
    q=np.column_stack((q,np.zeros(len(q))))*reparam**2
    s=np.column_stack((s,np.zeros(len(s))))*reparam**3
    cross=np.cross(p,q);D=np.sum(cross*cross,axis=1);N=np.sum(cross*s,axis=1)
    speed3=np.sum(p*p,axis=1)**1.5
    valid=D>np.finfo(float).eps*D.max()
    tau=np.divide(N,D,out=np.full(len(D),np.nan),where=valid)
    x,y,z=p.T;xx,yy,zz=q.T
    almost=np.sqrt((yy*z-y*zz)**2+x*x*(zz-z)**2+(xx*y-x*yy)**2)/speed3
    return {'abs_torsion':abs(tau),'curvature':np.sqrt(D)/speed3,'almost_curvature':almost,'D':D,'N':N}

def measure(g,lo,hi):
    assert 0<=lo<hi<len(g['D']);idx=slice(lo,hi+1);T=(hi-lo)*2.
    def avg(v):return float(np.trapezoid(v[idx],dx=2.)/T) if np.isfinite(v[idx]).all() else np.nan
    out={m:avg(g[m]) for m in METRICS[:3]}
    d=float(np.trapezoid(g['D'][idx],dx=2.))
    out['weighted_abs_torsion']=float(np.trapezoid(abs(g['N'][idx]),dx=2.)/d) if d>0 else np.nan
    ok=np.isfinite(g['abs_torsion'][idx]);out['valid_duration_fraction']=float(np.mean(ok[:-1]&ok[1:]))
    return out

def ann_points(path):
    a=wfdb.rdann(str(path),'atr');triples=[];incomplete=0
    if a.fs!=500:raise ValueError('Annotation sampling frequency is not 500 Hz')
    labels=np.asarray(a.aux_note);samples=np.asarray(a.sample)
    peaks=samples[labels=='R peak'];onsets=samples[labels=='QRS onset'];offsets=samples[labels=='QRS offset']
    for j,pk in enumerate(peaks):
        prev=peaks[j-1] if j else -np.inf;nxt=peaks[j+1] if j+1<len(peaks) else np.inf
        starts=onsets[(onsets>prev)&(onsets<=pk)];ends=offsets[(offsets>=pk)&(offsets<nxt)]
        if len(starts)!=1 or len(ends)!=1:incomplete+=1;continue
        triples.append((int(starts[0]),int(pk),int(ends[0])))
    return np.asarray(triples,dtype=int).reshape(-1,3),incomplete

def external_windows(row):
    folder=Path(row.filename_hr).parent.name
    (a,ia),(b,ib)=[ann_points(ROOT/'data/plus/fiducial_points/ecgdeli'/folder/f'{row.ecg_id:05}_points_lead_{lead}') for lead in ['V1','V6']]
    audit={'record':str(row.ecg_id),'patient':int(row.patient_id),'qrs_v1':len(a),'qrs_v6':len(b),
           'incomplete_v1':ia,'incomplete_v6':ib,'matched':0,'excluded_duration':0,'excluded_edge':0,'eligible':0}
    ws=[]
    if len(a) and len(b):
        dist=abs(a[:,1,None]-b[None,:,1])
        for i,j in enumerate(dist.argmin(axis=1)):
            if dist[i,j]>50 or dist[:,j].argmin()!=i:continue
            audit['matched']+=1;lo=min(a[i,0],b[j,0]);hi=max(a[i,2],b[j,2])
            if not 20<=hi-lo<=150:audit['excluded_duration']+=1;continue
            if lo<500 or hi>4499:audit['excluded_edge']+=1;continue
            ws.append({'beat':len(ws)+1,'start_sample':int(lo),'end_sample':int(hi),
                       'v1_peak':int(a[i,1]),'v6_peak':int(b[j,1])})
    audit['eligible']=len(ws);audit['unmatched_v1']=len(a)-audit['matched'];audit['unmatched_v6']=len(b)-audit['matched']
    return ws,audit

def load_external(row):
    r=wfdb.rdrecord(str(ROOT/'data/ptbxl'/row.filename_hr))
    if r.fs!=500 or r.sig_len!=5000 or r.n_sig!=12:raise ValueError('Unexpected acquisition dimensions')
    names=[s.upper() for s in r.sig_name];ch=[names.index(s) for s in ['V1','V6']]
    if any(r.units[i]!='mV' for i in ch):raise ValueError('Unexpected physical units')
    xy=r.p_signal[:,ch]
    if not np.isfinite(xy).all():raise ValueError('Nonfinite voltage')
    return xy-np.median(xy,axis=0)

def cohort(name):
    lock(name);rows=[];audit=[];winrows=[];georows=[];shifts=[];extensions=[]
    if name=='LUDB':
        windows=pd.read_csv(P1/'results/qrs_windows.csv')
        items=list(windows.groupby('record'))
    else:items=list(pd.read_csv(ROOT/'data/external_selection.csv').itertuples(index=False))
    for count,item in enumerate(items,1):
        if name=='LUDB':
            record,wsdf=item;patient=int(record.split('/')[-1]);ws=wsdf.to_dict('records');_,xy=prior.read(record)
            aud={'record':record,'patient':patient,'eligible':len(ws),'status':'included'}
        else:
            record=str(item.ecg_id);patient=int(item.patient_id)
            try:
                xy=load_external(item);ws,aud=external_windows(item);aud['status']='included' if ws else 'no_eligible_QRS'
            except (FileNotFoundError,ValueError) as e:
                audit.append({'record':record,'patient':patient,'status':'excluded','reason':str(e),'eligible':0});continue
        audit.append(aud)
        if not ws:continue
        for w in ws:winrows.append(dict(cohort=name,record=record,patient=patient,**{k:w[k] for k in ['beat','start_sample','end_sample','v1_peak','v6_peak']}))
        for cut in CUTS:
            filtered=prior.lowpass(xy,cut)
            for method in METHODS:
                d=derive(filtered,method);g=geometry(d)
                for w in ws:
                    rows.append(dict(cohort=name,record=record,patient=patient,beat=w['beat'],cutoff=cut,method=method,
                                     **measure(g,w['start_sample'],w['end_sample'])))
                if cut==150 and method in ['FD','SPEC']:
                    for ds,de in [(-5,-5),(-5,5),(5,-5),(5,5)]:
                        for w in ws:
                            lo=w['start_sample']+ds;hi=w['end_sample']+de
                            if 0<=lo<hi<len(xy):
                                shifts.append(dict(cohort=name,patient=patient,record=record,beat=w['beat'],method=method,
                                                   onset_shift_ms=ds*2,offset_shift_ms=de*2,**measure(g,lo,hi)))
                if cut==150 and method=='SPEC':
                    gp=geometry(derive(filtered,'SPEC',periodic=True))
                    for w in ws:extensions.append(dict(cohort=name,patient=patient,record=record,beat=w['beat'],
                                                      **measure(gp,w['start_sample'],w['end_sample'])))
                if cut==150 and method=='FD':
                    for variant,gv in [('baseline',g),('rotation90',geometry(d,rotation=True)),
                                       ('parameter_seconds',geometry(d,reparam=1000)),('time_scale100',geometry(d,scale=.01))]:
                        if variant in ['rotation90','parameter_seconds']:
                            for m in ['abs_torsion','curvature']:np.testing.assert_allclose(g[m],gv[m],rtol=1e-9,atol=1e-11,equal_nan=True)
                        for w in ws:georows.append(dict(cohort=name,record=record,patient=patient,beat=w['beat'],variant=variant,
                                                       **measure(gv,w['start_sample'],w['end_sample'])))
        if count%100==0:print(name,'processed',count,'of',len(items),flush=True)
    for stem,values in [('beats',rows),('audit',audit),('windows',winrows),('geometry_beats',georows),('boundary_beats',shifts),('extension_beats',extensions)]:
        pd.DataFrame(values).to_csv(OUT/f'{name}_{stem}.csv',index=False)
    beats=pd.DataFrame(rows)
    summary=beats.groupby(['cohort','patient','record','cutoff','method'])[METRICS+['valid_duration_fraction']].median().reset_index()
    summary.to_csv(OUT/f'{name}_participants.csv',index=False)
    print(name,'participants',summary.patient.nunique(),'unique QRS',len(winrows),'measurement rows',len(rows),flush=True)


def calibration():
    lock('calibration');rng=np.random.default_rng(20260917);t=np.arange(4000)*2.;rows=[];targets=[];checks=[]
    for kind in ['helix_10','helix_30','helix_60','modulated_phase']:
        xy,d=truth_signal(kind,t);truth=measure(geometry(d),1000,3000)
        for cut in CUTS:
            filtered_target={m:np.nan for m in METRICS}
            if kind.startswith('helix'):
                f=float(kind.split('_')[1]);_,h=sosfreqz(butter(4,cut,fs=500,output='sos'),worN=[f],fs=500)
                amplitude=float(abs(h[0])**2)
                filtered_target=measure(geometry(tuple(v*amplitude for v in d)),1000,3000)
                # Periodic analytic signal with exact response, isolated from endpoint padding.
                exact=geometry(spectral(xy*amplitude,periodic=True));observed=measure(exact,1000,3000)
                for m in METRICS:np.testing.assert_allclose(observed[m],filtered_target[m],rtol=1e-8,atol=1e-10)
                checks.append({'model':kind,'cutoff':cut,'max_absolute_error':max(abs(observed[m]-filtered_target[m]) for m in METRICS)})
            for m in METRICS:targets.append(dict(model=kind,cutoff=cut,metric=m,unfiltered_truth=truth[m],filtered_truth=filtered_target[m]))
        for noise in [0.,.005,.02]:
            for realization in range(1 if noise==0 else 100):
                noisy=xy+rng.normal(0,noise,xy.shape)
                for cut in CUTS:
                    filtered=prior.lowpass(noisy,cut)
                    for method in METHODS:
                        val=measure(geometry(derive(filtered,method)),1000,3000)
                        for m in METRICS:rows.append(dict(model=kind,noise_sd_mV=noise,realization=realization,
                                                         cutoff=cut,method=method,metric=m,estimate=val[m],truth=truth[m]))
        print('Calibration',kind,'complete',flush=True)
    frame=pd.DataFrame(rows);truths=pd.DataFrame(targets)
    frame=frame.merge(truths[['model','cutoff','metric','filtered_truth']],on=['model','cutoff','metric'],validate='many_to_one')
    frame.to_csv(OUT/'calibration_estimates.csv',index=False);truths.to_csv(OUT/'calibration_targets.csv',index=False)
    out=[]
    for keys,a in frame.groupby(['model','noise_sd_mV','cutoff','method','metric']):
        r=dict(zip(['model','noise_sd_mV','cutoff','method','metric'],keys));e=(a.estimate-a.truth)/a.truth
        r.update(n=len(a),relative_bias_percent=100*e.mean(),relative_rmse_percent=100*np.sqrt(np.mean(e*e)),nonfinite=int((~np.isfinite(e)).sum()))
        if a.filtered_truth.notna().all():
            f=(a.estimate-a.filtered_truth)/a.filtered_truth
            r.update(filtered_relative_bias_percent=100*f.mean(),filtered_relative_rmse_percent=100*np.sqrt(np.mean(f*f)))
        out.append(r)
    pd.DataFrame(out).to_csv(OUT/'calibration_summary.csv',index=False)
    (OUT/'periodic_spectral_checks.json').write_text(json.dumps(checks,indent=2))

if __name__=='__main__':
    ap=argparse.ArgumentParser();ap.add_argument('task',choices=['LUDB','PTBXL','calibration']);a=ap.parse_args()
    if a.task=='calibration':calibration()
    else:cohort(a.task)
```
