## Supplementary material for "Calibration and processing sensitivity of time-embedded three-dimensional electrocardiographic descriptors: a two-dataset benchmark": Research materials and reproducible analysis code: ANNOTATION_AMENDMENT.md

# Annotation reader specification

14 September 2026, before external descriptor calculations. Structural inspection used PTB-XL record 00001, which is outside metadata fold 10 and is not included in the external sample.

ECGDeli annotations store an explicit 500 Hz annotation sampling frequency. Their WFDB symbol is a comment marker, and event identity is encoded in `aux_note`: `QRS onset`, `R peak`, and `QRS offset`. They do not use LUDB's parenthesis/N/parenthesis convention. Other wave landmarks can occur between QRS landmarks.

For each annotated R peak, require exactly one QRS onset between the preceding R peak and the current R peak, and exactly one QRS offset between the current R peak and the next R peak. At record ends, use the open-ended available interval. Missing or ambiguous triples are counted as incomplete. Require onset <= R peak <= offset. Apply the locked V1/V6 matching, duration and edge criteria afterward. No signal-derived redetection, manual correction, diagnostic selection or descriptor-based exclusion is introduced. The source annotation indices are used directly, without shifting them, after checking their declared sampling frequency and waveform alignment.

The original protocol, sample selection and all estimator settings remain unchanged.
