## Supplementary material for "Calibration and processing sensitivity of time-embedded three-dimensional electrocardiographic descriptors: a two-dataset benchmark": Research materials and reproducible analysis code: download_external.py

```
"""Download only the locked external selection and a reader-calibration record."""
from pathlib import Path
from urllib.request import urlopen,Request
from urllib.error import HTTPError
from concurrent.futures import ThreadPoolExecutor,as_completed
from datetime import datetime,timezone
import hashlib,json,time
import pandas as pd

ROOT=Path(__file__).resolve().parent;DATA=ROOT/'data'
BASE={'ptbxl':'https://physionet.org/files/ptb-xl/1.0.3/',
      'plus':'https://physionet.org/files/ptb-xl-plus/1.0.1/'}
def digest(p):return hashlib.sha256(p.read_bytes()).hexdigest()
def fetch(kind,rel,expected=None):
    path=DATA/kind/rel;path.parent.mkdir(parents=True,exist_ok=True)
    url=BASE[kind]+rel
    for attempt in range(3):
        try:
            if not path.exists():
                req=Request(url,headers={'User-Agent':'3D-ECG-reproducible-research/1.0'})
                with urlopen(req,timeout=60) as response:
                    with path.with_suffix(path.suffix+'.part').open('wb') as out:
                        while block:=response.read(1024*1024):out.write(block)
                path.with_suffix(path.suffix+'.part').replace(path)
            h=digest(path)
            if expected and h!=expected:raise ValueError(f'Official checksum mismatch {rel}')
            return dict(dataset=kind,path=rel,url=url,bytes=path.stat().st_size,sha256=h,
                        official_verified=bool(expected),status='ok')
        except Exception as e:
            error=str(e)
            if isinstance(e,HTTPError) and e.code==404:break
            if isinstance(e,ValueError):raise
            if attempt<2:time.sleep(1+attempt)
    return dict(dataset=kind,path=rel,url=url,status='error',error=error)

def main():
    lock=json.loads((ROOT/'protocol_lock.json').read_text())
    assert digest(ROOT/'PROTOCOL.md')==lock['sha256']
    manifests={};rows=[]
    for kind in BASE:
        row=fetch(kind,'SHA256SUMS.txt');rows.append(row)
        if row['status']!='ok':raise RuntimeError(row)
        manifests[kind]={line.split(maxsplit=1)[1].lstrip('*./'):line.split(maxsplit=1)[0]
                         for line in (DATA/kind/'SHA256SUMS.txt').read_text().splitlines()}
        print(kind,'official checksum entries',len(manifests[kind]),flush=True)
    selection=pd.read_csv(DATA/'external_selection.csv')
    jobs=[]
    for item in selection.itertuples():
        folder=Path(item.filename_hr).parent.name
        jobs.extend([('ptbxl',item.filename_hr+ext) for ext in ['.hea','.dat']])
        jobs.extend([('plus',f'fiducial_points/ecgdeli/{folder}/{item.ecg_id:05}_points_lead_{lead}.atr')
                     for lead in ['V1','V6']])
    # Non-test record 1 validates the format only; never enters external results.
    jobs.extend([('ptbxl','records500/00000/00001_hr'+ext) for ext in ['.hea','.dat']])
    jobs.extend([('plus',f'fiducial_points/ecgdeli/00000/00001_points_lead_{lead}.atr') for lead in ['V1','V6']])
    for kind in BASE:
        for rel in ['LICENSE.txt','RECORDS','ptbxl_database.csv']:
            if rel in manifests[kind]:jobs.append((kind,rel))
    assert len(jobs)==len(set(jobs))
    for kind,rel in jobs:
        if rel not in manifests[kind]:raise ValueError(f'Not in official manifest: {kind} {rel}')
    with ThreadPoolExecutor(max_workers=8) as pool:
        futures=[pool.submit(fetch,kind,rel,manifests[kind][rel]) for kind,rel in jobs]
        for i,f in enumerate(as_completed(futures),1):
            rows.append(f.result())
            if i%200==0:
                print('Files completed',i,'of',len(jobs),'errors',sum(r['status']!='ok' for r in rows),flush=True)
                (ROOT/'download_manifest.json').write_text(json.dumps(rows,indent=2))
    (ROOT/'download_manifest.json').write_text(json.dumps(rows,indent=2))
    print('Completed',len(rows),'errors',sum(r['status']!='ok' for r in rows),flush=True)

if __name__=='__main__':main()
```
