## Supplementary material for "Calibration and processing sensitivity of time-embedded three-dimensional electrocardiographic descriptors: a two-dataset benchmark": Research materials and reproducible analysis code: make_figures.py

```
from pathlib import Path
import pandas as pd,numpy as np
import matplotlib;matplotlib.use('Agg')
import matplotlib.pyplot as plt
from matplotlib.colors import LogNorm
R=Path(__file__).resolve().parent;O=R/'results';D=R.parent/'output/paper_3decg';D.mkdir(parents=True,exist_ok=True)
plt.rcParams.update({'font.size':11,'axes.spines.top':False,'axes.spines.right':False,'savefig.facecolor':'white'})
p=pd.read_csv(O/'planned_comparisons.csv');s=pd.read_csv(O/'calibration_summary.csv')
methods=['FD','SG','SPEC','SG0SPEC'];col={'LUDB':'#151515','PTBXL':'#537b95'}
fig,(ax,txt)=plt.subplots(1,2,figsize=(8.5,4.5),gridspec_kw={'width_ratios':[3.8,1.5]})
figdata=[]
for j,cohort in enumerate(['LUDB','PTBXL']):
 for i,method in enumerate(methods):
  r=p[(p.cohort==cohort)&(p.metric=='abs_torsion')&(p.comparison==f'{method}40_vs_{method}150')].iloc[0]
  y=3-i+(-.13 if j else .13)
  ax.errorbar(r.change_median,y,xerr=[[r.change_median-r.ci025],[r.ci975-r.change_median]],fmt='o' if j==0 else 's',color=col[cohort],capsize=3,label=cohort if i==0 else None)
  txt.text(.05 if j==0 else .56,y,f'{r.spearman:.3f}',va='center',color=col[cohort])
  figdata.append(r.to_dict())
ax.set_yticks(range(4),list(reversed(methods)));ax.set_ylim(-.45,3.55);ax.set_xlabel('Median participant change from 150 to 40 Hz (%)')
ax.axvline(0,color='.6',lw=.8);ax.grid(axis='x',color='.88',lw=.6);ax.legend(frameon=False,loc='lower right')
txt.set_ylim(ax.get_ylim());txt.set_xlim(0,1);txt.axis('off');txt.text(.05,3.6,'LUDB',weight='bold');txt.text(.56,3.6,'PTB-XL',weight='bold');txt.set_title('Spearman correlation',pad=25,fontsize=11)
fig.subplots_adjust(wspace=.20,left=.12,right=.97,bottom=.18,top=.84)
fig.savefig(D/'Figure1.png',dpi=300);fig.savefig(D/'Figure1.tiff',dpi=600,pil_kwargs={'compression':'tiff_lzw'});plt.close(fig)
pd.DataFrame(figdata).to_csv(D/'Figure1_data.csv',index=False)
models=['helix_10','helix_30','helix_60','modulated_phase'];labels=['Helix 10 Hz','Helix 30 Hz','Helix 60 Hz','Modulated phase']
fig,axes=plt.subplots(1,2,figsize=(8.8,4.4));data=[]
for ax,noise,title in zip(axes,[0.,.02],['No added noise','20 microvolt noise SD']):
 mat=np.zeros((4,4))
 for i,model in enumerate(models):
  for j,method in enumerate(methods):
   r=s[(s.model==model)&(s.noise_sd_mV==noise)&(s.method==method)&(s.cutoff==150)&(s.metric=='abs_torsion')].iloc[0]
   mat[i,j]=r.relative_rmse_percent;data.append(r.to_dict())
 im=ax.imshow(np.maximum(mat,1e-4),norm=LogNorm(1e-4,1000),cmap='Greys',aspect='auto')
 ax.set_xticks(range(4),methods,rotation=25,ha='right');ax.set_yticks(range(4),labels if ax is axes[0] else ['']*4)
 for i in range(4):
  for j in range(4):
   v=mat[i,j];label='<0.001' if v<.001 else f'{v:.3g}'
   ax.text(j,i,label,ha='center',va='center',fontsize=10.5,color='white' if v>1 else 'black')
 ax.set_title(title,pad=12);ax.tick_params(length=0)
fig.subplots_adjust(left=.18,right=.88,bottom=.23,top=.87,wspace=.12)
cax=fig.add_axes([.90,.23,.018,.64]);cb=fig.colorbar(im,cax=cax);cb.set_label('Relative RMSE (%)',fontsize=10)
fig.savefig(D/'Figure2.png',dpi=300);fig.savefig(D/'Figure2.tiff',dpi=600,pil_kwargs={'compression':'tiff_lzw'});plt.close(fig)
pd.DataFrame(data).to_csv(D/'Figure2_data.csv',index=False)
print('Two figures and their exact source-data tables saved')
```
