## Supplementary material for "Calibration and processing sensitivity of time-embedded three-dimensional electrocardiographic descriptors: a two-dataset benchmark": Research materials and reproducible analysis code: PROTOCOL.md

# Locked computational study protocol

This third phase follows exploratory analyses of 200 LUDB participants and 1,748 QRS windows. It is not a public preregistration. The protocol is archived before computing the new estimators or inspecting external 3D ECG measurements. Metadata and annotation-file layout are inspected for feasibility. A non-test PTB-XL record may be used only to validate the reader and annotation convention.

## Question and scope

Determine how coordinate conventions and derivative operators affect time-embedded 3D ECG descriptors, calibrate an explicit spectral derivative implementation against analytic controls, and replicate processing comparisons in an external public cohort. Do not claim clinical accuracy, diagnostic improvement, a universally optimal method, new physiological information from a deterministic representation, or novelty of Fourier differentiation itself.

## Cohorts and selection

Development: all 200 LUDB 1.0.1 participants with exactly the 1,748 windows archived in phase 1.

External: PTB-XL 1.0.3, metadata fold 10. Retain the lowest ECG identifier for each patient. Rank patients by SHA-256 of `3decg-external-20260914:` followed by the integer patient identifier; take the first 1,000. This is a fixed computational sample, not a power-derived sample size. The remaining patients are not screened for more favorable results. Save the selected identifiers before downloading signals. Do not replace missing or unusable selected records.

Use original 500 Hz, 12-lead, 10-second waveforms in physical mV. Reuse PTB-XL+ 1.0.1 ECGDeli annotations for V1 and V6, verifying sample coordinates and marker meaning. These are automatic, not manually adjudicated QRS boundaries. Document incomplete markers, mutual nearest QRS matches within 100 ms, exclusion of union windows shorter than 40 or longer than 300 ms, and exclusion within one second of the record edges. If the annotation format is incompatible with this rule, document a dated amendment before looking at descriptor outcomes. Reject records with invalid lengths, frequency, units, or nonfinite voltage. Report all exclusions.

## Fixed geometry and processing

Coordinates are V1/(1 mV), V6/(1 mV), and t/(1 ms). Subtract each lead's whole-record median; do not normalize amplitude per person. Filters are fourth-order digital Butterworth with nominal 40, 100, and 150 Hz cutoffs, applied in both directions as in phases 1 and 2. Nominal frequencies refer to the single-pass design; the double-pass response is about -6 dB at cutoff.

Compare four derivative operators, with no patient- or outcome-specific tuning:

1. FD: successive second-order central differences, step 2 ms.
2. SG: Savitzky-Golay direct derivatives of orders 1, 2 and 3, degree 3, 11 samples.
3. SPEC: derivatives of a common trigonometric interpolant of the filtered signal, using an even reflection of the complete record (length 2N-2) and Fourier multipliers (i omega)^k, where omega is in radians/ms. For an even-length extension, remove the Nyquist component for all orders to define a common interpolant. Crop to the original record. This is an explicit continuous reconstruction convention, not proof that the underlying biological signal is band-limited or known.
4. SG0SPEC: smooth with SG degree 3 and 11 samples at derivative order zero, then apply SPEC. This control tests the role of added smoothing while retaining a common interpolation for differentiation.

For calibration with analytic controls, use the same filtering and reflection as for ECGs. Additionally evaluate a periodic-input spectral calculation on noiseless periodic controls to verify exact multipliers independently of boundary extension.

Primary descriptor: temporal mean absolute classical torsion in each QRS, followed by median across a person's QRS windows. Secondary: mean classical curvature, mean equation-6 almost-curvature with fixed axes, and denominator-weighted absolute torsion. Do not treat the last descriptor as the same estimand as temporal mean absolute torsion. The denominator must exceed machine epsilon times its record maximum; report invalid-duration fractions. Summaries with any invalid QRS samples are missing for that descriptor, with explicit counts and paired-complete comparisons.

## Mathematical and numerical calibration

Retain the analytic straight-line, rotation, reparametrization, torsion identity and Cauchy-Schwarz checks from phase 2. External coordinate audit: FD at 150 Hz, rotation by 90 degrees in the voltage plane and reparametrization by seconds while retaining the same coordinates; also the deliberate change from t/(1 ms) to t/(100 ms), labeled as a different geometry.

Analytic signals: unit-radius helices at 10, 30 and 60 Hz, and the phase-modulated circle phi(t)=2 pi 20 t/1000 + 0.8 sin(2 pi 3 t/1000), with t in ms. Generate 8-second records at 500 Hz and assess the central interval from 2 to 6 seconds. Noise SD 0, 0.005 and 0.02 mV, independent Gaussian noise in both coordinates; 1 realization at zero noise and 100 at each other noise level, seed 20260917. These levels are calibration scenarios, not estimates of patient noise. All methods share each realization. Report signed bias and relative RMSE against each descriptor's analytic value; do not label mathematical signals as simulated patients.

For the helices, separately report the exact target after the known Butterworth amplitude response. This distinguishes changing the curve by filtering from derivative error. No claim of a single true filtered target is made for the modulated control unless computed analytically. Report both biological-input-free analytic targets and the explicitly defined filtered-helix targets.

## Comparisons and uncertainty

Primary external contrast: SPEC versus FD at 150 Hz for absolute torsion, reporting paired percentage difference, absolute percentage difference, and Spearman rank correlation. This is operator disagreement, not measured clinical error. Also report SG versus FD and SG0SPEC versus FD at 150 Hz, plus 40 versus 150 and 100 versus 150 within every method, for all four descriptors. Publish the complete planned comparisons, including unfavorable results.

Use participant-level aggregation and paired percentile bootstrap confidence intervals for medians (5,000 resamples, seed 20260918). Do not count multiple beats as independent people. Do not rank approaches by p values or select an estimator using external outcomes. Confidence intervals describe sampling uncertainty, not uncertainty in an individual's ECG measurement. Correlation is not agreement; report changes in magnitude alongside it.

Boundary sensitivity: repeat SPEC and FD at 150 Hz after shifting each QRS onset and offset by -10 or +10 ms, all four combinations, without redetecting beats. Analyze only windows valid under all shifts, report the largest absolute percentage change per person as a descriptive sensitivity measure. Do not call the maximum a confidence interval.

Boundary-extension sensitivity: for SPEC at 150 Hz, compare reflected versus periodic full-record interpolation, using the same QRS windows. This documents extension effects, not a clinical performance endpoint.

## Reproducibility and manuscript

Store source versions, downloaded files, publisher checksums where supplied, local SHA-256 hashes, per-window values, exclusions, participant summaries, comparisons and calibration results. Verify aggregation independently and test analytic identities and spectral frequency predictions. Preserve prior phases without overwriting. Generate manuscript numbers and tables from result files. Write a full English original research manuscript with abstract, introduction, methods, results, discussion, limitations, conclusions, references and supplementary methods. Do not assert human author review, ethics exemption, funding, authorship or competing interests without confirmation. No journal submission is included.

Distribution note: one editorial instruction has been omitted from this copy; numerical methods are unchanged. The original protocol lock records the unabridged source, not this distributed copy.
