## Supplementary material for "Calibration and processing sensitivity of time-embedded three-dimensional electrocardiographic descriptors: a two-dataset benchmark": Research materials and reproducible analysis code: summarize_verify.py

```
"""Independent aggregation checks and planned comparisons from saved results."""
from pathlib import Path
from hashlib import sha256
import sys,json
import numpy as np
import pandas as pd
from scipy.stats import spearmanr
R=Path(__file__).resolve().parent;O=R/'results';sys.path.insert(0,str(R));import analysis as a
rng=np.random.default_rng(20260918)

def comparison(x,y):
    pair=pd.concat([x.rename('x'),y.rename('y')],axis=1).dropna();pair=pair[pair.y!=0]
    x=pair.x.to_numpy();y=pair.y.to_numpy();v=100*(x-y)/y
    if not len(v):return {'n':0}
    bootstrap=np.median(v[rng.integers(0,len(v),size=(5000,len(v)))],axis=1)
    return dict(n=len(v),candidate_median=float(np.median(x)),reference_median=float(np.median(y)),
                change_median=float(np.median(v)),q25=float(np.quantile(v,.25)),q75=float(np.quantile(v,.75)),
                ci025=float(np.quantile(bootstrap,.025)),ci975=float(np.quantile(bootstrap,.975)),
                absolute_change_median=float(np.median(abs(v))),
                spearman=float(spearmanr(x,y).statistic))

rows=[];geometry=[];boundary=[];extension=[];cohorts=[];auditchecks={}
for cohort in ['LUDB','PTBXL']:
    beats=pd.read_csv(O/f'{cohort}_beats.csv');p=pd.read_csv(O/f'{cohort}_participants.csv')
    w=pd.read_csv(O/f'{cohort}_windows.csv');aud=pd.read_csv(O/f'{cohort}_audit.csv')
    assert not beats.duplicated(['patient','record','beat','cutoff','method']).any()
    assert (beats.groupby(['patient','record','beat']).size()==12).all()
    assert len(w)*12==len(beats)
    assert p.groupby('patient').record.nunique().max()==1
    for m in a.METRICS:
        computed=beats.groupby(['cohort','patient','record','cutoff','method'])[m].median().sort_index()
        saved=p.set_index(['cohort','patient','record','cutoff','method'])[m].sort_index()
        np.testing.assert_allclose(computed,saved,rtol=1e-11,atol=1e-12,equal_nan=True)
    assert int(aud.eligible.sum())==len(w)
    cohorts.append(dict(cohort=cohort,selected=len(aud),included=p.patient.nunique(),excluded=len(aud)-p.patient.nunique(),
                        qrs=len(w),qrs_per_person_median=float(w.groupby('patient').size().median()),
                        qrs_min=int(w.groupby('patient').size().min()),qrs_max=int(w.groupby('patient').size().max()),
                        qrs_duration_median_ms=float(np.median((w.end_sample-w.start_sample)*2)),
                        invalid_torsion_measurement_rows=int(beats.abs_torsion.isna().sum())))
    for metric in a.METRICS:
        base=p[(p.method=='FD')&(p.cutoff==150)].set_index('patient')[metric]
        for method in ['SG','SPEC','SG0SPEC']:
            x=p[(p.method==method)&(p.cutoff==150)].set_index('patient')[metric]
            rows.append(dict(cohort=cohort,metric=metric,comparison=f'{method}150_vs_FD150',**comparison(x,base)))
        for method in a.METHODS:
            y=p[(p.method==method)&(p.cutoff==150)].set_index('patient')[metric]
            for cut in [40,100]:
                x=p[(p.method==method)&(p.cutoff==cut)].set_index('patient')[metric]
                rows.append(dict(cohort=cohort,metric=metric,comparison=f'{method}{cut}_vs_{method}150',**comparison(x,y)))
    g=pd.read_csv(O/f'{cohort}_geometry_beats.csv').groupby(['patient','variant'])[a.METRICS].median()
    for metric in a.METRICS:
        y=g.xs('baseline',level='variant')[metric]
        for variant in ['rotation90','parameter_seconds','time_scale100']:
            x=g.xs(variant,level='variant')[metric]
            geometry.append(dict(cohort=cohort,metric=metric,variant=variant,**comparison(x,y)))
    b=pd.read_csv(O/f'{cohort}_boundary_beats.csv')
    b=b.groupby(['patient','method','onset_shift_ms','offset_shift_ms'])[a.METRICS].median().reset_index()
    for method in ['FD','SPEC']:
        ref=p[(p.method==method)&(p.cutoff==150)].set_index('patient')
        for metric in a.METRICS:
            vals=b[b.method==method].pivot(index='patient',columns=['onset_shift_ms','offset_shift_ms'],values=metric)
            pct=100*(vals.div(ref[metric],axis=0)-1)
            worst=pct.abs().max(axis=1,skipna=False).dropna()
            boundary.append(dict(cohort=cohort,method=method,metric=metric,n=len(worst),
                                 max_absolute_change_median=float(worst.median()),q25=float(worst.quantile(.25)),q75=float(worst.quantile(.75))))
    e=pd.read_csv(O/f'{cohort}_extension_beats.csv').groupby('patient')[a.METRICS].median()
    ref=p[(p.method=='SPEC')&(p.cutoff==150)].set_index('patient')
    for m in a.METRICS:extension.append(dict(cohort=cohort,metric=m,**comparison(e[m],ref[m])))
    auditchecks[cohort]='all participant aggregations, duplicate checks, 12 conditions and inclusion totals passed'
pd.DataFrame(rows).to_csv(O/'planned_comparisons.csv',index=False)
pd.DataFrame(geometry).to_csv(O/'geometry_comparisons.csv',index=False)
pd.DataFrame(boundary).to_csv(O/'boundary_sensitivity.csv',index=False)
pd.DataFrame(extension).to_csv(O/'extension_sensitivity.csv',index=False)
pd.DataFrame(cohorts).to_csv(O/'cohort_summary.csv',index=False)

# Check that copied metadata used for selection agrees with the official checksum-verified file.
assert (R/'data/ptbxl_database.csv').read_bytes()==(R/'data/ptbxl/ptbxl_database.csv').read_bytes()
meta=pd.read_csv(R/'data/ptbxl_database.csv');sel=pd.read_csv(R/'data/external_selection.csv')
candidate=meta[meta.strat_fold==10].sort_values('ecg_id').drop_duplicates('patient_id').copy()
candidate['selection_hash']=candidate.patient_id.map(lambda x:sha256(f'3decg-external-20260914:{int(x)}'.encode()).hexdigest())
assert candidate.sort_values('selection_hash').head(1000).ecg_id.tolist()==sel.ecg_id.tolist()
assert sel.patient_id.nunique()==len(sel)==1000
for task in ['LUDB','PTBXL','calibration']:
    prov=json.loads((O/f'{task}_provenance.json').read_text())
    assert prov['script_sha256']==sha256((R/'analysis.py').read_bytes()).hexdigest()
    assert prov['protocol_sha256']==sha256((R/'PROTOCOL.md').read_bytes()).hexdigest()
# Algebraic verification using a held-out random smooth curve, no clinical outcomes.
t=np.arange(1000)*2.;xy=np.column_stack((np.sin(.023*t)+.2*np.cos(.1*t),np.cos(.051*t)))
d=a.spectral(xy);g=a.geometry(d);p,q,s=d
D=np.sum(q*q,axis=1)+(p[:,0]*q[:,1]-p[:,1]*q[:,0])**2
N=q[:,0]*s[:,1]-q[:,1]*s[:,0]
np.testing.assert_allclose(g['abs_torsion'],abs(N/D),rtol=1e-10,atol=1e-12)
# Explicit Fourier summation checks the multipliers independently of irfft.
ext=np.concatenate((xy,xy[-2:0:-1]));ft=np.fft.fft(ext,axis=0);n=len(ext)
w=2*np.pi*np.fft.fftfreq(n,d=2.);ft[n//2]=0
maxerror=0.
for k in [1,2,3]:
    for idx in [101,223,444,701,899]:
        direct=np.real(np.sum(ft*(1j*w[:,None])**k*np.exp(1j*w[:,None]*idx*2),axis=0)/n)
        np.testing.assert_allclose(d[k-1][idx],direct,rtol=1e-8,atol=1e-11)
        maxerror=max(maxerror,float(np.max(abs(d[k-1][idx]-direct))))
# Analytic helix and degenerate curve behavior.
omega=.1;phase=omega*t
hd=(np.column_stack((-omega*np.sin(phase),omega*np.cos(phase))),
    np.column_stack((-omega**2*np.cos(phase),-omega**2*np.sin(phase))),
    np.column_stack((omega**3*np.sin(phase),-omega**3*np.cos(phase))))
np.testing.assert_allclose(a.geometry(hd)['abs_torsion'],omega/(1+omega**2),atol=1e-14)
z=np.zeros((1000,2));line=a.geometry((np.ones_like(z),z,z));assert np.isnan(line['abs_torsion']).all()
# Reproduce prior FD results, which use the same LUDB processing and windows.
old=pd.read_csv(R.parent/'estudio_3decg_reproducibilidad/results/subject_metrics.csv')
new=pd.read_csv(O/'LUDB_participants.csv')
for cut in [40,100,150]:
    left=new[(new.method=='FD')&(new.cutoff==cut)].set_index('record').sort_index()
    right=old[old.condition==f'fd{cut}'].set_index('record').sort_index()
    for m,om in [('abs_torsion','abs_torsion_mean'),('curvature','curvature_mean'),('almost_curvature','almost_curvature_mean')]:
        np.testing.assert_allclose(left[m],right[om],rtol=1e-10,atol=1e-12)
checks={'cohort_checks':auditchecks,'selection':'reproduced exactly','metadata':'identical to official verified file',
        'spectral_direct_sum_max_absolute_error':maxerror,'scalar_vector_identity':'passed',
        'analytic_helix':'passed','straight_line_torsion':'undefined as expected','prior_LUDB_reproduction':'passed',
        'comparison_rows':len(rows),'no_clinical_accuracy_validation':True}
(O/'verification.json').write_text(json.dumps(checks,indent=2));print(json.dumps(checks,indent=2))
print(pd.DataFrame(cohorts).to_string(index=False))
```
