## Supplementary material for "Calibration and processing sensitivity of time-embedded three-dimensional electrocardiographic descriptors: a two-dataset benchmark": Research materials and reproducible analysis code: verify_calibration_and_units.py

```
"""Cross-check saved calibration summaries and physical waveform scaling."""
from pathlib import Path
import json
import numpy as np
import pandas as pd
import wfdb

R=Path(__file__).resolve().parent
est=pd.read_csv(R/'results/calibration_estimates.csv')
summ=pd.read_csv(R/'results/calibration_summary.csv')
keys=['model','noise_sd_mV','cutoff','method','metric']
assert len(summ)==576
assert not est.duplicated(keys+['realization']).any()
assert len(est[['model','noise_sd_mV','realization']].drop_duplicates())==804
checked=0
for row in summ.itertuples(index=False):
    mask=np.ones(len(est),dtype=bool)
    for k in keys: mask &= est[k].to_numpy()==getattr(row,k)
    group=est.loc[mask]
    assert len(group)==(1 if row.noise_sd_mV==0 else 100)==row.n
    residual=group.estimate.to_numpy()-group.truth.to_numpy()
    assert np.isfinite(residual).all()
    target=group.truth.iloc[0]
    value=100*np.linalg.norm(residual)/np.sqrt(len(group))/abs(target)
    np.testing.assert_allclose(value,row.relative_rmse_percent,rtol=1e-7,atol=1e-9)
    if group.filtered_truth.notna().all():
        residual=group.estimate.to_numpy()-group.filtered_truth.to_numpy()
        value=100*np.linalg.norm(residual)/np.sqrt(len(group))/abs(group.filtered_truth.iloc[0])
        np.testing.assert_allclose(value,row.filtered_relative_rmse_percent,rtol=1e-7,atol=1e-9)
    checked+=1

selection=pd.read_csv(R/'data/external_selection.csv')
units=[]
for row in selection.head(3).itertuples():
    path=str(R/'data/ptbxl'/row.filename_hr)
    digital=wfdb.rdrecord(path,physical=False)
    physical=wfdb.rdrecord(path,physical=True)
    for lead in ['V1','V6']:
        j=digital.sig_name.index(lead)
        assert digital.units[j]=='mV'
        explicit=(digital.d_signal[:,j].astype(float)-digital.baseline[j])/digital.adc_gain[j]
        np.testing.assert_allclose(explicit,physical.p_signal[:,j],rtol=0,atol=0)
        units.append({'ecg_id':row.ecg_id,'lead':lead,'samples_checked':len(explicit),'unit':'mV'})
meta=pd.read_csv(R/'data/ptbxl_database.csv')
assert int(meta.loc[meta.ecg_id==1,'strat_fold'].iloc[0])!=10
assert 1 not in set(selection.ecg_id)
report={'calibration_summary_rows_verified':checked,'signal_noise_realizations':804,
        'explicit_digital_to_physical_checks':units,'reader_calibration_record_excluded':True}
(R/'results/additional_verification.json').write_text(json.dumps(report,indent=2))
print(json.dumps(report,indent=2))
```
