## Supplementary material for "Calibration and processing sensitivity of time-embedded three-dimensional electrocardiographic descriptors: a two-dataset benchmark": Research materials and reproducible analysis code: analyze.py

```
"""Exploratory LUDB sensitivity audit. No diagnostic labels or synthetic patients.

Coordinates are numerical values V1/(1 mV), V6/(1 mV), t/(1 ms).
Geometry therefore depends on this fixed scale convention, not anatomical units.
"""
from pathlib import Path
from datetime import datetime, timezone
import argparse
import hashlib
import json
import platform
import importlib.metadata as metadata
import numpy as np
import pandas as pd
import wfdb
from scipy.signal import butter, sosfiltfilt, savgol_filter, resample_poly
from scipy.stats import spearmanr

ROOT = Path(__file__).resolve().parent
OUT = ROOT/'results'
OUT.mkdir(exist_ok=True)
DATA = next((ROOT/'data').glob('*/RECORDS')).parent
METRICS = ['arc_length', 'curvature_mean', 'almost_curvature_mean',
           'torsion_mean', 'abs_torsion_mean', 'shape_singular_ratio']
COMPARISONS = [('fd40', 'fd150'), ('fd100', 'fd150'),
               ('fd40_250', 'fd40'), ('sg40', 'sg150')]

def digest(p):
    return hashlib.sha256(p.read_bytes()).hexdigest()

def qrs_annotations(record, lead):
    a = wfdb.rdann(str(DATA/record), lead)
    result = []
    incomplete = 0
    for j, symbol in enumerate(a.symbol):
        if symbol == 'N':
            if not (0 < j < len(a.symbol)-1 and a.symbol[j-1] == '(' and a.symbol[j+1] == ')'):
                incomplete += 1
                continue
            onset, peak, offset = map(int, a.sample[j-1:j+2])
            assert onset <= peak <= offset
            result.append((onset, peak, offset))
    return np.array(result, dtype=int).reshape(-1,3), incomplete

def windows(record, fs, length):
    (a, ia), (b, ib) = (qrs_annotations(record, lead) for lead in ('v1','v6'))
    audit = dict(record=record, qrs_v1=len(a), qrs_v6=len(b),
                 incomplete_v1=ia, incomplete_v6=ib, matched=0,
                 excluded_duration=0, excluded_edge=0, eligible=0)
    selected = []
    if len(a) and len(b):
        distances = abs(a[:,1,None]-b[None,:,1])
        for i, j in enumerate(distances.argmin(axis=1)):
            if distances[i,j] > .1*fs or distances[:,j].argmin() != i:
                continue
            audit['matched'] += 1
            lo, hi = min(a[i,0], b[j,0]), max(a[i,2], b[j,2])
            duration = (hi-lo)/fs
            if duration < .04 or duration > .3:
                audit['excluded_duration'] += 1
                continue
            if lo < fs or hi > length-1-fs:
                audit['excluded_edge'] += 1
                continue
            selected.append(dict(record=record, beat=len(selected)+1,
                start_sample=int(lo), end_sample=int(hi),
                v1_peak=int(a[i,1]), v6_peak=int(b[j,1]),
                duration_ms=duration*1000))
    audit['eligible'] = len(selected)
    audit['unmatched_v1'] = len(a)-audit['matched']
    audit['unmatched_v6'] = len(b)-audit['matched']
    assert audit['matched'] == audit['eligible']+audit['excluded_duration']+audit['excluded_edge']
    return selected, audit

def read(record):
    r = wfdb.rdrecord(str(DATA/record))
    assert r.fs == 500 and r.sig_len == 5000 and r.n_sig == 12
    channels = [r.sig_name.index(name) for name in ('v1','v6')]
    assert all(r.units[i] == 'mV' for i in channels)
    xy = r.p_signal[:,channels]
    assert np.isfinite(xy).all()
    return r, xy-np.median(xy, axis=0)

def lowpass(xy, cutoff, fs=500):
    # Nominal single-pass -3 dB frequency; two passes yield about -6 dB there.
    sos = butter(4, cutoff, fs=fs, btype='lowpass', output='sos')
    return sosfiltfilt(sos, xy, axis=0, padtype='odd', padlen=15)

def derivatives(xy, dt, method):
    if method == 'fd':
        d1 = np.gradient(xy, dt, axis=0, edge_order=2)
        d2 = np.gradient(d1, dt, axis=0, edge_order=2)
        d3 = np.gradient(d2, dt, axis=0, edge_order=2)
    elif method == 'sg':
        assert dt == 2
        d1,d2,d3 = [savgol_filter(xy, 11, 3, deriv=d, delta=dt,
                           axis=0, mode='interp') for d in (1,2,3)]
    else:
        raise ValueError(method)
    return d1,d2,d3

def from_derivatives(d1,d2,d3):
    a,b = d1.T
    aa,bb = d2.T
    aaa,bbb = d3.T
    speed2 = 1+a*a+b*b
    den = aa*aa+bb*bb+(a*bb-b*aa)**2
    tau = np.full(len(a), np.nan)
    threshold = np.finfo(float).eps*np.max(den)
    valid = den > threshold
    tau[valid] = (aa[valid]*bbb[valid]-bb[valid]*aaa[valid])/den[valid]
    # Independent vector identity for every processed sample.
    p1 = np.column_stack((d1,np.ones(len(a))))
    p2 = np.column_stack((d2,np.zeros(len(a))))
    p3 = np.column_stack((d3,np.zeros(len(a))))
    cross = np.cross(p1,p2)
    vector_den = np.sum(cross*cross,axis=1)
    vector_tau = np.sum(cross*p3,axis=1)[valid]/vector_den[valid]
    np.testing.assert_allclose(tau[valid], vector_tau, rtol=1e-11, atol=1e-12)
    np.testing.assert_allclose(den, vector_den, rtol=1e-12, atol=1e-18)
    # Eq.6, 2025 paper, with the coordinate order x=V1,y=V6,z=time.
    almost = np.sqrt(bb*bb+a*a+(aa*b-a*bb)**2)/speed2**1.5
    return dict(speed=np.sqrt(speed2), curvature=np.sqrt(den)/speed2**1.5,
                almost=almost, torsion=tau)

def geometry(xy, dt, method):
    return from_derivatives(*derivatives(xy,dt,method))

def crop(t, arr, lo, hi):
    # Preserve identical physical endpoints after downsampling. Linear endpoint
    # interpolation only; no time warping and no extrapolation.
    inside=(t>lo)&(t<hi)
    target=np.r_[lo,t[inside],hi]
    if arr.ndim==1:
        values=np.interp(target,t,arr)
    else:
        values=np.column_stack([np.interp(target,t,arr[:,j]) for j in range(arr.shape[1])])
    return target,values

def valid_integral_mean(t,values):
    ok=np.isfinite(values[:-1])&np.isfinite(values[1:])
    spans=np.diff(t)
    duration=np.sum(spans[ok])
    if duration==0:
        return np.nan,0.
    integral=np.sum(((values[:-1]+values[1:])/2)[ok]*spans[ok])
    return integral/duration,duration/(t[-1]-t[0])

def beat_metrics(t,xy,geom,lo,hi):
    result={}
    for name,source in [('curvature_mean','curvature'),
                        ('almost_curvature_mean','almost'),('torsion_mean','torsion'),
                        ('abs_torsion_mean','torsion')]:
        tt,v=crop(t,geom[source],lo,hi)
        if name=='abs_torsion_mean':
            v=np.abs(v)
        value,fraction=valid_integral_mean(tt,v)
        result[name]=value
        if name=='abs_torsion_mean':
            result['torsion_valid_duration_fraction']=fraction
            result['torsion_valid_sample_fraction']=float(np.isfinite(v).mean())
    times,points=crop(t,xy,lo,hi)
    curve=np.column_stack((points,times))
    result['arc_length']=float(np.linalg.norm(np.diff(curve,axis=0),axis=1).sum())
    sv=np.linalg.svd(points-points.mean(axis=0),compute_uv=False)
    result['shape_singular_ratio']=sv[-1]/sv[0] if sv[0]>0 else np.nan
    return result

def checks():
    # Analytic controls are mathematical, not observations from patients.
    z=np.zeros((100,2))
    line=from_derivatives(np.ones((100,2)),z,z)
    assert np.max(line['curvature'])==0
    assert np.isnan(line['torsion']).all()
    t=np.linspace(0,6,1000)
    radius=2.
    d1=np.column_stack((-radius*np.sin(t),radius*np.cos(t)))
    d2=np.column_stack((-radius*np.cos(t),-radius*np.sin(t)))
    d3=-d1
    h=from_derivatives(d1,d2,d3)
    np.testing.assert_allclose(h['curvature'],2/5,atol=1e-14)
    np.testing.assert_allclose(h['torsion'],1/5,atol=1e-14)
    hn=geometry(np.column_stack((radius*np.cos(t),radius*np.sin(t))),t[1]-t[0],'fd')
    np.testing.assert_allclose(hn['torsion'][5:-5],1/5,rtol=1e-4)
    a,f=valid_integral_mean(np.arange(5),np.array([1.,1.,np.nan,3.,3.]))
    assert a==2 and f==.5
    return {'straight_line':'pass; torsion undefined', 'analytic_helix':'pass',
            'finite_difference_helix':'pass', 'no_bridging_invalid_gaps':'pass',
            'vector_scalar_torsion':'asserted for all processed samples'}

def preview():
    import matplotlib
    matplotlib.use('Agg')
    import matplotlib.pyplot as plt
    fig, axes=plt.subplots(5,2,figsize=(12,12))
    for row,record in enumerate(['data/1','data/50','data/100','data/150','data/200']):
        r,xy=read(record)
        beats,_=windows(record,r.fs,r.sig_len)
        t=np.arange(len(xy))*2
        for j,lead in enumerate(['V1','V6']):
            ax=axes[row,j]
            ax.plot(t/1000,xy[:,j],color='.65',lw=.7,label='Original')
            ax.plot(t/1000,lowpass(xy,40)[:,j],color='black',lw=.8,label='40 Hz')
            for b in beats:
                ax.axvspan(b['start_sample']/500,b['end_sample']/500,color='#93bdd3',alpha=.3)
            ax.set_title(f"LUDB {record.split('/')[-1]} - {lead}; {len(beats)} QRS")
            ax.set_ylabel('mV'); ax.set_xlabel('Tiempo (s)')
    fig.tight_layout()
    fig.savefig(OUT/'qa_windows.png',dpi=150)
    plt.close(fig)

def bootstrap_median(values,rng):
    values=np.asarray(values)
    values=values[np.isfinite(values)]
    if not len(values): return [np.nan,np.nan]
    boot=np.median(values[rng.integers(0,len(values),size=(5000,len(values)))],axis=1)
    return np.quantile(boot,[.025,.975]).tolist()

def summarize(subjects):
    rng=np.random.default_rng(20260914)
    comparisons=[]
    individual=[]
    for candidate,reference in COMPARISONS:
        for metric in METRICS:
            left=subjects[subjects.condition==candidate].set_index('record')[metric]
            right=subjects[subjects.condition==reference].set_index('record')[metric]
            paired=pd.concat([left.rename('candidate'),right.rename('reference')],axis=1).dropna()
            a,b=paired.candidate.to_numpy(),paired.reference.to_numpy()
            diff=a-b
            pct=np.divide(100*diff,b,out=np.full(len(a),np.nan),where=b!=0)
            sym=np.divide(200*np.abs(diff),np.abs(a)+np.abs(b),
                          out=np.zeros(len(a)),where=(np.abs(a)+np.abs(b))>0)
            lo,hi=bootstrap_median(pct,rng)
            row=dict(comparison=f'{candidate}_vs_{reference}',metric=metric,n=len(a),
                     candidate_median=float(np.median(a)),reference_median=float(np.median(b)),
                     difference_median=float(np.median(diff)),
                     percent_change_median=float(np.nanmedian(pct)),
                     percent_change_q25=float(np.nanquantile(pct,.25)),
                     percent_change_q75=float(np.nanquantile(pct,.75)),
                     percent_change_ci025=lo,percent_change_ci975=hi,
                     absolute_percent_change_median=float(np.nanmedian(abs(pct))),
                     symmetric_relative_difference_median=float(np.median(sym)),
                     spearman_rho=float(spearmanr(a,b).statistic))
            comparisons.append(row)
            for k,record in enumerate(paired.index):
                individual.append(dict(record=record,comparison=row['comparison'],metric=metric,
                    candidate=a[k],reference=b[k],difference=diff[k],percent_change=pct[k],
                    symmetric_relative_difference=sym[k]))
    pd.DataFrame(comparisons).to_csv(OUT/'comparisons.csv',index=False)
    indiv=pd.DataFrame(individual)
    indiv.to_csv(OUT/'individual_changes.csv',index=False)
    fd=indiv[(indiv.comparison=='fd40_vs_fd150')&(indiv.metric=='abs_torsion_mean')].set_index('record')
    sg=indiv[(indiv.comparison=='sg40_vs_sg150')&(indiv.metric=='abs_torsion_mean')].set_index('record')
    paired=pd.concat([abs(fd.percent_change).rename('fd'),abs(sg.percent_change).rename('sg')],axis=1).dropna()
    delta=paired.sg-paired.fd
    return dict(sg_minus_fd_absolute_percent_change_median=float(delta.median()),
                ci95=bootstrap_median(delta.to_numpy(),rng),n=len(delta))

def run():
    lock=ROOT/'plan_lock.json'
    if not lock.exists():
        lock.write_text(json.dumps(dict(plan_sha256=digest(ROOT/'PLAN_PILOTO.md'),
                         created_utc=datetime.now(timezone.utc).isoformat()),indent=2))
    assert json.loads(lock.read_text())['plan_sha256']==digest(ROOT/'PLAN_PILOTO.md')
    validation=checks()
    beats_all=[]; audits=[]; windows_all=[]; headers=[]
    records=(DATA/'RECORDS').read_text().splitlines()
    assert len(records)==len(set(records))==200
    records=sorted(records,key=lambda r:int(r.split('/')[-1]))
    for record in records:
        r,xy=read(record)
        windows_record,audit=windows(record,r.fs,r.sig_len)
        audits.append(audit); windows_all.extend(windows_record)
        headers.append(dict(record=record,fs=r.fs,samples=r.sig_len,channels=r.n_sig,
                            units='mV',comments=' | '.join(r.comments)))
        signals={f'fd{cutoff}':(lowpass(xy,cutoff),2.,'fd') for cutoff in (40,100,150)}
        signals['fd40_250']=(resample_poly(signals['fd40'][0],1,2,axis=0,
                                window=('kaiser',5.),padtype='constant'),4.,'fd')
        signals['sg40']=(signals['fd40'][0],2.,'sg')
        signals['sg150']=(signals['fd150'][0],2.,'sg')
        for condition,(signal,dt,method) in signals.items():
            t=np.arange(len(signal))*dt
            geom=geometry(signal,dt,method)
            for beat in windows_record:
                row=dict(record=record,beat=beat['beat'],condition=condition)
                row.update(beat_metrics(t,signal,geom,beat['start_sample']*2,beat['end_sample']*2))
                beats_all.append(row)
        if int(record.split('/')[-1])%50==0:
            print(f"Processed {record}",flush=True)
    beats=pd.DataFrame(beats_all)
    subjects=beats.groupby(['record','condition'],sort=True)[METRICS+['torsion_valid_duration_fraction','torsion_valid_sample_fraction']].median().reset_index()
    beats.to_csv(OUT/'beat_metrics.csv',index=False)
    subjects.to_csv(OUT/'subject_metrics.csv',index=False)
    pd.DataFrame(audits).to_csv(OUT/'inclusion_audit.csv',index=False)
    pd.DataFrame(windows_all).to_csv(OUT/'qrs_windows.csv',index=False)
    pd.DataFrame(headers).to_csv(OUT/'record_metadata.csv',index=False)
    estimator=summarize(subjects)
    validation['records_with_missing_metrics']=int(subjects[METRICS].isna().any(axis=1).sum())
    validation['minimum_valid_torsion_duration_fraction']=float(beats.torsion_valid_duration_fraction.min())
    audit_df=pd.DataFrame(audits)
    totals={col:int(audit_df[col].sum()) for col in audit_df.columns if col!='record'}
    summary=dict(dataset='LUDB 1.0.1',records_loaded=200,records_analyzed=int(subjects.record.nunique()),
                 eligible_beats=len(windows_all),conditions=6,totals=totals,validation=validation,
                 sg_exploratory_comparison=estimator,script_sha256=digest(Path(__file__)),
                 plan_sha256=digest(ROOT/'PLAN_PILOTO.md'),run_utc=datetime.now(timezone.utc).isoformat())
    (OUT/'run_summary.json').write_text(json.dumps(summary,indent=2))
    versions={name:metadata.version(name) for name in ['numpy','pandas','scipy','wfdb','matplotlib']}
    versions.update(python=platform.python_version(),platform=platform.platform())
    (ROOT/'versions.json').write_text(json.dumps(versions,indent=2))
    print(json.dumps(summary,indent=2),flush=True)

if __name__=='__main__':
    p=argparse.ArgumentParser()
    p.add_argument('--preview',action='store_true')
    args=p.parse_args()
    preview() if args.preview else run()
```
