## Supplementary material for "Calibration and processing sensitivity of time-embedded three-dimensional electrocardiographic descriptors: a two-dataset benchmark": Research materials and reproducible analysis code: download_data.py

```
"""Download the official LUDB archive; retain provenance and safe extraction."""
from pathlib import Path
from urllib.request import urlopen
from urllib.parse import urljoin
import hashlib
import json
import re
import zipfile
from datetime import datetime, timezone

ROOT = Path(__file__).resolve().parent
RAW = ROOT / 'data'
RAW.mkdir(exist_ok=True)
PAGE = 'https://physionet.org/content/ludb/1.0.1/'

def fetch(url, path):
    if not path.exists():
        with urlopen(url, timeout=60) as r:
            with path.with_suffix(path.suffix+'.part').open('wb') as f:
                while True:
                    block=r.read(1024*1024)
                    if not block: break
                    f.write(block)
        path.with_suffix(path.suffix+'.part').replace(path)
    b=path.read_bytes()
    return {'url':url,'path':str(path.relative_to(ROOT)),'bytes':len(b),
            'sha256':hashlib.sha256(b).hexdigest(),
            'checked_utc':datetime.now(timezone.utc).isoformat()}

page=RAW/'ludb_source_page.html'
manifest=[fetch(PAGE,page)]
links=re.findall(r'href=[\"\']([^\"\']+(?:\.zip|/get-zip/[^\"\']+))[\"\']',page.read_text())
links=[urljoin(PAGE,x) for x in links if 'ludb' in x or 'lobachevsky' in x]
if len(links)!=1: raise RuntimeError(f'Expected one dataset ZIP: {links}')
archive=RAW/'ludb-1.0.1.zip'
manifest.append(fetch(links[0],archive))
with zipfile.ZipFile(archive) as z:
    for info in z.infolist():
        target=(RAW/info.filename).resolve()
        if not target.is_relative_to(RAW.resolve()):
            raise ValueError('Unsafe archive member')
    z.extractall(RAW)
datasets=list(RAW.glob('*/RECORDS'))
if len(datasets)!=1: raise RuntimeError(f'Unexpected extracted datasets: {datasets}')
dataset=datasets[0].parent
checksum=dataset/'SHA256SUMS.txt'
verified=0
if checksum.exists():
    for line in checksum.read_text().splitlines():
        expected,rel=line.split(maxsplit=1)
        rel=rel.lstrip('*')
        f=dataset/rel
        if not f.exists(): raise ValueError(f'Missing official file: {rel}')
        observed=hashlib.sha256(f.read_bytes()).hexdigest()
        if observed!=expected: raise ValueError(f'Checksum mismatch: {rel}')
        verified+=1
manifest.append({'extracted_root':str(dataset.relative_to(ROOT)),
                 'official_checksums_verified':verified,
                 'record_count':len(datasets[0].read_text().splitlines())})
(ROOT/'data_manifest.json').write_text(json.dumps(manifest,indent=2))
print(json.dumps(manifest[-1]),flush=True)
```
