## Supplementary material for "Calibration and processing sensitivity of time-embedded three-dimensional electrocardiographic descriptors: a two-dataset benchmark": Research materials and reproducible analysis code: figures_revision.py

```
from pathlib import Path
import numpy as np
import pandas as pd
import matplotlib
matplotlib.use('Agg')
import matplotlib.pyplot as plt
from matplotlib.colors import LogNorm

ROOT=Path(__file__).resolve().parent.parent
R=ROOT/'estudio_3decg_revision1/results'
D=ROOT/'Biomedical Signal Processing and Control/figures'
D.mkdir(exist_ok=True)
METHODS=['FD','SG','SG0SPEC','SPEC','TIK']
COLORS=['#2166ac','#d95f02','#7570b3','#00876c','#c51b7d']
MODELS=['helix_10','helix_30','helix_60','modulated_phase','gaussian_QRS']
NAMES=['Helix 10 Hz','Helix 30 Hz','Helix 60 Hz','Modulated phase','Gaussian transient']
plt.rcParams.update({'font.family':'DejaVu Sans','font.size':10,'axes.spines.top':False,'axes.spines.right':False,'pdf.fonttype':42,'ps.fonttype':42})

def save(fig,name):
    fig.savefig(D/(name+'.pdf'),bbox_inches='tight',metadata={'Creator':None,'Producer':None,'CreationDate':None,'ModDate':None})
    fig.savefig(D/(name+'.png'),dpi=160,bbox_inches='tight')
    plt.close(fig)

def heat(ax,data,models,field,title):
    vals=data.pivot(index='model',columns='method',values=field).reindex(index=models,columns=METHODS).to_numpy()
    im=ax.imshow(np.maximum(vals,.001),cmap='YlOrRd',norm=LogNorm(.001,1100),aspect='auto')
    ax.set(xticks=range(5),xticklabels=METHODS,yticks=range(len(models)),yticklabels=[NAMES[MODELS.index(m)] for m in models])
    ax.tick_params(length=0,labelsize=9);ax.set_title(title,fontsize=10,loc='left',pad=9)
    for i in range(len(models)):
        for j in range(5):
            v=vals[i,j];s='<0.001' if v<.001 else f'{v:.3f}' if v<.01 else f'{v:.2f}' if v<100 else f'{v:.0f}'
            ax.text(j,i,s,ha='center',va='center',fontsize=8,color='white' if v>90 else '#152030')
    return im

def make():
    p=pd.read_csv(ROOT/'estudio_3decg_paper/results/planned_comparisons.csv')
    p=p[(p.metric=='abs_torsion')&p.comparison.str.contains('40_vs_')].copy()
    p.to_csv(D/'Figure1_data.csv',index=False)
    fig,ax=plt.subplots(figsize=(7.6,3.7),layout='constrained')
    methods=['FD','SG','SG0SPEC','SPEC']
    for off,cohort,color,marker in [(-.14,'LUDB','#2166ac','o'),(.14,'PTBXL','#d95f02','s')]:
        for i,m in enumerate(methods):
            r=p[(p.cohort==cohort)&(p.comparison==f'{m}40_vs_{m}150')].iloc[0]
            ax.errorbar(r.change_median,i+off,xerr=[[r.change_median-r.ci025],[r.ci975-r.change_median]],fmt=marker,color=color,capsize=3,label='PTB-XL' if cohort=='PTBXL' else cohort if i==0 else None)
            ax.text(3,i+off,f'{r.spearman:.3f}',color=color,fontsize=10,va='center',ha='left')
    handles,labels=ax.get_legend_handles_labels();seen={};[(seen.setdefault(l,h)) for h,l in zip(handles,labels)]
    ax.legend(seen.values(),seen.keys(),loc='upper left',frameon=False,ncol=2)
    ax.set(xlim=(-80,15),ylim=(-.65,3.55),yticks=range(4),yticklabels=methods,xlabel='Median paired change from 150 to 40 Hz (%)')
    ax.set_xticks([-80,-60,-40,-20,0]);ax.invert_yaxis();ax.axvline(0,color='.7',lw=.8)
    ax.text(3,-.60,'Spearman ρ',ha='left',fontsize=9);ax.grid(axis='x',alpha=.15)
    save(fig,'Figure_1')

    c=pd.read_csv(R/'extended_calibration_summary.csv');c=c[c.metric=='abs_torsion']
    fig,axes=plt.subplots(2,2,figsize=(8.6,7.5),layout='constrained',gridspec_kw={'height_ratios':[5,3]})
    data=[]
    for ax,noise,title in [(axes[0,0],0,'A  150 Hz; no noise'),(axes[0,1],.02,'B  150 Hz; 20 µV noise')]:
        z=c[(c.cutoff==150)&~c.matched&(c.noise_sd_mV==noise)];data.append(z)
        im=heat(ax,z,MODELS,'relative_rmse_percent',title)
    z=c[(c.cutoff==40)&~c.matched&(c.noise_sd_mV==0)&c.model.str.startswith('helix')];data.append(z)
    heat(axes[1,0],z,MODELS[:3],'relative_rmse_percent','C  40 Hz; unfiltered helix target')
    heat(axes[1,1],z,MODELS[:3],'filtered_relative_rmse_percent','D  40 Hz; exact filtered helix target')
    cb=fig.colorbar(im,ax=axes.ravel().tolist(),location='bottom',fraction=.035,pad=.07);cb.set_label('Relative RMSE (%)');cb.set_ticks([.001,.01,.1,1,10,100,1000])
    pd.concat(data).to_csv(D/'Figure2_data.csv',index=False);save(fig,'Figure_2')

    f=pd.read_csv(R/'operator_responses.csv');f=f[f.cutoff==150]
    fig,axes=plt.subplots(2,3,figsize=(8.6,5.8),layout='constrained',sharey=True)
    for row,matched in enumerate([False,True]):
        for col,k in enumerate([1,2,3]):
            ax=axes[row,col]
            for method,color in zip(METHODS,COLORS):
                z=f[(f.matched==matched)&(f.order==k)]
                z=z[z.method==method]
                ax.semilogy(z.frequency_Hz,np.maximum(abs(z.normalized_response),1e-8),color=color,label=method,lw=1.5)
            ax.axhline(1/np.sqrt(2),color='.4',lw=.8,ls='--')
            if matched:ax.axvline(20,color='.65',lw=.8,ls=':')
            ax.set(xlim=(0,100 if matched else 180),ylim=(.001,1.15),title=f'{"ABCDEF"[row*3+col]}  Derivative order {k}',xlabel='Frequency (Hz)')
            ax.grid(alpha=.12)
    axes[0,0].set_ylabel('Native chain\nNormalized magnitude');axes[1,0].set_ylabel('20-Hz crossing matched\nNormalized magnitude')
    fig.legend(*axes[0,0].get_legend_handles_labels(),loc='outside lower center',ncol=5,frameon=False)
    f.to_csv(D/'Figure3_data.csv',index=False);save(fig,'Figure_3')

    fig=plt.figure(figsize=(8.6,7.8),layout='constrained');gs=fig.add_gridspec(2,2,height_ratios=[1.25,1]);axes=[fig.add_subplot(gs[0,i]) for i in range(2)]
    texts=[['PTB-XL fold 10\n2,198 records; 1,904 people','One ECG per person; fixed hash ranking\n1,000 selected and included','12,188 R peaks and complete triples per lead\n12,188 mutually matched windows','Duration exclusions: 0\nEdge exclusions: 2,453','9,735 eligible QRS windows\n1,000 participants'],['LUDB\n200 records; 200 people','Complete triples: V1 1,816; V6 1,823\nIncomplete: V1 15; V6 7','1,811 mutually matched windows\nUnmatched: V1 5; V6 12','Duration exclusions: 0\nEdge exclusions: 63','1,748 eligible QRS windows\n200 participants']]
    for ax,content in zip(axes,texts):
        ax.set(xlim=(0,1),ylim=(0,1));ax.axis('off')
        for j,txt in enumerate(content):
            y=.92-.205*j
            ax.text(.5,y,txt,ha='center',va='center',fontsize=8.9,bbox=dict(boxstyle='round,pad=.55',facecolor='#edf3f7',edgecolor='#7690a4'),transform=ax.transAxes)
            if j<4:ax.annotate('',xy=(.5,y-.14),xytext=(.5,y-.06),arrowprops=dict(arrowstyle='->',color='#596b79'),xycoords='axes fraction')
    ax=fig.add_subplot(gs[1,:]);duration=pd.read_csv(R/'duration_histogram.csv')
    for cohort,color in [('LUDB','#2166ac'),('PTBXL','#d95f02')]:
        z=duration[duration.cohort==cohort].sort_values('duration_ms');ax.step(z.duration_ms,z.windows.cumsum()/z.windows.sum(),where='post',label='PTB-XL' if cohort=='PTBXL' else cohort,color=color,lw=2)
    ax.axvspan(80,160,color='#e5e9ed',zorder=0);ax.text(83,.15,'80–160 ms\nsensitivity range',fontsize=9,color='#425567')
    ax.set(xlim=(60,300),ylim=(0,1.01),xlabel='Eligible QRS-window duration (ms)',ylabel='Cumulative fraction of windows');ax.legend(frameon=False,loc='lower right');ax.grid(alpha=.15)
    duration.to_csv(D/'Figure4_duration_data.csv',index=False);pd.read_csv(R/'inclusion_flow.csv').to_csv(D/'Figure4_flow_data.csv',index=False);save(fig,'Figure_4')

    g=pd.read_csv(R/'gaussian_control_exact.csv');g=g[g.time_ms.between(3860,4140)]
    fig,axes=plt.subplots(4,1,figsize=(7.6,6.8),layout='constrained',sharex=True)
    for k,ax in enumerate(axes):
        for lead,color in [(1,'#2166ac'),(6,'#d95f02')]:
            col=f'V{lead}_mV' if k==0 else f'd{k}_V{lead}'
            ax.plot(g.time_ms-4000,g[col],label=f'Voltage {lead}',color=color)
        ax.axvspan(-80,80,color='#e5e9ed',zorder=0);ax.set_ylabel('Voltage (mV)' if k==0 else f'Order {k}\n(mV/ms$^{k}$)');ax.ticklabel_format(axis='y',style='sci',scilimits=(-2,2));ax.grid(alpha=.1)
    axes[0].legend(frameon=False,ncol=2);axes[-1].set_xlabel('Time relative to 4 s (ms)')
    g.to_csv(D/'FigureS1_data.csv',index=False);save(fig,'Figure_S1')
    fig,axes=plt.subplots(1,2,figsize=(8.6,4.4),layout='constrained');data=[]
    for ax,noise,title in [(axes[0],0,'A  20-Hz crossing matched; no noise'),(axes[1],.02,'B  20-Hz crossing matched; 20 µV noise')]:
        z=c[c.matched&(c.noise_sd_mV==noise)];data.append(z);im=heat(ax,z,MODELS,'relative_rmse_percent',title)
    cb=fig.colorbar(im,ax=axes,location='bottom',fraction=.045,pad=.07);cb.set_label('Relative RMSE against unfiltered target (%)');cb.set_ticks([.001,.01,.1,1,10,100,1000])
    pd.concat(data).to_csv(D/'FigureS2_data.csv',index=False);save(fig,'Figure_S2')

if __name__=='__main__':make()
```
