## Supplementary material for "Calibration and processing sensitivity of time-embedded three-dimensional electrocardiographic descriptors: a two-dataset benchmark": Research materials and reproducible analysis code: revision_analysis.py

```
"""Post hoc revision analyses; original study outputs are read-only inputs."""
from pathlib import Path
import argparse, hashlib, json, sys
import numpy as np
import pandas as pd
import wfdb
from scipy.signal import butter, sosfreqz, savgol_coeffs
from scipy.stats import spearmanr
from scipy.optimize import brentq

R=Path(__file__).resolve().parent; P=R.parent
sys.path.insert(0,str(P/'estudio_3decg_paper')); import analysis as old
O=R/'results'; O.mkdir(exist_ok=True)
METHODS=old.METHODS+['TIK']; METRICS=old.METRICS
SEED=20260919

def save(frame,name):
    pd.DataFrame(frame).to_csv(O/(name+'.csv'),index=False)

def sha(path): return hashlib.sha256(path.read_bytes()).hexdigest()

def check_plan():
    assert sha(R/'REVISION_PLAN.md')==json.loads((R/'revision_plan_lock.json').read_text())['sha256']

def kernel(f,method,k):
    f=np.asarray(f,dtype=float); w=2*np.pi*f/1000
    if method=='FD': return np.sinc(2*f/500)**k
    if method=='SPEC': return np.ones_like(f)
    if method=='TIK': return 1/(1+(np.sqrt(2)-1)*(f/40)**6)
    n=np.arange(-5,6)
    c=savgol_coeffs(11,3,deriv=k if method=='SG' else 0,delta=2,use='dot')
    resp=np.exp(1j*w[...,None]*2*n)@c
    if method=='SG': resp=resp/(1j*w)**k
    return np.real(resp)

def bresponse(f,cut):
    return abs(sosfreqz(butter(4,cut,fs=500,output='sos'),worN=np.atleast_1d(f),fs=500)[1])**2

def alpha(method,k):
    v=float(abs(kernel(20.,method,k))*bresponse(20,150)[0])
    a=np.log(np.sqrt(2)*v)
    assert a>=0,(method,k,a)
    return a

def crossing(fun):
    grid=np.linspace(.1,249.99,25000);v=abs(fun(grid))-1/np.sqrt(2)
    ix=np.flatnonzero(v<=0)
    assert len(ix)
    j=ix[0]
    return brentq(lambda f: float(abs(fun(np.array([f])))[0])-1/np.sqrt(2),grid[max(0,j-1)],grid[j])

def transfer():
    rows=[]; curves=[]
    frequencies=np.linspace(.1,249.9,2499)
    for cut in old.CUTS:
        for method in METHODS:
            for k in [1,2,3]:
                response=lambda f: kernel(f,method,k)*bresponse(f,cut)
                fc=crossing(response)
                rows.append(dict(method=method,cutoff=cut,order=k,matched=False,normalized_f3db_Hz=fc,
                                 gaussian_alpha=0.,noise_gain_integral=float(np.trapezoid(abs(response(frequencies)*(1j*2*np.pi*frequencies/1000)**k)**2,frequencies))))
                for f,v in zip(frequencies,response(frequencies)):
                    curves.append(dict(method=method,cutoff=cut,order=k,matched=False,frequency_Hz=f,normalized_response=v))
                if cut==150:
                    aa=alpha(method,k)
                    fun=lambda f: response(f)*np.exp(-aa*(f/20)**2)
                    matched=crossing(fun); np.testing.assert_allclose(matched,20,atol=1e-7)
                    rows.append(dict(method=method,cutoff=cut,order=k,matched=True,normalized_f3db_Hz=matched,
                                     gaussian_alpha=aa,noise_gain_integral=float(np.trapezoid(abs(fun(frequencies)*(1j*2*np.pi*frequencies/1000)**k)**2,frequencies))))
                    for f,v in zip(frequencies,fun(frequencies)):
                        curves.append(dict(method=method,cutoff=cut,order=k,matched=True,frequency_Hz=f,normalized_response=v))
    save(rows,'operator_bandwidths');save(curves,'operator_responses')
    checks=[]
    t=np.arange(4000)*2.;idx=np.arange(1000,3000)
    for f in [10.,30.,60.,100.,150.]:
        phase=2*np.pi*f*t/1000; w=2*np.pi*f/1000
        xy=np.column_stack((np.cos(phase),np.sin(phase)))
        for method in old.METHODS:
            if method=='SPEC':
                d=old.spectral(xy,periodic=True)
            elif method=='SG0SPEC':
                d=old.spectral(xy*kernel(f,method,1),periodic=True)
            else:
                d=old.derive(xy,method)
            for k in [1,2,3]:
                exact=w**k*np.column_stack((np.cos(phase+k*np.pi/2),np.sin(phase+k*np.pi/2)))
                predicted=exact*kernel(f,method,k)
                discrepancy=float(abs(d[k-1][idx]-predicted[idx]).max())
                tol=1e-10
                assert discrepancy<tol,(f,method,k,discrepancy)
                checks.append(dict(frequency=f,method=method,order=k,max_error=discrepancy,
                                   boundary_model='periodic exact harmonic' if method in ['SPEC','SG0SPEC'] else 'interior stencil'))
    save(checks,'transfer_numerical_checks')

def smooth(v,weight):
    ext=np.concatenate((v,v[-2:0:-1]),axis=0); f=np.fft.rfftfreq(len(ext),d=.002)
    ft=np.fft.rfft(ext,axis=0)
    return np.fft.irfft(ft*weight(f)[:,None],n=len(ext),axis=0)[:len(v)]

def derive(xy,method,matched=False):
    if method=='TIK':
        ext=np.concatenate((xy,xy[-2:0:-1]),axis=0);f=np.fft.rfftfreq(len(ext),d=.002);w=2*np.pi*f/1000
        ft=np.fft.rfft(ext,axis=0);ft[-1]=0
        d=tuple(np.fft.irfft(ft*kernel(f,'TIK',k)[:,None]*(1j*w[:,None])**k,n=len(ext),axis=0)[:len(xy)] for k in [1,2,3])
    else: d=old.derive(xy,method)
    if matched:
        d=tuple(smooth(v,lambda f,k=k:np.exp(-alpha(method,k)*(f/20)**2)) for k,v in enumerate(d,1))
    return d

def paired(x,y,boot=True):
    pair=pd.concat([x.rename('x'),y.rename('y')],axis=1).dropna();pair=pair[pair.y!=0]
    v=100*(pair.x/pair.y-1).to_numpy(); n=len(v)
    if not n:return dict(n=0)
    result=dict(n=n,change_median=float(np.median(v)),absolute_change_median=float(np.median(abs(v))),
                spearman=float(spearmanr(pair.x,pair.y).statistic),q25=float(np.quantile(v,.25)),q75=float(np.quantile(v,.75)))
    if boot:
        rng=np.random.default_rng(SEED); b=np.median(v[rng.integers(0,n,size=(5000,n))],axis=1)
        result.update(ci025=float(np.quantile(b,.025)),ci975=float(np.quantile(b,.975)))
    return result

def basic_tables():
    levels=[]; duration=[]; hist=[]; flow=[]; compare=[]; heuristics=[]; restricted=[]; restriction_counts=[]
    for cohort in ['LUDB','PTBXL']:
        p=pd.read_csv(old.OUT/f'{cohort}_participants.csv')
        windows=pd.read_csv(old.OUT/f'{cohort}_windows.csv'); windows['duration_ms']=2*(windows.end_sample-windows.start_sample)
        save(windows,'duration_windows_'+cohort)
        for (method,cut),group in p.groupby(['method','cutoff']):
            for metric in METRICS:
                v=group[metric].dropna()
                levels.append(dict(cohort=cohort,method=method,cutoff=cut,metric=metric,n=len(v),median=v.median(),q25=v.quantile(.25),q75=v.quantile(.75),unit='dimensionless normalized coordinates'))
        for unit,v in [('window',windows.duration_ms),('participant_median',windows.groupby('patient').duration_ms.median())]:
            duration.append(dict(cohort=cohort,unit=unit,n=len(v),minimum=v.min(),p05=v.quantile(.05),q25=v.quantile(.25),median=v.median(),q75=v.quantile(.75),p95=v.quantile(.95),maximum=v.max()))
        for dur,count in windows.duration_ms.value_counts().sort_index().items():hist.append(dict(cohort=cohort,duration_ms=dur,windows=count))
        audited=pd.read_csv(old.P1/'results/inclusion_audit.csv') if cohort=='LUDB' else pd.read_csv(old.OUT/'PTBXL_audit.csv')
        row=dict(cohort=cohort,selected=200 if cohort=='LUDB' else 1000,included=p.patient.nunique())
        row.update({col:int(audited[col].sum()) for col in ['qrs_v1','qrs_v6','incomplete_v1','incomplete_v6','matched','excluded_duration','excluded_edge','eligible','unmatched_v1','unmatched_v6']})
        if cohort=='PTBXL':
            selection=pd.read_csv(old.ROOT/'data/external_selection.csv'); totals={'R_peaks_V1':0,'R_peaks_V6':0}
            for r in selection.itertuples(index=False):
                folder=Path(r.filename_hr).parent.name
                for lead in ['V1','V6']:
                    ann=wfdb.rdann(str(old.ROOT/'data/plus/fiducial_points/ecgdeli'/folder/f'{r.ecg_id:05}_points_lead_{lead}'),'atr')
                    totals['R_peaks_'+lead]+=sum(label=='R peak' for label in ann.aux_note)
            assert totals['R_peaks_V1']==row['qrs_v1']+row['incomplete_v1']
            assert totals['R_peaks_V6']==row['qrs_v6']+row['incomplete_v6']
            row.update(totals,fold_records=2198,fold_people=1904)
        flow.append(row)
        for cut in old.CUTS:
            values=p[p.cutoff==cut].pivot(index='patient',columns='method',values='abs_torsion')
            s=float(np.sinc(2*cut/500))
            compare.append(dict(cohort=cohort,cutoff=cut,sinc_edge=s,edge_percent=100*(1/s-1),**paired(values.SPEC,values.FD)))
        pv=p.pivot(index='patient',columns=['method','cutoff'],values='abs_torsion')
        for cut in [40,100]:
            heuristic=100*((pv[('SPEC',cut)]/pv[('SPEC',150)])*np.sinc(2*cut/500)/np.sinc(2*150/500)-1)
            observed=100*(pv[('FD',cut)]/pv[('FD',150)]-1)
            heuristics.append(dict(cohort=cohort,cutoff=cut,predicted_median=heuristic.median(),observed_median=observed.median(),median_absolute_prediction_error_pp=(heuristic-observed).abs().median()))
        keep=windows[windows.duration_ms.between(80,160)]
        b=pd.read_csv(old.OUT/f'{cohort}_beats.csv').merge(keep[['patient','beat']],on=['patient','beat'],validate='many_to_one')
        q=b.groupby(['patient','method','cutoff'])[METRICS].median().reset_index()
        save(q,'common_duration_participants_'+cohort)
        restriction_counts.append(dict(cohort=cohort,windows=len(keep),participants=keep.patient.nunique(),excluded_participants=p.patient.nunique()-keep.patient.nunique()))
        for metric in METRICS:
            v=q.pivot(index='patient',columns=['method','cutoff'],values=metric)
            for method in ['FD','SPEC']:
                restricted.append(dict(cohort=cohort,metric=metric,comparison=f'{method}40_vs_{method}150',**paired(v[(method,40)],v[(method,150)])))
            restricted.append(dict(cohort=cohort,metric=metric,comparison='SPEC150_vs_FD150',**paired(v[('SPEC',150)],v[('FD',150)])))
    for name,rows in [('absolute_descriptors',levels),('duration_summary',duration),('duration_histogram',hist),('inclusion_flow',flow),('spec_fd_all_cutoffs',compare),('cutoff_edge_heuristic',heuristics),('common_duration_comparisons',restricted),('common_duration_counts',restriction_counts)]:save(rows,name)
    print('Saved absolute levels, durations, inclusion and cutoff contrasts',flush=True)

def extra_cohort(cohort):
    windows=pd.read_csv(old.OUT/f'{cohort}_windows.csv');wgroups=dict(tuple(windows.groupby('patient')))
    items=list(windows.groupby('record')) if cohort=='LUDB' else list(pd.read_csv(old.ROOT/'data/external_selection.csv').itertuples(index=False))
    rows=[]; audits=[]; differences=[]
    for count,item in enumerate(items,1):
        if cohort=='LUDB':
            record,ws=item;patient=int(ws.patient.iloc[0]);_,xy=old.prior.read(record)
        else:
            patient=int(item.patient_id);record=str(item.ecg_id);ws=wgroups[patient];xy=old.load_external(item)
        wlist=list(ws.itertuples(index=False))
        for cut in old.CUTS:
            filtered=old.prior.lowpass(xy,cut)
            configs=[('TIK',False)]+([(method,True) for method in METHODS] if cut==150 else [])
            for method,matched in configs:
                g=old.geometry(derive(filtered,method,matched))
                for w in wlist:
                    rows.append(dict(cohort=cohort,patient=patient,record=record,beat=w.beat,cutoff=cut,method=method,matched=matched,**old.measure(g,w.start_sample,w.end_sample)))
            if cut==150:
                d=old.derive(filtered,'FD');p,q,s=d
                A=np.sum(q*q,axis=1);C=p[:,0]*q[:,1]-p[:,1]*q[:,0];G=(1+np.sum(p*p,axis=1))**1.5
                K=np.sqrt(p[:,0]**2+q[:,1]**2+C*C)/G
                proxy=abs(p[:,0])/G; limit=np.sqrt(q[:,1]**2+C*C)/G
                classic=np.sqrt(A+C*C)/G
                transformed=np.sqrt(p[:,0]**2/1e6+q[:,1]**2+C*C)/G
                np.testing.assert_allclose(transformed,old.geometry(d,reparam=1000)['almost_curvature'],rtol=1e-10,atol=1e-14)
                base=old.geometry(d);np.testing.assert_allclose(K,base['almost_curvature'],rtol=1e-12,atol=1e-15)
                slope_correction=np.divide(C*C,A,out=np.full(len(A),np.nan),where=A>0)
                share=np.divide(p[:,0]**2,p[:,0]**2+q[:,1]**2+C*C,out=np.zeros(len(A)),where=(p[:,0]**2+q[:,1]**2+C*C)>0)
                for w in wlist:
                    ix=slice(w.start_sample,w.end_sample+1);T=(w.end_sample-w.start_sample)*2
                    avg=lambda v:float(np.trapezoid(v[ix],dx=2)/T)
                    audits.append(dict(cohort=cohort,patient=patient,beat=w.beat,K=avg(K),slope_proxy=avg(proxy),K_limit=avg(limit),classical_curvature=avg(classic),K_reparam=avg(transformed),first_term_share=avg(share),C2_over_A_mean=avg(slope_correction),C2_over_A_max=float(np.nanmax(slope_correction[ix]))))
        if count%100==0:print('Revision',cohort,count,'of',len(items),flush=True)
    frame=pd.DataFrame(rows);save(frame,'additional_beats_'+cohort)
    part=frame.groupby(['cohort','patient','method','cutoff','matched'])[METRICS].median().reset_index();save(part,'additional_participants_'+cohort)
    audit=pd.DataFrame(audits);save(audit,'index_decomposition_beats_'+cohort)
    save(audit.groupby(['cohort','patient']).median(numeric_only=True).reset_index(),'index_decomposition_participants_'+cohort)

def gaussian_control(t):
    xy=np.zeros((len(t),2));d=[np.zeros_like(xy) for _ in range(3)]
    components=[[( -.15,3975.,12.),(1.,4000.,8.),(-.25,4035.,15.)],[(.10,3985.,14.),(.8,4012.,10.),(-.35,4045.,16.)]]
    for lead,items in enumerate(components):
        for amp,center,sigma in items:
            z=(t-center)/sigma;g=amp*np.exp(-.5*z*z);xy[:,lead]+=g
            d[0][:,lead]+=-z*g/sigma
            d[1][:,lead]+=(z*z-1)*g/sigma**2
            d[2][:,lead]+=(3*z-z**3)*g/sigma**3
    return xy,tuple(d)

def calibration():
    rng=np.random.default_rng(20260917);t=np.arange(4000)*2.;rows=[];targetrows=[]
    models=['helix_10','helix_30','helix_60','modulated_phase','gaussian_QRS']
    for model in models:
        xy,d=gaussian_control(t) if model=='gaussian_QRS' else old.truth_signal(model,t)
        lo,hi=(1960,2040) if model=='gaussian_QRS' else (1000,3000)
        truth=old.measure(old.geometry(d),lo,hi)
        assert all(np.isfinite(truth[m]) and truth[m]>0 for m in METRICS),(model,truth)
        if model=='gaussian_QRS':
            example={'time_ms':t,'V1_mV':xy[:,0],'V6_mV':xy[:,1]}
            for k in range(3):
                for lead in range(2):example[f'd{k+1}_V{1 if lead==0 else 6}']=d[k][:,lead]
            save(example,'gaussian_control_exact')
        for cut in old.CUTS:
            filtered={m:np.nan for m in METRICS}
            if model.startswith('helix'):
                f=float(model.split('_')[1]);amp=float(bresponse(f,cut)[0]);filtered=old.measure(old.geometry(tuple(v*amp for v in d)),lo,hi)
            for m in METRICS:targetrows.append(dict(model=model,cutoff=cut,metric=m,truth=truth[m],filtered_truth=filtered[m],start_sample=lo,end_sample=hi))
        for noise in [0.,.005,.02]:
            for realization in range(1 if noise==0 else 100):
                noisy=xy+rng.normal(0,noise,xy.shape)
                for cut in old.CUTS:
                    fxy=old.prior.lowpass(noisy,cut)
                    for matched in ([False,True] if cut==150 else [False]):
                        for method in METHODS:
                            val=old.measure(old.geometry(derive(fxy,method,matched)),lo,hi)
                            for m in METRICS:rows.append(dict(model=model,noise_sd_mV=noise,realization=realization,cutoff=cut,method=method,matched=matched,metric=m,estimate=val[m],truth=truth[m]))
        print('Extended calibration',model,'complete',flush=True)
    df=pd.DataFrame(rows);targets=pd.DataFrame(targetrows)
    df=df.merge(targets[['model','cutoff','metric','filtered_truth']],on=['model','cutoff','metric'],validate='many_to_one')
    save(df,'extended_calibration_estimates');save(targets,'extended_calibration_targets')
    previous=pd.read_csv(old.OUT/'calibration_estimates.csv')
    legacy=df[(df.model!='gaussian_QRS')&(df.method!='TIK')&~df.matched]
    keys=['model','noise_sd_mV','realization','cutoff','method','metric']
    pair=legacy.merge(previous,on=keys,validate='one_to_one',suffixes=('_new','_old'))
    assert len(pair)==len(previous)
    np.testing.assert_allclose(pair.estimate_new,pair.estimate_old,rtol=1e-12,atol=1e-14,equal_nan=True)
    (O/'legacy_calibration_reproduction.json').write_text(json.dumps({'rows':len(pair),'max_absolute_difference':float(abs(pair.estimate_new-pair.estimate_old).max())},indent=2))
    summaries=[]
    for keys,group in df.groupby(['model','noise_sd_mV','cutoff','method','matched','metric']):
        row=dict(zip(['model','noise_sd_mV','cutoff','method','matched','metric'],keys));err=100*(group.estimate/group.truth-1)
        row.update(n=len(group),nonfinite=int((~np.isfinite(group.estimate)).sum()),relative_bias_percent=err.mean(),relative_rmse_percent=np.sqrt((err*err).mean()))
        if group.filtered_truth.notna().all():
            errf=100*(group.estimate/group.filtered_truth-1)
            row.update(filtered_relative_bias_percent=errf.mean(),filtered_relative_rmse_percent=np.sqrt((errf*errf).mean()))
        summaries.append(row)
    save(summaries,'extended_calibration_summary')

def summarize_extra():
    comparisons=[];descriptions=[];indices=[]
    for cohort in ['LUDB','PTBXL']:
        p=pd.read_csv(O/f'additional_participants_{cohort}.csv')
        legacy=pd.read_csv(old.OUT/f'{cohort}_participants.csv');legacy['matched']=False
        allp=pd.concat([legacy,p],ignore_index=True)
        for (method,cut,match),v in allp.groupby(['method','cutoff','matched']):
            for m in METRICS:
                a=v[m].dropna();descriptions.append(dict(cohort=cohort,method=method,cutoff=cut,matched=match,metric=m,n=len(a),median=a.median(),q25=a.quantile(.25),q75=a.quantile(.75)))
        for metric in METRICS:
            pv=allp.pivot(index='patient',columns=['method','cutoff','matched'],values=metric)
            for method in METHODS:
                if method!='FD':comparisons.append(dict(cohort=cohort,metric=metric,comparison=method+'_vs_FD_matched20',**paired(pv[(method,150,True)],pv[('FD',150,True)])))
                comparisons.append(dict(cohort=cohort,metric=metric,comparison=method+'_matched20_vs_native150',**paired(pv[(method,150,True)],pv[(method,150,False)])))
            for cut in old.CUTS:
                comparisons.append(dict(cohort=cohort,metric=metric,comparison=f'TIK{cut}_vs_SPEC{cut}',**paired(pv[('TIK',cut,False)],pv[('SPEC',cut,False)])))
        g=pd.read_csv(O/f'index_decomposition_participants_{cohort}.csv').set_index('patient')
        for name,a,b in [('slope_proxy_vs_K','slope_proxy','K'),('K_limit_vs_curvature','K_limit','classical_curvature'),('K_reparam_vs_K_limit','K_reparam','K_limit')]:
            indices.append(dict(cohort=cohort,comparison=name,**paired(g[a],g[b])))
        for metric in ['first_term_share','C2_over_A_mean','C2_over_A_max']:
            descriptions.append(dict(cohort=cohort,method='FD',cutoff=150,matched=False,metric=metric,n=len(g),median=g[metric].median(),q25=g[metric].quantile(.25),q75=g[metric].quantile(.75)))
    save(comparisons,'additional_comparisons');save(descriptions,'expanded_absolute_descriptors');save(indices,'index_decomposition_comparisons')

if __name__=='__main__':
    ap=argparse.ArgumentParser();ap.add_argument('task',choices=['transfer','tables','LUDB','PTBXL','calibration','summarize']);args=ap.parse_args();check_plan()
    if args.task=='transfer':transfer()
    elif args.task=='tables':basic_tables()
    elif args.task in ['LUDB','PTBXL']:extra_cohort(args.task)
    elif args.task=='calibration':calibration()
    else:summarize_extra()
```
