## Supplementary material for "Calibration and processing sensitivity of time-embedded three-dimensional electrocardiographic descriptors: a two-dataset benchmark": Research materials and reproducible analysis code: REVISION_PLAN.md

# Revision analyses following the supplied review

These are explicitly post hoc analyses prompted by review of the completed manuscript. The original protocol, selections, software and 88 planned comparisons remain unchanged. No new preregistration or outcome-blinded design is claimed.

1. Derive and independently verify the interior frequency responses of FD, SG, SPEC and SG0SPEC by derivative order. Report the first -3 dB crossing of response normalized by the ideal derivative, after the double-pass Butterworth filter. A cutoff-edge sinusoidal calculation is a heuristic for multicomponent ECGs, not a universal correction or proof of patient-level error.
2. Add a coherent Tikhonov spectral estimator with a third-derivative quadratic penalty: W(f) = 1/[1 + (sqrt(2)-1)(f/40 Hz)^6]. Use the same W for derivative orders 1–3. The 40-Hz regularization setting is fixed, not optimized on either cohort or the control outcomes. Evaluate at the original three nominal Butterworth cutoffs.
3. At nominal Butterworth 150 Hz, match the first -3 dB crossing of each normalized derivative response to 20 Hz using an additional Gaussian frequency taper for each derivative order. The setting is deliberately below the intrinsic crossings of all operators. This matches one bandwidth criterion, not the entire response; derivative-specific tapers need not represent derivatives of one common curve. Apply identical boundary extension conventions and report these limitations.
4. Re-express the printed almost-curvature formula with z=t, quantify its first-derivative contribution and its approximation by a normalized V1 slope, and derive its exact reparametrization limit. Do not label the formula a typographical error: the source explicitly describes lowering a derivative order. Do not confuse the limit with classical curvature.
5. Report participant-level median and IQR of all four descriptors for each cohort/operator/cutoff, with dimensionless normalized-coordinate conventions. Add the per-window duration distributions and an 80–160 ms inclusive common-duration analysis retaining eligible windows and reporting missing participants.
6. Reconstruct the inclusion flow from original annotations and saved audits, retaining lead-specific counts. The original R-peak and incomplete counts will be checked directly from files for PTB-XL. Distinguish participant exclusion from window exclusion.
7. Report unfiltered and exact post-filter targets for all original helical controls and all three cutoffs, including 5-microvolt noise. Add a transient two-coordinate Gaussian-mixture pulse with exact derivatives, evaluated on a fixed 160-ms window centered at 4 s within an 8-s, 500-Hz record. This is a mathematical QRS-like stress test, not a validated physiological simulator.
8. Use the original noise seed 20260917 and shared noise realizations (one noiseless and 100 each at 5 and 20 microvolts). Recreate the first four original controls in their original sequence and compare all legacy estimates numerically; append the transient control afterwards. The expanded calibration comprises 1,005 signal/noise realizations, distinct from the original 804.
9. Additional paired summaries use participant medians, descriptive 5,000-resample percentile bootstrap intervals with fixed seed 20260919, and no multiplicity-based method selection. Report all added conditions, including unfavorable results.
10. Verify source bibliography against primary publisher metadata. Disclose the author's authorship of the prior works. Keep the already requested omission of the manuscript-assistance declaration; do not invent affiliation, funding, other competing interests, a repository DOI, or completed submission.

The revision will preserve the distinction between exact algebra, finite-sample numerical checks, empirical ECG comparisons, and unverified causal interpretations.
