## Supplementary material for "Calibration and processing sensitivity of time-embedded three-dimensional electrocardiographic descriptors: a two-dataset benchmark": Research materials and reproducible analysis code: verify_revision.py

```
from pathlib import Path
import json, hashlib, zipfile
import numpy as np
import pandas as pd
from scipy.special import eval_hermitenorm
import revision_analysis as a

out = {}
a.check_plan()
t = np.arange(4000)*2.
xy, ds = a.gaussian_control(t)
independent = [np.zeros_like(xy) for _ in range(4)]
components = [[(-.15,3975,12),(1,4000,8),(-.25,4035,15)],[(.10,3985,14),(.8,4012,10),(-.35,4045,16)]]
for lead, group in enumerate(components):
    for amp, center, sigma in group:
        z = (t-center)/sigma
        for k in range(4):
            independent[k][:,lead] += amp*(-1)**k*eval_hermitenorm(k,z)*np.exp(-z*z/2)/sigma**k
err = max(float(abs(x-y).max()) for x,y in zip([xy,*ds],independent))
assert err < 1e-14
out['Gaussian_Hermite_max_absolute_error'] = err

rng = np.random.default_rng(1824)
y = rng.normal(size=(32,2)); ext = np.concatenate([y,y[-2:0:-1]])
n = len(ext); omega=2*np.pi*np.fft.fftfreq(n,d=2.)
nyq = (-1.)**np.arange(n); projected=ext-np.outer(nyq,nyq@ext/n)
operators=[]
for k in [1,2,3]:
    multip=(1j*omega)**k; multip[n//2]=0
    operators.append(np.fft.ifft(multip[:,None]*np.fft.fft(np.eye(n),axis=0),axis=0).real)
lam=(np.sqrt(2)-1)/(2*np.pi*.040)**6
solution=np.linalg.solve(np.eye(n)+lam*operators[2].T@operators[2],projected)
direct=[(op@solution)[:32] for op in operators]
err=max(float(abs(d-v).max()) for d,v in zip(direct,a.derive(y,'TIK')))
assert err<1e-10
out['Tikhonov_dense_normal_equations_max_absolute_error']=err

est=pd.read_csv(a.O/'extended_calibration_estimates.csv')
rows=est[(est.model.str.startswith('helix'))&(est.noise_sd_mV==0)&(est.method=='FD')&~est.matched&(est.metric=='abs_torsion')].copy()
errors=[]
for r in rows.itertuples():
    f=float(r.model.split('_')[1]); omega=2*np.pi*f/1000; s=np.sin(2*omega)/(2*omega)
    radius=float(a.bresponse(f,r.cutoff)[0]); exact=s*omega/(1+radius**2*s*s*omega**2)
    errors.append(abs(r.estimate-exact))
assert max(errors)<1e-9
out['FD_helix_transfer_max_absolute_error']=max(errors)
assert len(est)==1005*20*4
summary=pd.read_csv(a.O/'extended_calibration_summary.csv')
assert len(summary)==1200 and summary.nonfinite.sum()==0
out['calibration_realizations']=1005
out['calibration_scalar_estimates']=len(est)
out['calibration_aggregate_rows']=len(summary)
out['calibration_nonfinite_estimates']=int(summary.nonfinite.sum())

invalid_rows=[]; common_rows=[]
for cohort,n in [('LUDB',200),('PTBXL',1000)]:
    beats=pd.read_csv(a.O/f'additional_beats_{cohort}.csv')
    parts=pd.read_csv(a.O/f'additional_participants_{cohort}.csv')
    assert not beats.duplicated(['patient','beat','cutoff','method','matched']).any()
    assert parts.patient.nunique()==n and len(parts)==n*8
    assert not np.isinf(beats[a.METRICS].to_numpy()).any()
    for key,g in beats.groupby(['method','cutoff','matched']):
        invalid_rows.append(dict(cohort=cohort,method=key[0],cutoff=key[1],matched=key[2],rows=len(g),undefined_abs_torsion=int(g.abs_torsion.isna().sum()),affected_participants=g[g.abs_torsion.isna()].patient.nunique()))
    recalc=beats.groupby(['patient','method','cutoff','matched'])[a.METRICS].apply(lambda v:pd.Series(np.nanmedian(v.to_numpy(),axis=0),index=a.METRICS))
    saved=parts.set_index(['patient','method','cutoff','matched'])[a.METRICS].sort_index()
    np.testing.assert_allclose(recalc.sort_index(),saved,rtol=1e-12,atol=1e-14)
    out[cohort+'_additional_window_rows']=len(beats)
    out[cohort+'_additional_participant_rows']=len(parts)
    mat=beats[beats.matched].pivot(index=['patient','beat'],columns='method',values='abs_torsion')
    common=mat.dropna(); reduced=common.groupby('patient').median()
    assert len(reduced)==n
    for method in ['SG','SPEC','SG0SPEC','TIK']:
        common_rows.append(dict(cohort=cohort,method=method,retained_windows=len(common),excluded_windows=len(mat)-len(common),**a.paired(reduced[method],reduced.FD)))

pd.DataFrame(invalid_rows).to_csv(a.O/'additional_invalid_counts.csv',index=False)
pd.DataFrame(common_rows).to_csv(a.O/'matched_common_valid_comparisons.csv',index=False)

band=pd.read_csv(a.O/'operator_bandwidths.csv')
np.testing.assert_allclose(band[band.matched].normalized_f3db_Hz,20,atol=1e-7)
out['matched_bandwidth_conditions']=len(band[band.matched])
out['matched_bandwidth_max_error_Hz']=float(abs(band[band.matched].normalized_f3db_Hz-20).max())

p,q,c=2.,2.,0.
G=(1+p*p)**1.5
assert np.sqrt(c*c)/G==0 and abs(q)/G>0
out['almost_curvature_limit_is_not_classical_counterexample']='x(t)=t^2, y(t)=0 at t=1: limit 0; curvature 2/(5 sqrt(5))'
out['legacy_reproduction']=json.loads((a.O/'legacy_calibration_reproduction.json').read_text())
out['status']='passed'
(a.O/'independent_verification.json').write_text(json.dumps(out,indent=2))
print(json.dumps(out,indent=2))
```
