## Supplementary material for "Calibration and processing sensitivity of time-embedded three-dimensional electrocardiographic descriptors: a two-dataset benchmark": Research materials and reproducible analysis code: audit_revision2.py

```
from pathlib import Path
import sys,json,hashlib
import numpy as np
import pandas as pd
import wfdb
R=Path(__file__).resolve().parent;ROOT=R.parent;O=R/'results'
sys.path.insert(0,str(ROOT/'estudio_3decg_revision1'))
import revision_analysis as a

def triples(path,lead,cohort):
    raw=wfdb.rdann(str(path),lead)
    if cohort=='LUDB':
        peaks=[j for j,s in enumerate(raw.symbol) if s=='N'];out=[]
        for j in peaks:
            if j>0 and j+1<len(raw.symbol) and raw.symbol[j-1]=='(' and raw.symbol[j+1]==')':out.append(tuple(map(int,raw.sample[j-1:j+2])))
    else:
        assert raw.fs==500
        labels=np.asarray(raw.aux_note);values=np.asarray(raw.sample);peaks=values[labels=='R peak'];out=[]
        ons=values[labels=='QRS onset'];offs=values[labels=='QRS offset']
        for i,p in enumerate(peaks):
            lo=peaks[i-1] if i else -1;hi=peaks[i+1] if i+1<len(peaks) else 5001
            starts=ons[(ons>lo)&(ons<=p)];ends=offs[(offs>=p)&(offs<hi)]
            if len(starts)==len(ends)==1:out.append((int(starts[0]),int(p),int(ends[0])))
    arr=np.asarray(out).reshape(-1,3)
    return arr,raw

def borders():
    records=[];candidates=[];peaksrows=[];leadrows=[];checks={}
    for cohort in ['LUDB','PTBXL']:
        saved=pd.read_csv(a.old.OUT/f'{cohort}_windows.csv')
        items=sorted(saved.record.unique()) if cohort=='LUDB' else list(pd.read_csv(a.old.ROOT/'data/external_selection.csv').itertuples(index=False))
        for item in items:
            if cohort=='LUDB':
                record=str(item);pid=int(record.split('/')[-1]);paths=[(a.old.prior.DATA/record,l) for l in ['v1','v6']]
            else:
                record=str(item.ecg_id);pid=int(item.patient_id);folder=Path(item.filename_hr).parent.name
                paths=[(a.old.ROOT/'data/plus/fiducial_points/ecgdeli'/folder/f'{item.ecg_id:05}_points_lead_{l}','atr') for l in ['V1','V6']]
            arrays=[]
            for lname,(path,ext) in zip(['V1','V6'],paths):
                ar,raw=triples(path,ext,cohort);arrays.append(ar)
                rr=np.diff(ar[:,1])/500
                leadrows.append(dict(cohort=cohort,record=record,patient=pid,lead=lname,complete_qrs=len(ar),first_any_annotation_s=float(raw.sample.min()/500),last_any_annotation_s=float(raw.sample.max()/500),first_qrs_onset_s=ar[0,0]/500,last_qrs_offset_s=ar[-1,2]/500,qrs_landmark_envelope_s=(ar[-1,2]-ar[0,0])/500,first_qrs_peak_s=ar[0,1]/500,last_qrs_peak_s=ar[-1,1]/500,median_RR_s=float(np.median(rr)),peak_envelope_plus_median_RR_s=(ar[-1,1]-ar[0,1])/500+np.median(rr)))
                for onset,peak,offset in ar:peaksrows.append(dict(cohort=cohort,record=record,patient=pid,lead=lname,onset=int(onset),peak=int(peak),offset=int(offset),peak_second=int(peak//500)))
            left,right=arrays;keep=[];matched=[]
            for i,one in enumerate(left):
                j=min(range(len(right)),key=lambda j:abs(int(one[1])-int(right[j,1])))
                back=min(range(len(left)),key=lambda k:abs(int(right[j,1])-int(left[k,1])))
                if back!=i or abs(int(one[1])-int(right[j,1]))>50:continue
                lo=int(min(one[0],right[j,0]));hi=int(max(one[2],right[j,2]))
                valid_duration=20<=hi-lo<=150;leftbad=lo<500;rightbad=hi>4499
                selected=valid_duration and not(leftbad or rightbad)
                row=dict(cohort=cohort,record=record,patient=pid,start_sample=lo,end_sample=hi,v1_peak=int(one[1]),v6_peak=int(right[j,1]),valid_duration=valid_duration,left_guard_fail=leftbad,right_guard_fail=rightbad,eligible=selected)
                candidates.append(row);matched.append(row)
                if selected:keep.append((lo,hi,int(one[1]),int(right[j,1])))
            prior=saved[saved.patient==pid][['start_sample','end_sample','v1_peak','v6_peak']].to_numpy(dtype=int)
            assert np.array_equal(np.asarray(keep),prior),(cohort,record)
            peaksidentical=len(left)==len(right) and np.array_equal(left[:,1],right[:,1])
            records.append(dict(cohort=cohort,record=record,patient=pid,matched=len(matched),eligible=len(keep),left_excluded=sum(r['left_guard_fail'] and r['valid_duration'] for r in matched),right_excluded=sum(r['right_guard_fail'] and r['valid_duration'] for r in matched),duration_excluded=sum(not r['valid_duration'] for r in matched),peaks_identical=peaksidentical))
        assert saved.start_sample.min()>=500 and saved.end_sample.max()<=4499
        checks[cohort]={'all_saved_windows_match_independent_annotation_reconstruction':True,'eligible':len(saved),'minimum_start_sample':int(saved.start_sample.min()),'maximum_end_sample':int(saved.end_sample.max()),'guard_violations':0}
    for name,rows in [('annotation_coverage',leadrows),('guard_records',records),('guard_candidates',candidates),('annotated_qrs_landmarks',peaksrows)]:pd.DataFrame(rows).to_csv(O/(name+'.csv'),index=False)
    lead=pd.DataFrame(leadrows);summary=[]
    for (c,l),v in lead.groupby(['cohort','lead']):
        for field in ['first_any_annotation_s','last_any_annotation_s','first_qrs_onset_s','last_qrs_offset_s','qrs_landmark_envelope_s','first_qrs_peak_s','last_qrs_peak_s','median_RR_s','peak_envelope_plus_median_RR_s']:
            x=v[field];summary.append(dict(cohort=c,lead=l,measure=field,n=len(x),minimum=x.min(),q25=x.quantile(.25),median=x.median(),q75=x.quantile(.75),maximum=x.max()))
    pd.DataFrame(summary).to_csv(O/'annotation_coverage_summary.csv',index=False)
    pd.DataFrame(peaksrows).groupby(['cohort','lead','peak_second']).size().rename('count').reset_index().to_csv(O/'annotation_time_histogram.csv',index=False)
    for c,g in pd.DataFrame(records).groupby('cohort'):
        checks[c].update({k:int(g[k].sum()) for k in ['matched','left_excluded','right_excluded','duration_excluded','peaks_identical']})
    (O/'guard_verification.json').write_text(json.dumps(checks,indent=2));print(json.dumps(checks,indent=2))

def decomposition():
    rows=[];summary=[]
    for c in ['LUDB','PTBXL']:
        b=pd.read_csv(a.O/f'index_decomposition_beats_{c}.csv')
        b['window_ratio_with_G']=b.K_limit/b.K
        b['window_ratio_reparam']=b.K_reparam/b.K
        p=b.groupby('patient').median(numeric_only=True)
        p['ratio_of_participant_medians']=p.K_limit/p.K
        p['reparam_ratio_of_participant_medians']=p.K_reparam/p.K
        p['slope_ratio_of_participant_medians']=p.slope_proxy/p.K
        p['cohort']=c;rows.append(p.reset_index())
        for field in ['first_term_share','window_ratio_with_G','ratio_of_participant_medians','reparam_ratio_of_participant_medians','slope_ratio_of_participant_medians']:
            v=p[field];summary.append(dict(cohort=c,measure=field,n=len(v),median=v.median(),q25=v.quantile(.25),q75=v.quantile(.75)))
    pd.concat(rows).to_csv(O/'aggregation_decomposition_participants.csv',index=False)
    pd.DataFrame(summary).to_csv(O/'aggregation_decomposition_summary.csv',index=False)
    table=[]
    for f in [20,30,40]:
        for method in ['FD','SG']:table.append(dict(frequency_Hz=f,method=method,g2=float(a.kernel(f,method,2)),g3=float(a.kernel(f,method,3)),g3_over_g2=float(a.kernel(f,method,3)/a.kernel(f,method,2))))
    pd.DataFrame(table).to_csv(O/'derivative_ratio_examples.csv',index=False)
    cal=pd.read_csv(a.O/'extended_calibration_summary.csv');cal=cal[cal.matched&(cal.metric=='abs_torsion')&(cal.noise_sd_mV.isin([0,.02]))]
    z=cal.pivot(index=['model','method'],columns='noise_sd_mV',values='relative_rmse_percent').reset_index()
    z.columns=['model','method','rmse_noiseless','rmse_noise20'];z['delta_rmse_pp']=z.rmse_noise20-z.rmse_noiseless
    z.to_csv(O/'matched_panel_difference.csv',index=False)
    cand=pd.read_csv(O/'guard_candidates.csv');diff=[]
    for c,g in cand.groupby('cohort'):
        v=2*(g.v1_peak-g.v6_peak).abs()
        diff.append(dict(cohort=c,matched=len(g),zero_difference=int((v==0).sum()),median_ms=v.median(),q25_ms=v.quantile(.25),q75_ms=v.quantile(.75),maximum_ms=v.max(),over_100_ms=int((v>100).sum())))
    pd.DataFrame(diff).to_csv(O/'cross_lead_peak_differences.csv',index=False)
    t=np.arange(4000)*2.;xy,d=a.gaussian_control(t);truth=a.old.geometry(d)['abs_torsion'];idx=slice(1960,2041)
    target_area=np.trapezoid(truth[idx],dx=2.);bias=[]
    for method in ['FD','SG','SPEC']:
        estimated=a.old.geometry(a.derive(a.old.prior.lowpass(xy,150),method))['abs_torsion'];e=estimated[idx]-truth[idx]
        assert np.isfinite(e).all()
        pos=100*np.trapezoid(np.maximum(e,0),dx=2.)/target_area
        neg=100*np.trapezoid(np.minimum(e,0),dx=2.)/target_area
        bias.append(dict(method=method,positive_error_contribution_percent=pos,negative_error_contribution_percent=neg,net_bias_percent=pos+neg,integrated_absolute_error_percent=pos-neg))
    pd.DataFrame(bias).to_csv(O/'gaussian_error_cancellation.csv',index=False)

if __name__=='__main__':
    if not (O/'guard_verification.json').exists():borders()
    decomposition()
```
