## Supplementary material for "Calibration and processing sensitivity of time-embedded three-dimensional electrocardiographic descriptors: a two-dataset benchmark": Research materials and reproducible analysis code: AUDIT_SCOPE.md

# Second-review audit scope

These are post hoc checks prompted by the second supplied review. No preregistration is claimed. Original input data, processing settings, QRS selections and clinical descriptor estimates are preserved. The second-review audit re-parses the supplied annotations and reconstructs windows independently, reports annotation-landmark envelopes rather than an assumed continuous coverage duration, compares aggregation routes for the printed index, and separates positive/negative local torsion error in the existing Gaussian control. Existing matched calibration outputs are inspected at full precision. The known historical author interest is retained. Author-specific facts are incorporated only as confirmed.
