## Supplementary material for "Calibration and processing sensitivity of time-embedded three-dimensional electrocardiographic descriptors: a two-dataset benchmark": Research materials and reproducible analysis code: matching_sensitivity.py

```
from pathlib import Path
import hashlib,json
import numpy as np
import pandas as pd
ROOT=Path(__file__).resolve().parent.parent;R=Path(__file__).resolve().parent;O=R/'results';O.mkdir(exist_ok=True)
source=ROOT/'estudio_3decg_revision2/results'
c=pd.read_csv(source/'guard_candidates.csv');a=pd.read_csv(source/'annotated_qrs_landmarks.csv')
assert not a.duplicated(['cohort','record','patient','lead','peak']).any()
# Reconstruct mutual nearest neighbors from the saved complete annotation triples.
rows=[]
for (cohort,record,patient),g in a.groupby(['cohort','record','patient'],sort=False):
    x=g[g.lead=='V1'].sort_values('peak');y=g[g.lead=='V6'].sort_values('peak')
    dist=abs(x.peak.to_numpy()[:,None]-y.peak.to_numpy()[None,:])
    j=dist.argmin(axis=1);i=dist.argmin(axis=0)
    for ix,jy in enumerate(j):
        if i[jy]!=ix:continue
        left=x.iloc[ix];right=y.iloc[jy];lo=min(left.onset,right.onset);hi=max(left.offset,right.offset)
        if dist[ix,jy]>50:continue
        rows.append(dict(cohort=cohort,record=record,patient=int(patient),v1_peak=int(left.peak),v6_peak=int(right.peak),start_sample=int(lo),end_sample=int(hi),eligible=bool(20<=hi-lo<=150 and lo>=500 and hi<=4499),separation_ms=int(2*dist[ix,jy]),v1_duration_ms=int(2*(left.offset-left.onset)),v6_duration_ms=int(2*(right.offset-right.onset)),union_duration_ms=int(2*(hi-lo)),union_excess_over_longer_lead_ms=int(2*((hi-lo)-max(left.offset-left.onset,right.offset-right.onset))),onset_difference_ms=int(2*abs(left.onset-right.onset)),offset_difference_ms=int(2*abs(left.offset-right.offset))))
d=pd.DataFrame(rows);keys=['cohort','record','patient','v1_peak','v6_peak'];cols=keys+['start_sample','end_sample','eligible']
pd.testing.assert_frame_equal(d[cols].sort_values(keys).reset_index(drop=True),c[cols].sort_values(keys).reset_index(drop=True),check_dtype=False)
d['retained_at_80_ms']=d.separation_ms<=80
d.to_csv(O/'matching_tolerance_candidates.csv',index=False)
summary=[]
for cohort,g in d.groupby('cohort'):
    v=g[g.eligible];new=v[v.retained_at_80_ms]
    for tol in [100,80]:
        p=g[g.separation_ms<=tol];w=p[p.eligible]
        summary.append(dict(cohort=cohort,tolerance_ms=tol,matched_pairs=len(p),pairs_lost_from_100_ms=len(g)-len(p),eligible_windows=len(w),windows_lost_from_100_ms=len(v)-len(w),percent_eligible_windows_lost=100*(len(v)-len(w))/len(v),participants=w.patient.nunique(),participants_lost_from_100_ms=v.patient.nunique()-w.patient.nunique(),participants_with_any_window_removed=v.loc[v.separation_ms>tol,'patient'].nunique()))
pd.DataFrame(summary).to_csv(O/'matching_tolerance_summary.csv',index=False)
fields=['separation_ms','v1_duration_ms','v6_duration_ms','union_duration_ms','union_excess_over_longer_lead_ms','onset_difference_ms','offset_difference_ms']
stats=[]
for (cohort,eligible),g in d.groupby(['cohort','eligible']):
    if not eligible:continue
    for f in fields:
        v=g[f];stats.append(dict(cohort=cohort,population='eligible_windows',measure=f,n=len(g),median=float(v.median()),q25=float(v.quantile(.25)),q75=float(v.quantile(.75)),minimum=int(v.min()),maximum=int(v.max())))
pd.DataFrame(stats).to_csv(O/'lead_duration_decomposition_summary.csv',index=False)
# Source hashes make the input version checkable without exporting machine-specific paths.
provenance={str(p.relative_to(ROOT)):hashlib.sha256(p.read_bytes()).hexdigest() for p in [source/'guard_candidates.csv',source/'annotated_qrs_landmarks.csv']}
(O/'matching_sensitivity_verification.json').write_text(json.dumps({'independent_matching_reconstruction_equal_to_saved_candidates':True,'source_sha256':provenance,'sampling_frequency_Hz':500,'original_tolerance_samples':50,'sensitivity_tolerance_samples':40,'original_window_rules_preserved':True,'descriptor_estimates_recomputed':False,'interpretation':'Post hoc count sensitivity and descriptive duration decomposition; no causal separation of cohort and annotation method.'},indent=2))
print(pd.DataFrame(summary).to_string(index=False))
print(pd.DataFrame(stats)[['cohort','measure','median','q25','q75']].to_string(index=False))
```
