## Supplementary material for "Calibration and processing sensitivity of time-embedded three-dimensional electrocardiographic descriptors: a two-dataset benchmark": Research materials and reproducible analysis code: verify_aggregation.py

```
from pathlib import Path
import numpy as np
import pandas as pd
ROOT=Path(__file__).resolve().parent.parent
rows=[]
for cohort in ['LUDB','PTBXL']:
    frame=pd.read_csv(ROOT/f'estudio_3decg_paper/results/{cohort}_participants.csv')
    expected=pd.read_csv(ROOT/'estudio_3decg_revision1/results/spec_fd_all_cutoffs.csv')
    for cutoff in [40,100,150]:
        paired=frame[frame.cutoff.eq(cutoff)].pivot(index='patient',columns='method',values='abs_torsion')
        median=float(np.median(100*(paired.SPEC-paired.FD)/paired.FD))
        reference=expected[(expected.cohort==cohort)&(expected.cutoff==cutoff)].change_median.item()
        assert np.isclose(median,reference,rtol=0,atol=1e-10)
        rows.append(dict(cohort=cohort,cutoff=cutoff,n=len(paired),median_paired_percent=median,ratio_of_medians_percent=100*(paired.SPEC.median()/paired.FD.median()-1)))
result=pd.DataFrame(rows)
out=ROOT/'estudio_3decg_revision4/results';out.mkdir(parents=True,exist_ok=True)
result.to_csv(out/'aggregation_audit.csv',index=False)
print(result.to_string(index=False))
```
