## Supplementary material for "Calibration and processing sensitivity of time-embedded three-dimensional electrocardiographic descriptors: a two-dataset benchmark": Research materials and reproducible analysis code: README.md

# Reproducible research materials

This archive supports the manuscript Calibration and processing sensitivity of time-embedded three-dimensional electrocardiographic descriptors: a two-dataset benchmark. It contains the original and explicitly post hoc numerical analyses, complete saved outputs, protocols, selected record identifiers, source manifests, and supplementary table descriptions. It does not contain source ECG waveforms, published reference articles, editorial correspondence or document-assembly utilities. The scripts retrieve the original public datasets from PhysioNet with source version and checksum checks.

Preserve the five sibling study directories. Use Python 3.12 and the package versions in estudio_3decg_reproducibilidad/requirements.txt. Run from the extracted archive root. To retrieve the original recordings and annotations:

    python estudio_3decg_reproducibilidad/download_data.py
    python estudio_3decg_paper/download_external.py

To reproduce the original benchmark:

    python estudio_3decg_paper/analysis.py calibration
    python estudio_3decg_paper/analysis.py LUDB
    python estudio_3decg_paper/analysis.py PTBXL
    python estudio_3decg_paper/summarize_verify.py
    python estudio_3decg_paper/verify_calibration_and_units.py

For the first post hoc extensions:

    python estudio_3decg_revision1/revision_analysis.py transfer
    python estudio_3decg_revision1/revision_analysis.py tables
    python estudio_3decg_revision1/revision_analysis.py LUDB
    python estudio_3decg_revision1/revision_analysis.py PTBXL
    python estudio_3decg_revision1/revision_analysis.py calibration
    python estudio_3decg_revision1/revision_analysis.py summarize
    python estudio_3decg_revision1/verify_revision.py
    python estudio_3decg_revision1/figures_revision.py

For the second post hoc annotation and aggregation audits:

    python -c "from estudio_3decg_revision2.audit_revision2 import borders, decomposition; borders(); decomposition()"

The original clinical dataset includes 200 LUDB participants (1,748 QRS windows) and 1,000 PTB-XL participants (9,735 QRS windows). Coordinates use V1/(1 mV), V6/(1 mV), and time/(1 ms). Internal key PTBXL means PTB-XL. Undefined measurements are retained and counted; common-valid-window analyses are separately identified. Analytic controls are mathematical trajectories, not patients. No diagnostic superiority is established.

Original numerical code and outputs are preserved byte-for-byte. A subsequent explicitly post hoc matching-tolerance and duration audit is included in estudio_3decg_revision3; it does not alter the original descriptor measurements. Existing provenance records describe the original execution, including original hashes and timestamps. Document metadata cleanup does not change or replace scientific provenance. The locally hashed plans were fixed before the added analyses but after the original results; this is not public preregistration. Separate computational checks are not review by an independent research team. Downloading data requires internet access; examining existing outputs does not.

LUDB 1.0.1: https://physionet.org/content/ludb/1.0.1/ (Open Data Commons Attribution 1.0).
PTB-XL 1.0.3: https://physionet.org/content/ptb-xl/1.0.3/ (CC BY 4.0).
PTB-XL+ 1.0.1: https://physionet.org/content/ptb-xl-plus/1.0.1/ (CC BY 4.0).
Original data licensing and attribution remain applicable. No new blanket license has been granted over third-party data. Repository and submission licenses are selected by the author.

FILE_DESCRIPTIONS.csv lists each payload file, byte count, column names where applicable and checksum. SHA256SUMS.json provides the complete payload integrity manifest. A dedicated data-repository DOI has not yet been assigned.

For the additional matching-tolerance and duration checks, run:

    python estudio_3decg_revision3/matching_sensitivity.py

This step reads the included complete annotation landmarks and saved candidate windows, reconstructs mutual-nearest matches, and reports the 80-ms count sensitivity and lead-wise duration decomposition. It does not require downloading waveforms or recomputing descriptors.

To check paired percentage changes against changes in cohort medians without downloading data, run python estudio_3decg_revision4/verify_aggregation.py. Random seeds are fixed integers, not execution dates. Existing provenance timestamps and hashes are unchanged.

The distributed estudio_3decg_paper/PROTOCOL.md omits one non-methodological editorial instruction. Its protocol_lock.json retains the original timestamp and hash, which apply to the unabridged source. SHA256SUMS.json provides the checksums of the actual distributed files. Analysis code and numerical outputs are unchanged.
