## Supplementary material for "Calibration and processing sensitivity of time-embedded three-dimensional electrocardiographic descriptors: a two-dataset benchmark": Research materials and reproducible analysis code: Supplementary_captions.txt

Supplementary_methods.pdf
Exact methods, post hoc extensions, printed calibration tables and supplementary figures.

planned_comparisons.csv
Supplementary Table S1. Original planned contrasts.

geometry_comparisons.csv
Supplementary Table S2. Original coordinate audit.

boundary_sensitivity.csv
Supplementary Table S3. Original boundary and extension sensitivity.

extension_sensitivity.csv
Supplementary Table S3. Original boundary and extension sensitivity.

calibration_summary.csv
Supplementary Table S4. Original calibration: 576 aggregate rows and all realization estimates.

calibration_targets.csv
Supplementary Table S4. Original calibration: 576 aggregate rows and all realization estimates.

calibration_estimates.csv
Supplementary Table S4. Original calibration: 576 aggregate rows and all realization estimates.

operator_bandwidths.csv
Supplementary Table S5. Native and matched transfer functions, normalized crossings and numerical harmonic checks.

operator_responses.csv
Supplementary Table S5. Native and matched transfer functions, normalized crossings and numerical harmonic checks.

transfer_numerical_checks.csv
Supplementary Table S5. Native and matched transfer functions, normalized crossings and numerical harmonic checks.

expanded_absolute_descriptors.csv
Supplementary Table S6. Absolute descriptor medians and interquartile ranges by cohort, operator, cutoff and matched status; additional rows describe index decomposition.

extended_calibration_summary.csv
Supplementary Table S7. Expanded calibration: all 1200 aggregate rows, both targets where exact, all 80400 scalar estimates; transient waveform and derivatives.

extended_calibration_targets.csv
Supplementary Table S7. Expanded calibration: all 1200 aggregate rows, both targets where exact, all 80400 scalar estimates; transient waveform and derivatives.

extended_calibration_estimates.csv
Supplementary Table S7. Expanded calibration: all 1200 aggregate rows, both targets where exact, all 80400 scalar estimates; transient waveform and derivatives.

gaussian_control_exact.csv
Supplementary Table S7. Expanded calibration: all 1200 aggregate rows, both targets where exact, all 80400 scalar estimates; transient waveform and derivatives.

inclusion_flow.csv
Supplementary Table S8. Inclusion, full window-duration frequencies and distributions at window and participant levels.

duration_summary.csv
Supplementary Table S8. Inclusion, full window-duration frequencies and distributions at window and participant levels.

duration_histogram.csv
Supplementary Table S8. Inclusion, full window-duration frequencies and distributions at window and participant levels.

duration_windows_LUDB.csv
Supplementary Table S8. Inclusion, full window-duration frequencies and distributions at window and participant levels.

duration_windows_PTBXL.csv
Supplementary Table S8. Inclusion, full window-duration frequencies and distributions at window and participant levels.

common_duration_counts.csv
Supplementary Table S9. Post hoc 80–160 ms duration restriction: counts, all contrasts and participant summaries.

common_duration_comparisons.csv
Supplementary Table S9. Post hoc 80–160 ms duration restriction: counts, all contrasts and participant summaries.

common_duration_participants_LUDB.csv
Supplementary Table S9. Post hoc 80–160 ms duration restriction: counts, all contrasts and participant summaries.

common_duration_participants_PTBXL.csv
Supplementary Table S9. Post hoc 80–160 ms duration restriction: counts, all contrasts and participant summaries.

index_decomposition_comparisons.csv
Supplementary Table S10. Almost-curvature slope, limiting-expression and classical-curvature comparisons; per-window and participant decompositions.

index_decomposition_beats_LUDB.csv
Supplementary Table S10. Almost-curvature slope, limiting-expression and classical-curvature comparisons; per-window and participant decompositions.

index_decomposition_beats_PTBXL.csv
Supplementary Table S10. Almost-curvature slope, limiting-expression and classical-curvature comparisons; per-window and participant decompositions.

index_decomposition_participants_LUDB.csv
Supplementary Table S10. Almost-curvature slope, limiting-expression and classical-curvature comparisons; per-window and participant decompositions.

index_decomposition_participants_PTBXL.csv
Supplementary Table S10. Almost-curvature slope, limiting-expression and classical-curvature comparisons; per-window and participant decompositions.

additional_comparisons.csv
Supplementary Table S11. All post hoc operator contrasts, cutoff heuristics, invalid-value counts and common-valid-window follow-up.

spec_fd_all_cutoffs.csv
Supplementary Table S11. All post hoc operator contrasts, cutoff heuristics, invalid-value counts and common-valid-window follow-up.

cutoff_edge_heuristic.csv
Supplementary Table S11. All post hoc operator contrasts, cutoff heuristics, invalid-value counts and common-valid-window follow-up.

additional_invalid_counts.csv
Supplementary Table S11. All post hoc operator contrasts, cutoff heuristics, invalid-value counts and common-valid-window follow-up.

matched_common_valid_comparisons.csv
Supplementary Table S11. All post hoc operator contrasts, cutoff heuristics, invalid-value counts and common-valid-window follow-up.

additional_beats_LUDB.csv
Supplementary Table S11. All post hoc operator contrasts, cutoff heuristics, invalid-value counts and common-valid-window follow-up.

additional_beats_PTBXL.csv
Supplementary Table S11. All post hoc operator contrasts, cutoff heuristics, invalid-value counts and common-valid-window follow-up.

additional_participants_LUDB.csv
Supplementary Table S11. All post hoc operator contrasts, cutoff heuristics, invalid-value counts and common-valid-window follow-up.

additional_participants_PTBXL.csv
Supplementary Table S11. All post hoc operator contrasts, cutoff heuristics, invalid-value counts and common-valid-window follow-up.

Figure1_data.csv
Exact numerical source data for the corresponding figure.

Figure2_data.csv
Exact numerical source data for the corresponding figure.

Figure3_data.csv
Exact numerical source data for the corresponding figure.

Figure4_duration_data.csv
Exact numerical source data for the corresponding figure.

Figure4_flow_data.csv
Exact numerical source data for the corresponding figure.

FigureS1_data.csv
Exact numerical source data for the corresponding figure.

FigureS2_data.csv
Exact numerical source data for the corresponding figure.

Research_materials.zip
Source-record identifiers, download scripts, versions and licenses, original protocols, post hoc revision plan and hash, source code, all numerical outputs and integrity manifest. Mathematical controls are distinguished from real ECG recordings.

aggregation_decomposition_participants.csv
Supplementary Table S10. Second-review verification of annotation timing and guards, aggregation or transient error as named. These audits do not change original measurements.

aggregation_decomposition_summary.csv
Supplementary Table S10. Second-review verification of annotation timing and guards, aggregation or transient error as named. These audits do not change original measurements.

annotation_coverage.csv
Supplementary Table S12. Second-review verification of annotation timing and guards, aggregation or transient error as named. These audits do not change original measurements.

annotation_coverage_summary.csv
Supplementary Table S12. Second-review verification of annotation timing and guards, aggregation or transient error as named. These audits do not change original measurements.

guard_records.csv
Supplementary Table S12. Second-review verification of annotation timing and guards, aggregation or transient error as named. These audits do not change original measurements.

guard_candidates.csv
Supplementary Table S12. Second-review verification of annotation timing and guards, aggregation or transient error as named. These audits do not change original measurements.

annotated_qrs_landmarks.csv
Supplementary Table S12. Second-review verification of annotation timing and guards, aggregation or transient error as named. These audits do not change original measurements.

annotation_time_histogram.csv
Supplementary Table S12. Second-review verification of annotation timing and guards, aggregation or transient error as named. These audits do not change original measurements.

cross_lead_peak_differences.csv
Supplementary Table S12. Second-review verification of annotation timing and guards, aggregation or transient error as named. These audits do not change original measurements.

derivative_ratio_examples.csv
Supplementary Table S13. Second-review verification of annotation timing and guards, aggregation or transient error as named. These audits do not change original measurements.

gaussian_error_cancellation.csv
Supplementary Table S13. Second-review verification of annotation timing and guards, aggregation or transient error as named. These audits do not change original measurements.

matched_panel_difference.csv
Supplementary Table S13. Second-review verification of annotation timing and guards, aggregation or transient error as named. These audits do not change original measurements.

Tables S12b and S12c: matching_tolerance_summary.csv provides the post hoc 100/80-ms matching counts; matching_tolerance_candidates.csv includes all candidate boundaries, separations, eligibility and individual lead durations; lead_duration_decomposition_summary.csv provides eligible-window medians and quartiles. Sections of the supplement are Supplementary Notes 1–11; table numbers remain S1–S13.

aggregation_audit.csv
Supplementary Table S11. Recalculated median paired SPEC-FD changes and percentage changes in cohort medians at 40, 100 and 150 Hz; these are different estimands.
