## Supplementary methods, figures and tables for "Calibration and processing sensitivity of time-embedded three-dimensional electrocardiographic descriptors: a two-dataset benchmark"

### Supplementary methods for calibration of time embedded 3D ECG descriptors

#### Supplementary Note 1. Selection and provenance

The external selection rule ranks SHA-256 digests of the string 3decg-external-20260914: followed by each integer patient identifier. Only the lowest ECG identifier for each person within metadata fold 10 is considered. The first 1,000 patients are selected. The complete selection list, published-checksum records, download manifest, protocol and annotation-reader amendment are included. Source metadata are retrieved from the cited repositories by the download scripts. Data from non-test record 00001 were used only for annotation-format inspection and do not enter the external cohort.

External ECGDeli files contain comment symbols with explicit event labels in aux_note and a 500-Hz sampling-frequency declaration. For each R peak, the reader requires exactly one QRS onset after the preceding R peak and at or before the current peak, and one QRS offset at or after the current peak and before the next peak. The first and last intervals use their available unbounded side. Complete triples are mutually matched between V1 and V6 within 50 samples. Their onset/offset union is retained only if its duration is 20-150 samples and its bounds lie within samples 500-4499, inclusive. Annotation indices are used as stored. This is automated parsing, not a manual reference adjudication.

#### Supplementary Note 2. Fixed derivative operators

Butterworth filtering used scipy.signal.butter(4, cutoff, fs=500, output="sos") and sosfiltfilt with odd padding of 15 samples. FD applied numpy.gradient three successive times, each with delta=2 ms and edge_order=2. SG used scipy.signal.savgol_filter separately for derivative orders 1, 2 and 3, window_length=11, polyorder=3, delta=2 and mode="interp". SG0SPEC used the same polynomial window for order-zero smoothing, followed by SPEC. These implementation details and library versions are part of the measurement specification.

For SPEC, a record $x[0], \ldots, x[N-1]$ was extended with $x[N-2], \ldots, x[1]$, creating length $L = 2N - 2$. The real Fourier transform was computed once. The Nyquist coefficient was set to zero for all derivative orders. For order $k$, each coefficient was multiplied by $(i\omega)^k$, with $\omega = 2\pi f$ and $f$ in cycles/ms, and the inverse real transform was cropped to its first $N$ values. Thus all derivative orders refer to the same trigonometric interpolant. The extension comparison used the original $N$-sample signal as a periodic interval with the same Nyquist convention.

#### Supplementary Note 3. Geometric formulas and interpretation

$$\kappa(u) = \frac{\|\mathbf{r}' \times \mathbf{r}''\|}{\|\mathbf{r}'\|^3}, \qquad \tau(u) = \frac{\det(\mathbf{r}', \mathbf{r}'', \mathbf{r}''')}{\|\mathbf{r}' \times \mathbf{r}''\|^2}. \tag{S1}$$

The time-averaged absolute torsion uses the integral of absolute pointwise torsion divided by QRS duration. It is not the absolute value of mean signed torsion. The weighted alternative integrates the absolute determinant and divides by the integral of the squared cross-product norm. This weights pointwise absolute torsion by its denominator wherever defined; it therefore changes the estimand when torsion varies over the window.

$$K = \frac{\sqrt{(y''z' - y'z'')^2 + (x')^2(z'' - z')^2 + (x''y' - x'y'')^2}}{\left[(x')^2 + (y')^2 + (z')^2\right]^{3/2}}. \tag{S2}$$

The almost-curvature expression above is equation 6 as printed in the 2025 source [main reference 1]. It contains derivative orders that do not scale homogeneously. The original source explicitly describes a reduction of derivative order in the XZ term when defining almost-curvature (page 5, subsection on RT almost-curvature). The source states: "Reducing the order of the second derivative in x within the term that evaluates the XZ plane" ... [reference 1, p. 5, "RT almost-curvature in acute ischemia," immediately before equation 6; https://doi.org/10.1016/j.jelectrocard.2025.153875]. This short excerpt documents the intended derivative-order modification; it does not validate the source's accompanying clinical interpretation. We therefore analyze the printed empirical index without calling it an erratum. Classical geometric invariances cannot be assumed. For the straight line $\mathbf{r}(u) = (u, 0, u)$, classical curvature is zero and this expression equals $1/(2\sqrt{2})$. After the proper voltage-plane rotation to $\mathbf{r}(u) = (0, u, u)$, it equals zero. Straight-line torsion is undefined, not zero. These are exact algebraic controls, not novel physiological results.

$$\tau = \frac{x''y''' - y''x'''}{(x'')^2 + (y'')^2 + (x'y'' - y'x'')^2} = \frac{N/A}{1 + C^2/A}. \tag{S3}$$

For $\mathbf{r}(t) = (x(t), y(t), t)$, set $A = (x'')^2 + (y'')^2$, $C = x'y'' - y'x''$, and $N = x''y''' - y''x'''$. Where $A > 0$, $N/A$ is the angular velocity of the planar second-derivative vector. The correction $C^2/A$ is bounded above by $(x')^2 + (y')^2$. Changing voltage/time scales changes the correction, so this identity does not attach anatomical units to the computed torsion.

#### Supplementary Note 4. Invalid values and aggregation

For pointwise torsion, the squared cross-product norm must exceed machine epsilon times its maximum over the processed record. Other locations are set to missing. A temporal QRS torsion summary is missing if any included sample is undefined; the valid interval fraction is also saved. Curvature can legitimately equal zero. Weighted torsion is defined if its integrated denominator is positive. Per-person medians are calculated from available window summaries, and paired comparisons omit participants missing either member. The invalid-value counts are explicitly reported; a positive denominator is not a clinical reliability guarantee.

#### Supplementary Note 5. Analytic calibration

The three helices use $x(t) = \cos(\omega t)$, $y(t) = \sin(\omega t)$, $z(t) = t$, with $\omega = 2\pi f/1000$, $f = 10$, 30 or 60 Hz, and $t$ in ms. The modulated trajectory uses $\phi(t) = 2\pi\, 20\, t/1000 + 0.8 \sin(2\pi\, 3\, t/1000)$, $x = \cos\phi$, $y = \sin\phi$, $z = t$. Exact derivatives follow by the chain rule. The true descriptor is integrated on the same sampled central interval as the estimate; therefore the comparison includes a common trapezoidal integration convention rather than comparing different QRS grids.

For a helix, the double-pass filter amplitude is $L = |H(f)|^2$. The exact filtered trajectory has radius $L$ and classical torsion $\omega/(1 + L^2\omega^2)$. Filtered curvature and the equation-6 index are calculated from the analytically scaled derivatives. The phase-modulated curve has a known unfiltered analytic target; no exact post-filter target is asserted for it. Both temporal and denominator-weighted targets are computed separately. Noise is independent between coordinates, with a fixed seed of 20260917 and common realizations across operators. The original 804 signal/noise realizations generate 576 aggregated rows: 4 models times 3 noise levels times 3 cutoffs times 4 operators times 4 descriptors. Realizations and aggregated combinations are different counts. The post hoc extension is specified below.

##### Supplementary Note 6. Sensitivity and statistical outputs

The onset/offset perturbations are (-10,-10), (-10,+10), (+10,-10) and (+10,+10) ms. Each variant is summarized per participant; its percentage difference is calculated against the corresponding unperturbed summary. The maximum absolute difference across variants is then summarized across participants. The extension comparison pairs periodic versus reflected SPEC summaries. The coordinate audit retains the voltage/time coordinates during reparametrization: derivatives of order k are multiplied by $1000^k$, including the time-axis derivative. In the separate scale experiment, the time-axis slope is changed to 0.01 with derivatives taken in ms.

Bootstrap intervals use 5,000 paired participant resamples, seed 20260918, and the 2.5th and 97.5th percentiles of the resampled median relative changes. Intervals are descriptive, without adjustment across secondary outcomes. All processing contrasts are calculated within a cohort and participant; they do not estimate a between-cohort physiological effect. The primary external contrast was specified before external descriptor outcomes were inspected.

##### Supplementary Note 7. Transfer functions, regularization and matching

Let $h = 2$ ms and $\omega = 2\pi f/1000$ rad/ms. For interior FD stencils, the normalized response of derivative order $k$ is

$$g_{\mathrm{FD},k}(f) = \left[\frac{\sin(\omega h)}{\omega h}\right]^k . \tag{S4}$$

For direct SG differentiation, with dot-product coefficients $c_{k,n}$, $n = -5, \ldots, 5$, the response is

$$g_{\mathrm{SG},k}(f) = \frac{\sum_{n=-5}^{5} c_{k,n} \exp(i\omega h n)}{(i\omega)^k}. \tag{S5}$$

For SPEC, $g = 1$ below Nyquist. SG0SPEC uses the order-zero SG polynomial response for all derivative orders. These real normalized responses may change sign; bandwidth crossings use their magnitudes. The normalized SG responses have different polynomial kernels, so their first crossings need not decrease with derivative order. At nominal 150 Hz, order 3 crosses at 36.61 Hz and order 2 at 31.80 Hz; at nominal 40 Hz the corresponding values are 30.68 and 28.80 Hz. The ordering concerns their passband shapes, not a necessary consequence of a later sign change. If $B_c(f) = |H_c(f)|^2$ is the double-pass Butterworth response at nominal cutoff $c$, the complete normalized response is $B_c g_{m,k}$. Table S5 reports the first crossing of $1/\sqrt{2}$, rather than the first crossing of the unnormalized derivative magnitude. The latter grows with frequency and does not define the same bandwidth criterion. Figure 3 includes the response curves. The tabulated noise-gain integral is $\int_0^{250} |B_c g_{m,k}(i\omega)^k|^2 df$, numerically integrated on the supplied grid; matching a crossing does not match this integral.

For a helix of post-Butterworth radius $R$, direct substitution of FD harmonic derivatives gives $\widehat{\tau} = s\omega/(1 + R^2 s^2 \omega^2)$, compared with $\tau = \omega/(1 + R^2\omega^2)$. The simpler ratio $\widehat{\tau}/\tau \approx s$ requires a negligible slope correction. In the real ECG audit at FD/150 Hz, the participant median of the window-mean $C^2/A$ was 0.000360 in LUDB and 0.000498 in PTB-XL; the analogous medians of window maxima were 0.00362 and 0.00765. Small correction terms do not turn a multicomponent transient into a single harmonic.

The cutoff-edge heuristic is calculated separately for each person as

$$100\left[\frac{T_{\mathrm{SPEC},c}}{T_{\mathrm{SPEC},150}}\frac{s(c)}{s(150)}-1\right],\qquad c\in\{40,100\}\,\mathrm{Hz},\tag{S6}$$

where $T$ is that person's median QRS temporal absolute torsion. The median of these predictions and the median absolute prediction error against the paired FD change are both retained. This uses observed SPEC values and is not a prospective estimator of true biological torsion.

For TIK, the same even-reflected, Nyquist-removed trigonometric representation as SPEC is used. On that representation, for each voltage coordinate, solve

$$\underset{v}{\text{minimize}}\quad \|v-y\|_2^2+\lambda\|D^3v\|_2^2,\qquad \lambda=\frac{\sqrt{2}-1}{(2\pi\,40/1000)^6}.\tag{S7}$$

Here $D$ is the periodic spectral derivative in ms and the optimization excludes the Nyquist mode. The solution multiplies the remaining Fourier coefficients by $W(f)=[1+(\sqrt{2}-1)(f/(40\,\mathrm{Hz}))^6]^{-1}$. Derivatives of orders 1–3 are then taken from this single regularized curve. This is a squared third-derivative Tikhonov penalty, not a total-variation penalty or a claim of optimal parameter selection.

For matching, retain the 150-Hz Butterworth chain and define

$$\alpha_{m,k}=\log\left[\sqrt{2}\,|B_{150}(20)g_{m,k}(20)|\right],\qquad G_{m,k}(f)=\exp\left[-\alpha_{m,k}\left(\frac{f}{20\,\mathrm{Hz}}\right)^2\right].\tag{S8}$$

All $\alpha$ are nonnegative. Apply $G_{m,k}$ to the even-reflected array of each computed derivative and crop to the original interval. The resulting ideal interior normalized responses first cross $1/\sqrt{2}$ at 20 Hz. The complete response shape is retained as numerical data, including side lobes. Smoothing different derivative orders separately generally breaks common-curve coherence; this is a controlled response comparison, not a new geometrically coherent estimator. For SPEC, smoothness of the even extension is not guaranteed at record endpoints. In particular, a first-derivative jump can prevent uniform convergence of a third-derivative series; the guard interval alone does not prove its absence. The measured reflection/periodicity difference cannot be assigned uniquely to one boundary mechanism.

##### Supplementary Note 8. Transient control and expanded calibration

Each transient coordinate is $x_j(t)=\sum_l a_{jl}\exp[-(t-\mu_{jl})^2/(2\sigma_{jl}^2)]$. Amplitudes are in mV and centers and widths in ms. The first coordinate uses the $(a,\mu,\sigma)$ triples $(-0.15,3975,12)$, $(1,4000,8)$ and $(-0.25,4035,15)$. The second uses $(0.10,3985,14)$, $(0.8,4012,10)$ and $(-0.35,4045,16)$. These fixed coefficients make a localized mathematical control with transient high-frequency content, not a clinically validated morphology. For an individual component $g=a\exp(-z^2/2)$, $z=(t-\mu)/\sigma$,

$$g'=-zg/\sigma,\quad g''=(z^2-1)g/\sigma^2,\quad g^{(3)}=(3z-z^3)g/\sigma^3.\tag{S9}$$

The eight-second 500-Hz signal is evaluated at sample indices 1960–2040, inclusive, or 3920–4080 ms. Figure S1 shows the waveform and derivatives. The unfiltered analytic descriptor is computed by the same trapezoidal convention. No exact post-filter target is asserted for this transient or the phase-modulated control.

The original four models are generated first using the original seed and order; the Gaussian control follows them. Every noisy realization is shared across methods and cutoffs. The five models times (1 + 100 + 100) realizations give 1,005. Each realization produces 20 processing configurations times 4 descriptors, or 80,400 scalar estimates. Aggregation gives 5 models times 3 noise levels times 20 configurations times 4 descriptors, or 1,200 rows. None of these calibration estimates was undefined. The original subset of 38,592 scalar estimates was reproduced to a maximum absolute discrepancy of $9.98 \times 10^{-17}$.

Table S7a presents noiseless torsion errors for both helix targets at every cutoff. Table S7b gives the absolute target values. The exact filtered target is the infinite steady-state Butterworth response, whereas the implementation filters a finite record with padding; any residual transient is included in the reported error. The full CSV table additionally reports signed bias, relative RMSE, both nonzero noise levels, all four descriptors and every matched condition. Figure S2 illustrates the matched results against unfiltered targets. For matched and TIK results, post-Butterworth targets deliberately exclude additional smoothing, whose bias remains part of the assessed estimator chain.

**Supplementary Note 9. Index decomposition and common-window analyses**

For $z = t$, write $p = x'$, $q = y'$, $a = x''$, $b = y''$, $C = pb - qa$ and $G = (1 + p^2 + q^2)^{3/2}$. Then

$$K = \frac{\sqrt{p^2 + b^2 + C^2}}{G}, \qquad \kappa = \frac{\sqrt{a^2 + b^2 + C^2}}{G}. \tag{S10}$$

To avoid confusing the scale factor with $b = y''$, let $t = \beta u$. All derivatives of order $k$, including those of the time coordinate, scale by $\beta^k$. Substitution in equation S2 yields

$$K_\beta = \frac{\sqrt{p^2/\beta^2 + b^2 + C^2}}{G}, \qquad \lim_{\beta\to\infty} K_\beta = \frac{\sqrt{b^2 + C^2}}{G}. \tag{S11}$$

The $a^2$ term required by classical curvature is absent from this limit. For $x(t) = t^2$, $y(t) = 0$, $z = t$, the limit is zero but classical curvature is $2/(1 + 4t^2)^{3/2}$. This disproves a general classical-curvature limit. The finite-$\beta$ formula was checked against direct rescaling of all derivatives at $\beta = 1000$.

For every original QRS at FD/150 Hz, the slope proxy $|p|/G$, the limiting expression, $K$, $K_{1000}$ and classical curvature were integrated and then aggregated within participant. The pointwise fraction $p^2/(p^2 + b^2 + C^2)$ was also time averaged and summarized per participant. Its cohort medians were 0.881 in LUDB and 0.824 in PTB-XL. These direct calculations do not assume that a ratio of cohort medians determines term dominance. If $F = p^2/(p^2 + b^2 + C^2)$, then pointwise $K_{\text{slope}}/K = \sqrt{F}$ and $K_\infty/K = \sqrt{1 - F}$. The ratio of window integrals is instead a $K$-weighted average, for example

$$\frac{\int K_\infty(t)\,dt}{\int K(t)\,dt} = \frac{\int \sqrt{b^2 + C^2}\,G^{-1}\,dt}{\int \sqrt{p^2 + b^2 + C^2}\,G^{-1}\,dt} = \frac{\int K(t)\sqrt{1 - F(t)}\,dt}{\int K(t)\,dt}. \tag{S12}$$

The unweighted mean of $F$, a mean of its square root and this weighted ratio are not interchangeable. Omitting $G^{-1}$ would produce another quantity. Finally, the original participant comparison is the ratio of separate medians of window means, not the median of their ratios. Table S10b reports both aggregation routes. The directly comparable limiting/original ratios are 0.149961 and 0.159050; finite reparametrization gives 0.149972 and 0.159062. The medians of window ratios are instead 0.150478 and 0.159880. The small differences among these three approximately 0.15 aggregation routes are explicitly retained rather than assumed to vanish; this statement does not compare them with the unweighted term fraction. In particular, $\sqrt{1-0.881} \approx 0.345$ applies a nonlinear transformation to a cohort summary of unweighted time averages, whereas the integral ratio above weights $\sqrt{1-F(t)}$ by $K(t)$ before participant aggregation. It is not an alternative estimate of the reported ratio 0.149961. At $\beta = 1000$, the median difference from the limiting expression was 0.00948% in LUDB and 0.01144% externally, yet the limiting expression remained materially different from classical curvature (Table S10).

The post hoc 80–160-ms restriction used original boundaries and outcomes, without redelineation or changing operators. Participant medians were recomputed from retained windows for all four descriptors. Four LUDB and 70 PTB-XL participants had no retained windows. Counts, complete duration frequencies, participant-level duration summaries and all restricted contrasts are retained in Tables S8–S9. Restriction to a common interval does not match the empirical distributions or disentangle annotation source from cohort.

The additional matched chains yielded 17 undefined temporal-torsion measurements across six external windows and six participants, as specified in Table S11. Every participant had other valid windows. A follow-up check restricted all five matched operators to the same complete set of 1,748 LUDB and 9,729 PTB-XL windows before participant aggregation. No parameters were adjusted after this check. The primary matched SPEC–FD median result was unchanged at the reported precision.

##### Supplementary Note 10. Verification and complete tables

Independent participant aggregation, duplicate checks, condition totals, selection reconstruction and reproduction of prior LUDB FD values passed. Explicit Fourier summation agreed with the inverse-transform implementation to a maximum absolute discrepancy of $6.47 \times 10^{-16}$. The scalar and vector torsion formulas agreed, the analytic helix had its expected value, and straight-line torsion was undefined. Official source-file checksums, as well as local protocol and script hashes, are retained. These computational cross-checks are not an independent human audit.

Supplementary Table S1 is planned_comparisons.csv (88 complete cohort/descriptor/contrast rows). Table S2 is geometry_comparisons.csv (24 coordinate-audit comparisons). Table S3 comprises boundary_sensitivity.csv and extension_sensitivity.csv. Table S4 is calibration_summary.csv, supplemented by calibration_targets.csv and calibration_estimates.csv for exact targets and all realizations. Per-window and participant-level clinical results, inclusion audits and figure data are also supplied as CSV files. Reference numbers refer to the bibliography of the main manuscript. The supplementary data archive (Research_materials.zip) contains the fixed selection, protocols, numerical code, complete results and file checksums; the supplied download scripts retrieve the original public recordings and annotations. The separate Supplementary_captions.txt file identifies the contents of each supplied supplementary file. The post hoc additions are Table S5 (operator bandwidths and responses), S6 (absolute descriptors), S7 (expanded calibration), S8 (inclusion and durations), S9 (common-duration sensitivity), S10 (index decomposition, with Table S10b distinguishing aggregation routes), S11 (new operator contrasts, cutoff heuristics, a paired-versus-cohort-median aggregation audit and undefined/common-valid-window counts), S12 (annotation coverage, with Tables S12b and S12c reporting tolerance sensitivity and duration decomposition), and S13 (local error cancellation and related control checks). Each table comprises the named CSV files listed in the captions manifest.

Additional checks independently express Gaussian derivatives using Hermite polynomials (maximum discrepancy $1.39 \times 10^{-17}$), solve the Tikhonov normal equations as a dense matrix problem ($1.79 \times 10^{-13}$), and compare noiseless FD helix results with their exact transfer formula

($1.11 \times 10^{-16}$). All 15 matched response crossings agreed with 20 Hz to $3.20 \times 10^{-14}$ Hz in the root-finding check. Participant aggregation and duplicates were checked separately from the analysis pipeline. These checks verify specified numerical operations, not physiological validity or error-free research.

Original bootstrap seed: 20260918; post hoc comparison seed: 20260919. The random seeds 20260917, 20260918 and 20260919 are fixed integer initialization values, not encoded execution dates. The selection salt is a literal reproducibility identifier; neither seeds nor salt replace the execution timestamps and hashes in the provenance records. All intervals use 5,000 paired participant resamples and are descriptive. The revision plan was fixed before its new calculations, after the original results were known. A later common-valid-window check responded to the observed undefined-value counts and is identified as such. Original source results remain available without alteration alongside the revision outputs.

**Supplementary Note 11. Second-review annotation and aggregation audits**

The second-review audits are post hoc checks of the existing files and fixed computations. Original window selections, derivative settings and descriptor estimates were not changed; the tolerance follow-up below counts an alternative selection only. Raw complete annotation triples were parsed independently and mutual-nearest matching was reconstructed with explicit loops. For every participant, the selected onset, offset and both peak indices matched the saved windows exactly. The accepted range was $500 \leq \text{start} < \text{end} \leq 4499$ in both cohorts. LUDB's observed extrema were 502 and 4497; PTB-XL's were 500 and 4499. There were zero retained guard violations. All 63 LUDB exclusions were reproduced (28 initial, 35 final), as were all 2,453 external exclusions (1,321 initial, 1,132 final).

The annotation distribution explains why an assumed 20% loss is inappropriate for LUDB. Complete V1 peaks numbered only 23 in the first second and 23 in the last, versus 211–228 in each interior second. The corresponding V6 counts were 22 and 24, versus 216–229 internally. For PTB-XL, counts were much more evenly distributed: V1 had 1,233 initial, 1,057 final and 1,224–1,260 per interior second. Guard exclusion is defined on the union of onset and offset, so it need not equal the number of peaks outside the guard. The medians of LUDB's first-onset-to-last-offset envelope were 7.277 s (V1) and 7.267 s (V6), with full distributions in Table S12. Such landmark envelopes omit portions of the first and last cycles and do not measure a continuous annotation-coverage interval. We therefore do not assign a fixed 8.3-s annotation duration or infer actual heart rates by dividing annotated candidate counts by ten seconds.

All external records had the same number of complete triples in both lead files, and every triple found a mutual-nearest partner within the fixed tolerance. Of 12,188 matched pairs, only 795 had identical peak indices; across all matched pairs the median absolute separation was 6 ms [IQR 4–10 ms], with a maximum of 98 ms. Only five records had identical complete peak arrays. Thus, complete matching is an observed property of these supplied annotations, not evidence that their lead-specific peak timestamps are identical. LUDB matched pairs had median separation 24 ms [20–30], maximum 92 ms. These are annotation comparisons, not an independent validation of ECGDeli accuracy.

A further post hoc audit repeated mutual-nearest matching from the saved complete annotation triples and reproduced every original matched pair and guard decision. At 500 Hz, 80 ms corresponds to 40 samples, compared with the original 50-sample tolerance. The nearest-neighbor relation is defined before applying the tolerance, so lowering it removes pairs without assigning different partners. A maximum separation alone does not show how many observations are near the boundary. At 80 ms, PTB-XL loses 14 of 12,188 candidate pairs and 11 of 9,735 eligible windows (0.113%); LUDB loses 21 of 1,811 pairs and 19 of 1,748 eligible windows (1.087%).

Two external and four LUDB participants lose at least one window. One participant in each cohort loses all windows, leaving 999 and 199 participants. The original 100-ms analysis remains unchanged. No claim of stability of descriptor contrasts at 80 ms is made, because only counts were recomputed (Table S12b).

For the original eligible windows, annotated V1 duration had median [IQR] 140 [130–154] ms in PTB-XL and 98 [88–108] ms in LUDB; corresponding V6 values were 114 [110–127] and 90 [82–100] ms. The union interval exceeds the longer single-lead interval by a median of 6 [2–10] ms externally and 0 [0–4] ms in LUDB. Table S12c also reports onset and offset differences. These are separately aggregated window-level quantities; differences of cohort medians are not an additive decomposition of the median union duration. Longer individual-lead intervals show that the duration contrast is not explained solely by taking the cross-lead union. Differences between cohorts and annotation procedures remain confounded.

For the Gaussian transient at nominal 150 Hz without noise, let $e(t) = |\widehat{\tau}(t)| - |\tau(t)|$ on the fixed evaluation window and $J = \int |\tau(t)|dt$. Table S13 reports $100 \int \max(e, 0)dt/J$, $100 \int \min(e, 0)dt/J$ and their sum. All integrals use the original sampled trapezoidal convention. SG's contributions are +16.2927% and -16.6721%, yielding a net -0.3794%. Its integrated absolute error, 32.9649%, is a distinct measure of local fidelity. FD contributions are +2.4941% and -4.5880%, yielding -2.0939%; its integrated absolute error is 7.0822%. Thus, the ranking by the error in a temporal summary can reverse the ranking by local absolute error. The example does not establish a general method ordering.

At 20, 30 and 40 Hz, normalized $g_3/g_2$ values are 0.9895, 0.9765 and 0.9584 for FD, versus 1.0332, 1.0800 and 1.1579 for SG. These are harmonic transfer ratios, not effective frequencies for the Gaussian mixture. Agreement of one FD transient summary with a hand-selected harmonic prediction does not independently validate a single-frequency model for the transient. Conversely, the SG summary's small bias does not imply its pointwise derivatives or torsion are equally accurate.

The identical displayed values in Figure S2 are rounded results, not duplicate simulations. For the 30-Hz and 60-Hz helices and the phase-modulated control, adding 20-$\mu$V noise changed RMSE by less than 0.000502 percentage points under every matched chain. Those aggregate errors are dominated by deterministic processing bias. The 10-Hz helix changes by up to 0.00328 points; the Gaussian transient changes by 24.45–35.87 points. The 60-Hz matched SG error is 30.34%, whereas the other methods are 13.99–14.21%; it is not approximately 14% for all five operators. Unrounded values for both panels are supplied in Table S13.

**Table S5. Native normalized derivative-response crossings**

| Operator | Cutoff (Hz) | Order 1 (Hz) | Order 2 (Hz) | Order 3 (Hz) |
|---|---|---|---|---|
| FD | 40 | 35.46 | 34.94 | 34.42 |
| FD | 100 | 83.76 | 73.94 | 64.18 |
| FD | 150 | 108.55 | 79.66 | 65.48 |
| SG | 40 | 35.15 | 28.80 | 30.68 |
| SG | 100 | 58.25 | 31.80 | 36.60 |
| SG | 150 | 58.53 | 31.80 | 36.61 |
| SG0SPEC | 40 | 34.48 | 34.48 | 34.48 |
| SG0SPEC | 100 | 49.27 | 49.27 | 49.27 |
| SG0SPEC | 150 | 49.32 | 49.32 | 49.32 |
| SPEC | 40 | 35.98 | 35.98 | 35.98 |
| SPEC | 100 | 91.82 | 91.82 | 91.82 |
| SPEC | 150 | 141.53 | 141.53 | 141.53 |
| TIK | 40 | 33.59 | 33.59 | 33.59 |
| TIK | 100 | 39.99 | 39.99 | 39.99 |
| TIK | 150 | 40.00 | 40.00 | 40.00 |

All values include the double-pass Butterworth filter. Matching at nominal 150 Hz sets all 15 order/operator crossings to 20.00 Hz. The full response curves, Gaussian taper coefficients and noise-gain integrals are available as CSV data.

**Table S7a. Noiseless helix torsion RMSE against both targets**

| Cutoff (Hz) | Operator | Helix 10 Hz | Helix 30 Hz | Helix 60 Hz |
|---|---|---|---|---|
| 40 | FD | 0.26 / 0.26 | 1.67 / 2.22 | 3.68 / 9.21 |
| 40 | SG | 0.80 / 0.80 | 8.78 / 8.17 | 74.92 / 53.17 |
| 40 | SG0SPEC | 0.002 / 0.002 | 0.86 / 0.29 | 14.21 / 0.01 |
| 40 | SPEC | <0.001 / <0.001 | 0.56 / <0.001 | 14.20 / <0.001 |
| 40 | TIK | <0.001 / <0.001 | 0.95 / 0.38 | 14.21 / 0.01 |
| 100 | FD | 0.26 / 0.26 | 2.20 / 2.20 | 7.03 / 7.21 |
| 100 | SG | 0.80 / 0.80 | 8.20 / 8.20 | 64.18 / 63.86 |
| 100 | SG0SPEC | 0.002 / 0.002 | 0.35 / 0.35 | 10.69 / 10.48 |
| 100 | SPEC | <0.001 / <0.001 | <0.001 / <0.001 | 0.19 / <0.001 |
| 100 | TIK | <0.001 / <0.001 | 0.46 / 0.46 | 13.73 / 13.51 |
| 150 | FD | 0.26 / 0.26 | 2.20 / 2.20 | 7.18 / 7.18 |
| 150 | SG | 0.80 / 0.80 | 8.20 / 8.20 | 64.02 / 64.02 |
| 150 | SG0SPEC | 0.002 / 0.002 | 0.35 / 0.35 | 10.64 / 10.64 |
| 150 | SPEC | <0.001 / <0.001 | <0.001 / <0.001 | 0.001 / <0.001 |
| 150 | TIK | <0.001 / <0.001 | 0.46 / 0.46 | 13.72 / 13.72 |

Each cell reports relative RMSE (%) against the unfiltered target / exact Butterworth-filtered target. The estimates are identical within each cell; only the reference changes. One noiseless realization is used per model. Bias, noisy realizations and all descriptors are retained without display rounding in the complete Table S7 CSV files.

**Table S7b. Absolute helix torsion targets**

| Helix (Hz) | Cutoff (Hz) | Unfiltered | Filtered |
|---|---|---|---|
| 10 | 40 | 0.06258478 | 0.06258478 |
| 10 | 100 | 0.06258478 | 0.06258478 |
| 10 | 150 | 0.06258478 | 0.06258478 |
| 30 | 40 | 0.18202800 | 0.18304980 |
| 30 | 100 | 0.18202800 | 0.18202828 |
| 30 | 150 | 0.18202800 | 0.18202800 |
| 60 | 40 | 0.33007946 | 0.37694184 |
| 60 | 100 | 0.33007946 | 0.33071221 |
| 60 | 150 | 0.33007946 | 0.33008332 |

Targets are dimensionless under the stated normalized coordinates. The filtered target is the steady-state curve after the double-pass Butterworth filter, before additional operator smoothing.

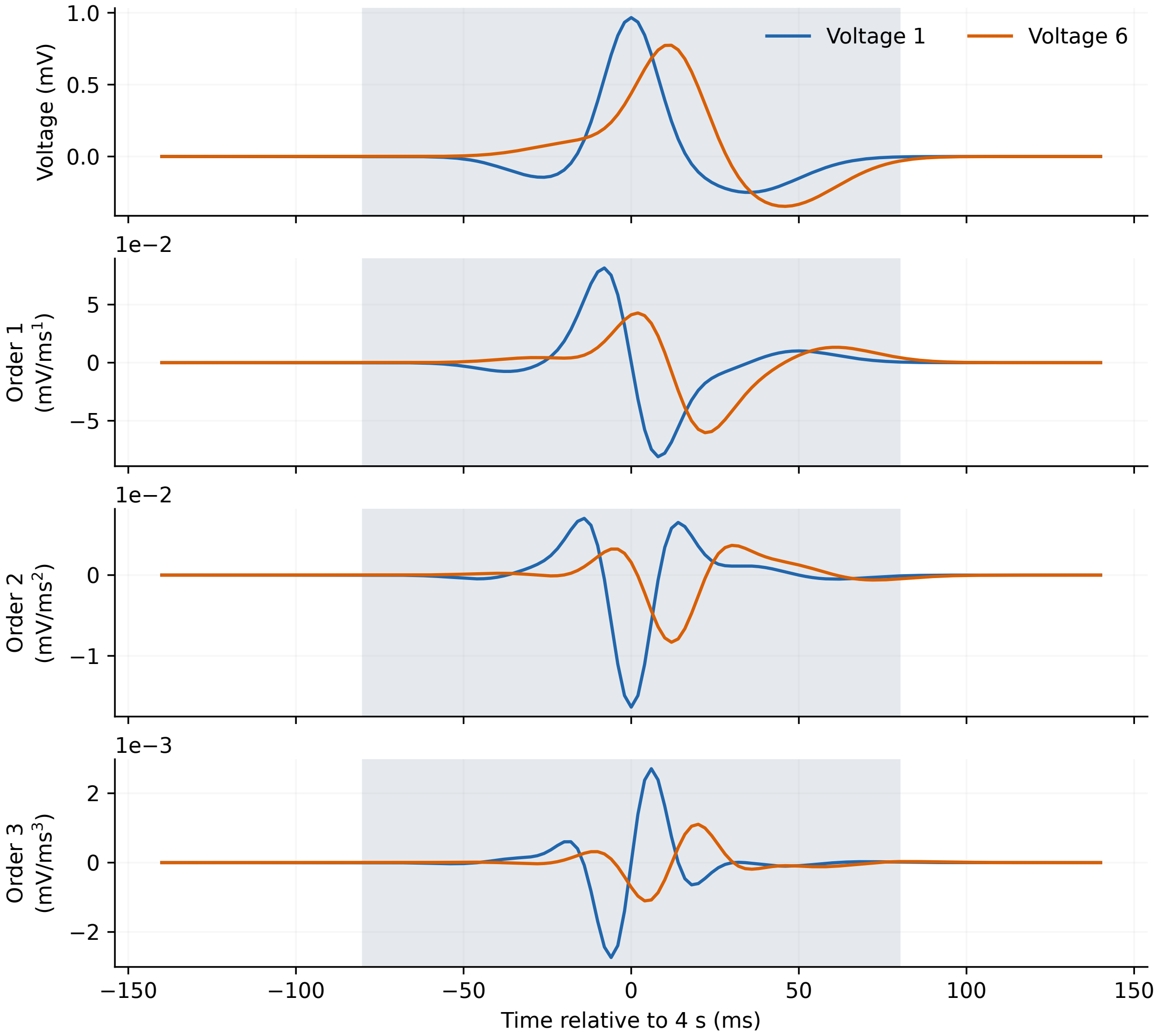


**Figure S1. Transient analytic control.** Two sums of three Gaussian components and their exact first three derivatives, plotted relative to 4 s. Shading marks the 160-ms evaluation interval. Coefficients are specified in Supplementary Note 8; amplitudes and widths were fixed before this control was computed. This mathematical QRS-like transient is not a validated physiological ECG simulator.

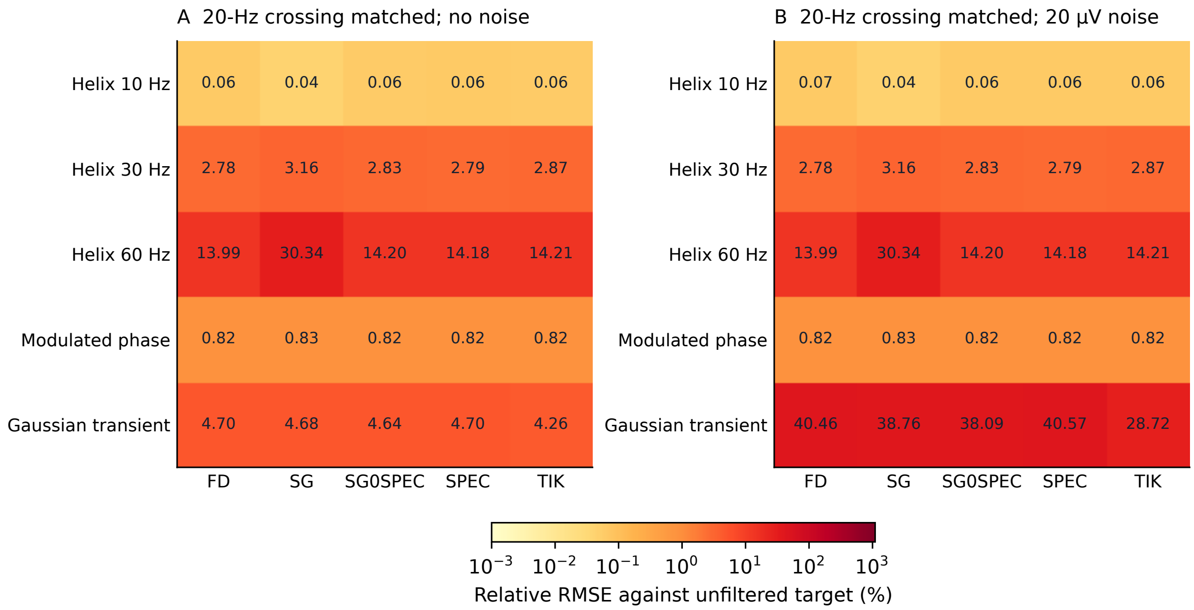


**Figure S2. Calibration after matching the 20-Hz crossing.** Relative RMSE (%) of temporal mean absolute torsion against the unfiltered analytic target. Panel A uses one noiseless realization; panel B uses 100 realizations with 20-$\mu$V noise per coordinate. Every chain starts with the nominal 150-Hz Butterworth filter. The smoothing changes the target curve and can produce substantial noiseless bias, even when the operators closely agree. Identical displayed entries for the 30/60-Hz helices and phase-modulated control reflect rounding of bias-dominated errors; their unrounded values differ by less than 0.000502 percentage points between panels. The matched 60-Hz SG error is 30.34%, not approximately 14% as for the other operators. Full 5-$\mu$V results and signed biases are retained in Table S7.

**Table S10b. Distinct aggregation routes in the index decomposition**

| Quantity | LUDB | PTB-XL |
|---|---|---|
| Unweighted mean pointwise fraction | 0.880740 | 0.824330 |
| Median of window integral ratios | 0.150478 | 0.159880 |
| Ratio of participant medians, limit/original | 0.149961 | 0.159050 |
| Ratio of participant medians, reparam/original | 0.149972 | 0.159062 |
| Ratio of participant medians, slope/original | 0.968312 | 0.962177 |

Each value is a cohort median across participant summaries. Window integral ratios include the common speed denominator. The limiting-index/original-index ratio of participant medians is directly comparable to the original reparametrization contrast; the other aggregation routes need not coincide.

**Table S12. Temporal extent of complete QRS annotations**

| Cohort | Lead | First onset (s) | Last offset (s) | Envelope (s) |
|---|---|---|---|---|
| LUDB | V1 | 1.330 [1.122–1.586] | 8.635 [8.387–8.892] | 7.277 [6.905–7.686] |
| LUDB | V6 | 1.330 [1.120–1.606] | 8.632 [8.386–8.875] | 7.267 [6.894–7.670] |
| PTB-XL | V1 | 0.339 [0.136–0.566] | 9.552 [9.336–9.782] | 9.170 [8.875–9.422] |
| PTB-XL | V6 | 0.334 [0.128–0.560] | 9.529 [9.318–9.754] | 9.157 [8.857–9.408] |

Entries are medians [IQR] across records. Envelope is the last complete offset minus the first complete onset within each record, not the difference between the two displayed cohort medians. It is not a certified continuous annotation-coverage interval. All per-record values, individual candidates and guard decisions are supplied as CSV files.

**Table S12b. Matching-tolerance count sensitivity**

| Cohort | Tolerance (ms) | Matched pairs | Eligible windows | Windows lost | Participants |
|---|---|---|---|---|---|
| LUDB | 100 | 1,811 | 1,748 | 0 | 200 |
| LUDB | 80 | 1,790 | 1,729 | 19 (1.087%) | 199 |
| PTB-XL | 100 | 12,188 | 9,735 | 0 | 1,000 |
| PTB-XL | 80 | 12,174 | 9,724 | 11 (0.113%) | 999 |

Losses are relative to the original 100-ms tolerance. Duration and edge-guard rules are identical. One participant loses all windows in each cohort. This is a post hoc count check; original descriptors and contrasts were not recomputed.

**Table S12c. Lead-wise window-duration decomposition**

| Quantity (ms) | LUDB | PTB-XL |
|---|---|---|
| Absolute peak separation | 24 [20–30] | 6 [4–10] |
| V1 annotated duration | 98 [88–108] | 140 [130–154] |
| V6 annotated duration | 90 [82–100] | 114 [110–127] |
| Union duration | 102 [94–114] | 150 [138–166] |
| Union excess over longer lead | 0 [0–4] | 6 [2–10] |
| Absolute onset separation | 6 [2–12] | 6 [4–10] |
| Absolute offset separation | 8 [4–14] | 28 [14–42] |

Entries are medians [IQR] across the original 1,748 and 9,735 eligible windows. Union excess is calculated within each window before aggregation. Separately reported medians cannot be added or subtracted as an exact decomposition. Annotation method and cohort are not causally separated.

**Table S13. Local error cancellation in the noiseless Gaussian transient**

| Operator | Positive | Negative | Net bias | Absolute error |
|---|---|---|---|---|
| FD | 2.4941 | -4.5880 | -2.0939 | 7.0822 |
| SG | 16.2927 | -16.6721 | -0.3794 | 32.9649 |
| SPEC | 0.0003 | -0.0004 | -0.0001 | 0.0007 |

Values are percentages normalized by the integral of true absolute torsion. Positive and negative contributions sum to the bias in the temporal summary; their difference is the integrated absolute error. All use the same 160-ms window and 150-Hz nominal cutoff. Net bias and local absolute error are different estimands.
